# The role of dopaminergic medication, lipid, and endocannabinoid pathway alterations in idiopathic and *PRKN/PINK1*-mediated Parkinson’s disease – a large-scale targeted metabolomics study

**DOI:** 10.1101/2024.03.01.24303613

**Authors:** Alexander Balck, Max Borsche, Philip Campbell, Xi Luo, John Harvey, Theresa Brückmann, Charlotte Ludwig, Amy Harms, Katja Lohmann, Huw R Morris, Anthony H Schapira, Thomas Hankemeier, Ronan Fleming, Silke Szymczak, Christine Klein

**Author notes:** Corresponding authors: Christine Klein, MD, Institute of Neurogenetics, University of Lübeck, Ratzeburger Allee 160, BMF, 23562 Lübeck, Germany, and Silke Szymczak, Dr., Institute of Medical Biometry and Statistics, Ratzeburger Allee 160, 23562 Lübeck. contributed equally (Alexander Balck, Max Borsche, Silke Szymczak, Christine Klein).

## Abstract

Alterations in many metabolites and biochemical pathways have been described in Parkinson’s Disease (PD). However, only a small fraction of these have been replicated in independent studies. As almost every PD patient is treated with dopaminergic medication, it is challenging to discriminate between disease- and drug-related effects, especially in relation to dopamine metabolism. We conducted a large-scale metabolomic study in plasma from 140 idiopathic (IPD), 19 *PRKN/PINK1*-linked PD patients, and 64 healthy controls to disentangle disease-related metabolite alterations from drug-related effects. We distinguished between L-Dopa and non-L-Dopa treated PD patients to uncover nuanced metabolic changes associated with different therapies. We demonstrate that L-Dopa treatment uniquely influences the metabolome, with methyldopa and methoxytyramine, both L-Dopa breakdown products, elevated in L-Dopa-treated IPD and *PRKN/PINK*-linked PD patients. These alterations were not seen in untreated IPD patients and those on agonist treatment only. Polyamine metabolism alterations, notably elevation of putrescine and ornithine, were partly caused by L-Dopa treatment but also found in non-L-Dopa treated PD patients. In non-L-Dopa-treated patients, endocannabinoid metabolites were lowered and associated with disease duration. We observed lipid metabolism alterations, highlighting potential crosslinks with alpha-synuclein and providing insights into pathophysiological mechanisms. All *PRKN/PINK1-*linked PD patients received L-Dopa treatment. However, our data potentially support the well-established role of oxidative damage in these subtypes of PD. In conclusion, our study emphasizes the significant impact of L-Dopa treatment on the metabolome, which might be of relevance not only for metabolomics studies but also for PD biomarker research in general. Finally, our study highlights potential biomarkers and pathways crucial for the understanding disease mechanisms of PD.

## Introduction

Studies investigating metabolites in the blood of Parkinson’s Disease (PD) patients have suggested alterations in a plethora of metabolites and biochemical pathways.^1^ However, only a small fraction of the suggested metabolites have been replicated in independent studies^2^ Possible explanations lie in major technological and procedural pitfalls that can significantly influence the results and, therefore, must be accounted for in metabolomic studies. First, almost every PD patient is treated with dopaminergic medication, which hampers the discrimination between disease- and drug-related effects; this is especially true of dopamine metabolism. Therefore, adjusting for treatment in statistical analyses may eliminate not only the effect of L-Dopa but also the alterations caused by the disease itself when comparing only L-Dopa-treated PD patients to healthy controls (HC). Second, as metabolite concentrations are very susceptible to handling, processing, and type of analysis, even slight differences in the procedures across laboratories can considerably impact the results.^2^ Third, sex, age, and lifestyle also significantly affect the metabolome.^3^

Adding another level of complexity, PD is a heterogeneous disorder, including several forms of monogenic PD, such as *PRKN*- or *PINK1*-linked PD.^4,5^ Most likely, differences in the underlying molecular course will involve different biochemical pathways, resulting in different levels of specific metabolites for each genetic subset of patients. For example, *PRKN-* and *PINK1*-linked PD involves mitochondrial dysfunction,^6^ making alterations in pathways of oxidative phosphorylation more likely.

An important role has been attributed to lipids in the pathogenesis of PD, as genetic alterations in several genes involved in lipid metabolism have been identified to cause (e.g., *PINK1*) PD or are involved in disease pathogenesis (e.g., VPS13).^7^ Strong evidence suggests that membrane lipids are highly important for alpha-synuclein (α-syn) metabolism, contributing to α-syn fibrilization and accumulation in laboratory models.^8^ Strikingly, α-syn-lipid interactions are likely an essential component in Lewy body formation and, possibly, for spreading pathology.^7^ Among other lipids, levels of phosphatidylcholines, lysophosphatidylcholines, and phosphatidylethanolamines (PE) have been found to discriminate IPD patients^9,10^ and carriers of pathogenic *LRRK2*^11^ variants from HC. However, these lipids have yet to be investigated in detail in biallelic *PRKN*- or *PINK1*-linked PD. Furthermore, investigations of the lipid metabolism in drug-naïve IPD patients are scarce. One study on untreated IPD patients had suggested different free fatty acids, Indolelactic acid, and Phenylacetyl-glutamine to be able to discriminate patients from HC.^12^

Our study addressed two aims: i) To take the influence of dopaminergic treatment on blood-based metabolomics into account, hypothesizing that such medication has a significant impact on metabolomics in idiopathic and monogenic PD patients. ii) To identify dopaminergic medication-independent metabolites distinguishing between idiopathic as well as *PRKN/PINK1*-linked PD and HC by large-scale targeted metabolomics, hypothesizing that distinct alterations in metabolites might serve as biomarkers for (“mitochondrial”) PD.

## Methods

### The SysMedPD study

The Systems Medicine of Mitochondrial Parkinson’s Disease (SysMedPD) study (www.sysmedpd.eu) was initiated by an international consortium, including basic and clinician scientists, to contribute to the elucidation of mitochondrial underpinnings of PD. The consortium consisted of eight partners from five European countries. At the same time, recruitment of patients with idiopathic and different forms of monogenic PD, as well as healthy controls, took place at the Institute of Neurogenetics at the University of Lübeck (UL) and the Department of Neurology at the University College London (UCL). Before recruitment, inclusion and exclusion criteria, the clinical work-up, and the sampling of biomaterials, were harmonized between both centers.

### Proband recruitment

The total sample at the two recruitment sites comprised 289 participants. Inclusion criteria for affected individuals consisted of a definite diagnosis of PD according to the MDS clinical diagnostic criteria for Parkinson’s disease,^13^ eligibility according to the harmonized genetic criteria (see below), and material availability to perform metabolomics analyses. Otherwise, patients with clinical suspicion of atypical PD, co-existing neurodegenerative disorders, diagnosed dementia, or a history of significant infection (defined by the need for antibiotics) or surgery in the two weeks prior to participation were excluded. A flowchart providing the selection process for inclusion in the study is depicted in **Figure 1**. All participants underwent an in-depth clinical assessment for motor symptoms and clinical severity using the Movement Disorder Society Unified Parkinson’s Disease Rating Scale III (MDS-UPDRS III) and the Hoehn and Yahr (H/Y) scale. Of note, we used both scales for association analyses to increase the reliability of the results. Moreover, the Levodopa Equivalent Daily Dosage (LEDD) was calculated as published.^14^

**Figure 1.**
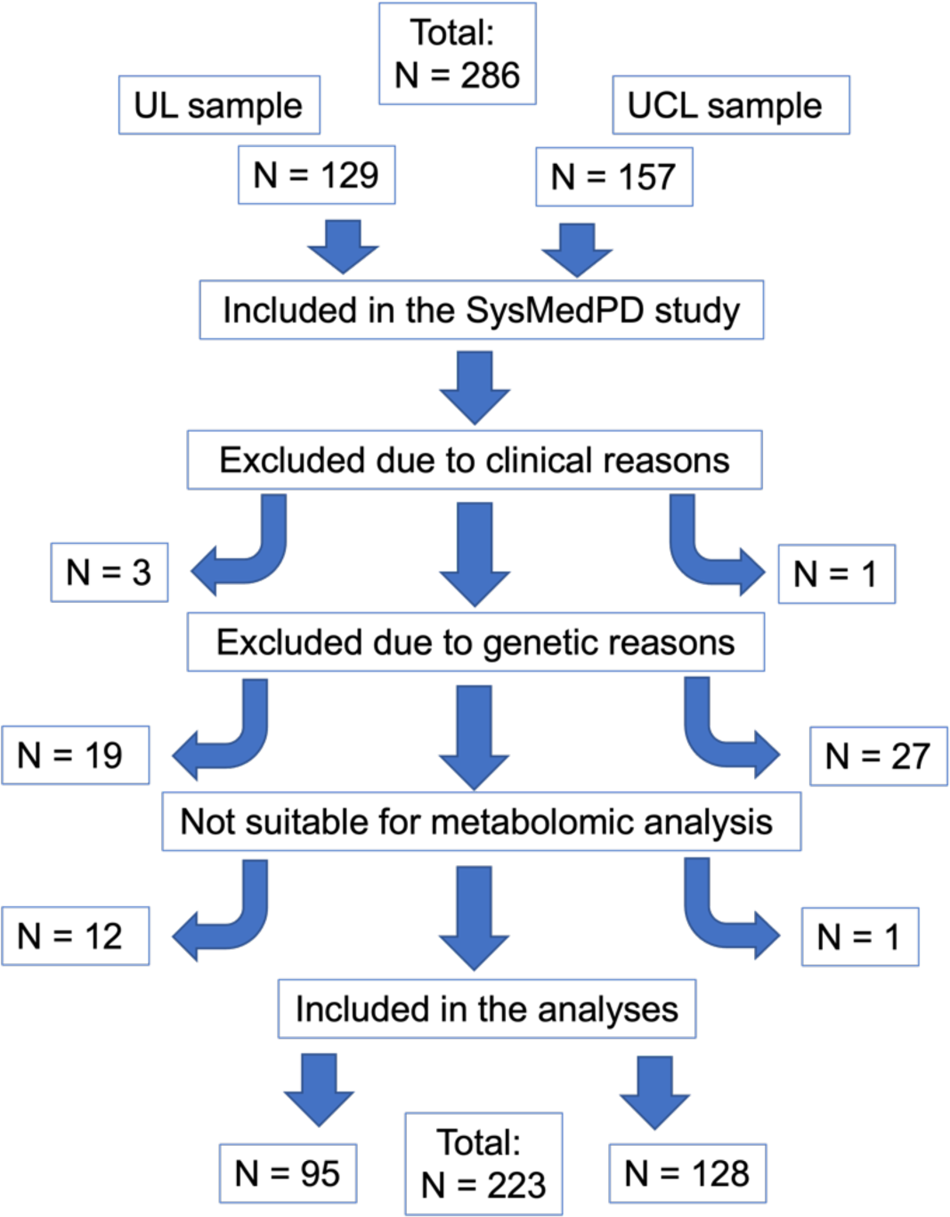
Flowchart of the exclusion and inclusion process. UL = University of Lübeck; UCL: University College London; Excluded due to clinical reasons: Patients who do not fulfill the diagnostic criteria of Parkinson’s disease as described in the methods section; Excluded due to genetic reasons: UL: Patients were excluded due to pathogenic variants/risk variants in *GBA1* (n=10), *LRRK2* (n=2), *SNCA* (n=1), heterozygous variants in *PRKN* or *PINK1* (n=4), and non-conclusive genetic results (n=2). UCL: Patients were excluded due to pathogenic variants/risk variants in *GBA1* (n=12), *LRRK2* (n=7), heterozygous variants in *PRKN* or *PINK1* (n=6), and non-conclusive genetic results (n=2). Patients were not suitable for metabolomic analyses if biomaterials to perform the analyses were not available.

### Genetic Testing

We recruited patients with known pathogenic variants in *PRKN* and *PINK1*, as well as uncovered priorly unknown genetic alterations by genotyping performed within the study. All participants underwent conclusive genetic testing within the study, which differed slightly between both sides. At UL, all samples underwent gene panel sequencing at Centogene (Rostock, Germany). The panel covered the genes *PRKN*, *PINK1*, *DJ-1*, *SNCA*, *LRRK2*, *GBA1*, *VPS35*, *GCH1*, and others irrelevant to the present study.^15^ We confirmed potentially disease-related variants by Sanger sequencing. At UCL, the Illumina NeuroChip array, a customized version of the Infinium HumanCore-24 v1.0 backbone, was used, covering nearly 200,000 variants associated with neurodegenerative diseases, including but not limited to PD.^16^ Furthermore, we performed Multiplex ligation-dependent probe amplification (MLPA) to investigate copy number variations (i.e., deletions or duplications) in PD-associated genes (*SNCA*, *PRKN*, *PINK1*, DJ-1, *LRRK2*, *GCH1*). We performed sequence analyses and MLPA in patients and control participants at both sites. Pathogenicity of detected variants was assessed according to the ACMG (American College of Medical Genetics and Genomics)^17^ and the MDSGene (www.mdsgene.org) criteria.

Genetic study inclusion criteria were likewise harmonized between both study sites: i) We excluded PD patients and controls with pathogenic variants in known PD genes besides pathogenic biallelic *PRKN* and *PINK1* variants. ii) We excluded individuals with heterozygous pathogenic variants in *PRKN* and *PINK1* due to conflicting evidence regarding their role in PD.^18,19^

### Study groups

After the exclusion of screened participants due to either clinical criteria (n=4), genetic criteria (n=46), or due to lack of sufficient biomaterial to perform the experiments (n=13), 223 participants were included. This comprised 140 idiopathic PD patients, 19 *PRKN/PINK1*-linked PD patients (*PRKN*: n=16 individuals; *PINK1*: n=3), and 64 healthy controls undergoing metabolomic analyses (**Figure 1**). Demographics and clinical data are depicted in **Table 1**. Carriers of biallelic pathogenic variants in *PRKN and PINK1* formed the “mitoPD” group for all analyses.

**Table 1.**
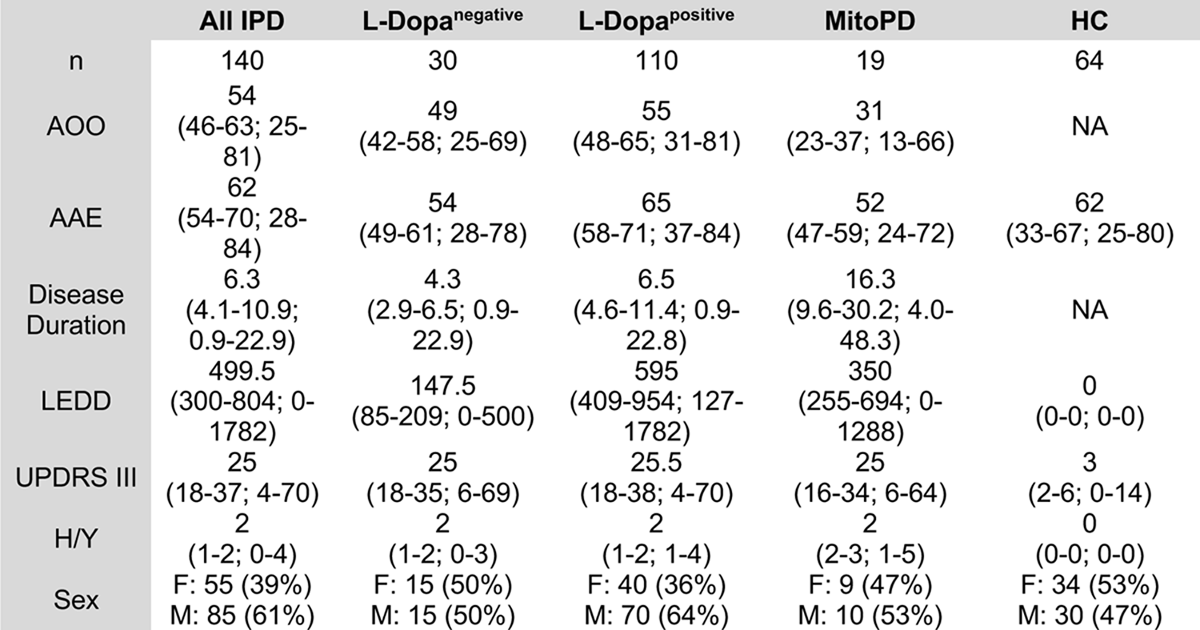
Overview of the study groups: IPD patients were separated into non-L-Dopa treated idiopathic PD (IPD) patients (L-Dopa^negative^) and L-Dopa treated IPD patients (L-Dopa^positive^). L-Dopa^negative^ patients included individuals that were treated with dopamine agonists or untreated. All numerical variables are displayed with mean (1. quartile – 3. quartile; Min. – Max.). MitoPD = *PRKN/PINK1*-linked Parkinson’s disease patients, HC = healthy controls, F = female, M = male, PD = Parkinson’s disease, AAO = Age at onset, AAE = Age at examination, LEDD = Levodopa equivalent daily dosage, UPDRS III = MDS - Unified Parkinson’s Disease rating scale part III, H/Y = Hoehn and Yahr stage

|  | All IPD | L-Dopa<sup>negative</sup> | L-Dopa<sup>positive</sup> | MitoPD | HC |
| --- | --- | --- | --- | --- | --- |
| n | 140 | 30 | 110 | 19 | 64 |
| AOO | 54<br>(46-63; 25-81) | 49<br>(42-58; 25-69) | 55<br>(48-65; 31-81) | 31<br>(23-37; 13-66) | NA |
| AAE | 62<br>(54-70; 28-84) | 54<br>(49-61; 28-78) | 65<br>(58-71; 37-84) | 52<br>(47-59; 24-72) | 62<br>(33-67; 25-80) |
| Disease Duration | 6.3<br>(4.1-10.9; 0.9-22.9) | 4.3<br>(2.9-6.5; 0.9-22.9) | 6.5<br>(4.6-11.4; 0.9-22.8) | 16.3<br>(9.6-30.2; 4.0-48.3) | NA |
| LEDD | 499.5<br>(300-804; 0-1782) | 147.5<br>(85-209; 0-500) | 595<br>(409-954; 127-1782) | 350<br>(255-694; 0-1288) | 0<br>(0-0; 0-0) |
| UPDRS III | 25<br>(18-37; 4-70) | 25<br>(18-35; 6-69) | 25.5<br>(18-38; 4-70) | 25<br>(16-34; 6-64) | 3<br>(2-6; 0-14) |
| H/Y | 2<br>(1-2; 0-4) | 2<br>(1-2; 0-3) | 2<br>(1-2; 1-4) | 2<br>(2-3; 1-5) | 0<br>(0-0; 0-0) |
| Sex | F: 55 (39%)<br>M: 85 (61%) | F: 15 (50%)<br>M: 15 (50%) | F: 40 (36%)<br>M: 70 (64%) | F: 9 (47%)<br>M: 10 (53%) | F: 34 (53%)<br>M: 30 (47%) |

### Pre-analytic sample processing

Blood samples for metabolomic and genetic analysis were collected from each participant at 8am in the morning after a 12-hour fasting period, where participants were only allowed to take medication and drink water. Following the venous blood collection, the samples were placed on ice and promptly conveyed to the laboratory within 15 minutes. The specimens underwent centrifugation at a force of 2000x g for 10 minutes at a controlled temperature of 4°C (Centrifuges: Beckman Coulter Allegra X-15R & Eppendorf centrifuge 5417R).

Subsequently, the resulting plasma supernatants were transferred into a fresh 15ml-Falcon tube, pre-chilled on ice, and subjected to thorough mixing through inversion. Following this step, aliquots of 300ml each were prepared, rapidly cryopreserved by immersion in liquid nitrogen, and ultimately stored at a temperature of −80°C. Specimens from UL and UCL were shipped on dry ice to the University of Leiden under strictly temperature-controlled conditions.

### Metabolomic analyses

Metabolites were assessed with five different metabolite platforms that are described below:

i. The amine platform^20^ covers amino acids and biogenic amines employing an AccQ-Tag derivatization strategy adapted from the protocol supplied by Waters. 5.0μL of each sample was spiked with an internal standard solution. Then, proteins were precipitated by adding MeOH and taken to dryness in a speed vac centrifuge. The residue was reconstituted in borate buffer (pH 8.8) with AQC reagent. 1.0μL of the reaction mixture was injected into the UPLC-MS/MS system. Chromatographic separation was achieved by an Agilent 1290 Infinity II LC System on an AccQ-Tag Ultra column (Waters) with a flow of 0.7mL/min over an 11-min gradient. The UPLC was coupled to electrospray ionization on a triple quadrupole mass spectrometer (AB SCIEX QTRAP 6500). Analytes were detected in the positive ion mode and monitored in Multiple Reaction Monitoring (MRM) using nominal mass resolution. Acquired data were evaluated using MultiQuant Software for Quantitative Analysis (AB SCIEX, Version 3.0.2) by integrating assigned MRM peaks and normalizing using internal standards.
ii. The acylcarnitine platform covers acylcarnitines and Trimethylamine-N-oxide, Choline, Betaine, and Carnitine. Ten µL of each sample were spiked with an internal standard solution, and proteins were precipitated by adding MeOH. The supernatant was transferred to an autosampler vial, and 1.0μL was injected into the UPLCMS/MS. Chromatographic separation was achieved by an Agilent 1290 Infinity II LC System on an Accq-Tag Ultra column (Waters) with a flow of 0.7mL/min over an 11-minute gradient. The UPLC was coupled to electrospray ionization on a triple quadrupole mass spectrometer (AB SCIEX Qtrap 6500). Analytes were detected in the positive ion mode and monitored in Multiple Reaction Monitoring (MRM) using nominal mass resolution. Acquired data were evaluated using MultiQuant Software for Quantitative Analysis (AB SCIEX, Version 3.0.2) by integrating assigned MRM peaks and normalizing using the internal standards.
iii. The organic acid profiling platform, performed with GC-MS technology, covers 28 organic acids. Sample preparation was done by adding the first protein precipitation of 50µL plasma with MeOH/H2O with ISTD. After centrifugation and transferring the supernatant, the samples were evaporated to complete dryness in the speedvac. Then, two-step derivatization procedures were performed online: oximation using methoxyamine hydrochloride (MeOX, 15mg/mL in pyridine) as first reaction and silylation using N-Methyl-N-(trimethylsilyl)-trifluoroacetamide (MSTFA) as second reaction were carried out. One μL of each sample was injected into the GC-MS. The metabolites were measured by gas chromatography on an Agilent Technologies 7890A equipped with an Agilent Technologies mass selective detector (MSD 5975C) and MultiPurpose Sampler (MPS, MXY016-02A, GERSTEL). Chromatographic separations were performed on an HP-5MS UI (5% Phenyl Methyl Silox), 30mÅ∼ 0.25 m ID column with a film thickness of 25μm, using helium as the carrier gas at a flow rate of 1.7mL/min. The mass spectrometer was operated in SCAN mode with a mass range of 50-500. Raw data were pre-processed using Agilent MassHunter Quantitative Analysis software (Agilent, Version B.05.01).
iv. The signaling lipids platform^21^ covers various isoprostane classes and their respective prostaglandin isomers from different polyunsaturated fatty acids. Also included in this platform are endocannabinoids, bile acids, and lipids from the sphingosine and sphinganine classes and their phosphorylated forms, as well as three classes of lysophosphatidic acids. The signaling and peroxidized lipids platform is divided into two chromatographic methods: low and high pH. Each sample was spiked with antioxidant and internal standard solution. The extraction of the compounds is performed via LLE. Butanol and Methyl tert-butyl ether extract the analytes from the aqueous phase. The organic phase is concentrated by drying, reconstituted, transferred into amber autosampler vials, and used for high and low pH injection. For the high pH method, a Kinetex EVO column (Phenomenex) was used on a Shimadzu UPLC system formed by three high-pressure pumps coupled online with an LCMS-8050 triple quadrupole mass spectrometer (Shimadzu). The acquired data was evaluated using LabSolutions Insight software (Version 3.3, Shimadzu). The low pH method used an Acquity UPLC BEH C18 column (Waters) on a Shimadzu UPLC system coupled to a QTRAP mass spectrometer (SCIEX). Analytes were monitored in dynamic Multiple Reaction Monitoring (dMRM) and evaluated using MultiQuant (version 3.0.2).
v. The lipidomics platform^22^ covers 185 compounds, including triglycerides, cholesterol esters, and phospholipids. 1000μL IPA containing internal standards were added to 10μL plasma. 2.5μL was injected on an HSS T3 column on an ACQUITY UPLC™ (Waters, Ettenleur, the Netherlands) with a 16 minutes gradient. The lipid analysis is performed on a UPLC-ESI-Triple-TOF (Sciex 6600+) high-resolution mass spectrometer using reference mass correction. Lipids were detected in full scan in the positive ion mode. MultiQuant Software pre-processed the raw data for Quantitative Analysis (AB SCIEX, Version 3.0.2). The lipid response was calculated as the peak area ratios of the target analyte to the respective internal standard.

For all metabolomic platforms, in-house developed algorithms were applied using pooled quality-controlled samples to compensate for shifts in the sensitivity of the mass spectrometer over batches.

### Statistics

Metabolite data were further preprocessed and quality controlled. If a metabolite was available on several platforms, measurements from the platform with the lowest relative standard deviation based on quality-control samples were kept. Missing values were replaced by a constant metabolic-specific value defined as 0.5 * minimally observed value. A log transformation (base2) was applied to make the distribution of the intensities of each metabolite more symmetric. Following this transformation, the z score was calculated for each metabolite to achieve a distribution with a mean of 0 and a standard deviation of 1, using the mean value of intensities of each metabolite divided by the standard deviation, facilitating the comparison of metabolites.

Additional quality control steps included missing frequencies per metabolite and proband, comparison of intensity distributions across subjects, principal component analysis (PCA) and checks for duplicate samples as implemented in the R package QCnormSE (version 0.99.4.9000) available on GitHub (https://github.com/szymczak-lab/QCnormSE). No metabolites or samples were excluded due to quality issues.

However, we observed a strong batch effect of recruitment sites on the overall distribution of the metabolite intensities in the PCA plots, especially for amines and lipid metabolites. Thus, we first applied a linear regression analysis for each metabolite separately, with transformed metabolite intensity as the dependent variable and recruitment site as the variable of interest and adjusted for age at examination, sex, and diagnosis group (IPD or control) as independent variables. P-values for the effect of the recruitment site were adjusted for multiple tests using the Benjamini-Hochberg procedure.^23^ Since many metabolites showed a significant association, we conducted all further association analyses of metabolite intensities with clinical variables stratified by recruitment site.

Again, we used a linear regression model for each metabolite separately, with transformed metabolite intensity as the dependent variable and the variable of interest (categorical or continuous) as the independent variable and included additional covariates. Details on the participants and covariates included in each analysis as well as the corresponding tables and figures can be found in the **Supplementary Methods.** In particular, most of the analyses were adjusted for age at examination. However, we did not adjust for age at examination when analyzing disease severity measured as H/Y stage and MDS-UPDRS III because of the significant overlap between increasing age and disease severity in PD patients that is similar to the overlap of PD status and levodopa treatment. Therefore, metabolites associated with age at examination and disease severity will be separately discussed.

Regression coefficients (betas) and standard errors of the variable of interest estimated for the two recruitment sites were combined in a fixed meta-analysis using the R package metafor (version 3.8-1)^24^ and P-values were adjusted for multiple testing using the Benjamini-Hochberg procedure.^23^ An association between a clinical variable and a metabolite was considered significant if the corresponding adjusted p-value < 0.05. We report regression coefficients (betas) of the meta-analysis with corresponding 95% confidence intervals (CI) in the results section.

## Results

### Study population

The study sample at both study sites consists of a total of 223 participants. The 140 idiopathic PD patients (55 females (39%)) had a median age at examination of 62 years (interquartile range (IQR) 54-70 years, range 28-84 years), a median age at onset of 54 years (IQR 46-63 years, range 25-81 years), and a median disease duration of 6.3 years (IQR 4.1-10.9 years, range 0.9-22.9 years). The 19 *PRKN/PINK1*-linked PD patients (9 females (47%)) had a median age at examination of 52 years (IQR 47-59 years, range 24-72 years), a median age of onset of 31 years (IQR 23-37 years, range 13-66 years), and a median disease duration of 16 years (IQR 9.6-30.2, range 4.0-48.3 years). The healthy control group comprised 64 individuals with a median age at examination of 62 years (IQR 33-67 years, range 25-80 years). Clinical scores and LEDD are shown in **Table 1**.

### Metabolomics

After preprocessing and quality control, 304 metabolites were available for analysis of which 32, 51, 9, 138 and 74 were acylcarnitines, amines, organic acids, lipids and signaling lipids, respectively (see **Supplementary Table 1** for an overview of all measured metabolites and their annotation).

In the exploration of recruitment site as batch effect, 100 metabolites had an adjusted p-value < 0.05 for recruitment site. However, only 17 significant metabolites had an absolute effect size > 1 (**Supplementary Table 2**). As described in the methods section, all further analyses were stratified for recruitment site and presented results correspond to the effects and p-values of a fixed effect meta-analysis combining the effects of the two strata (UL, UCL).

When combining IPD patients and HCs, 30 out of the 304 metabolites were significantly associated with age at examination (**Supplementary Table 3**). Regarding amino acids, we found Tyrosine, Cysteine, its precursors Cystathionine, and O-Acetylserine, as well as L-Homocitrulline, to be elevated. Two amino acids, Tryptophan and Threonine, were significantly reduced with increasing age. Five acylcarnitines were significantly elevated with age (Isobutyrylcarnitine, Tiglylcarnitine, Trimethylamine N-oxide, Tetradecanoylcarnitine, and Nonanoylcarnitine). and three sphingomyelins with different lipid-lengths were significantly reduced with age (Sphingomyelin(d18:1/22:0), (d18:1/24:0), and (d18:1/24:1)). Notably, we also found Citrate and Isocitrate, both part of the citric acid cycle, to be significantly elevated with increasing age.

### Analyses

Due to the site effects described in the methods section, the final analyses reported in the text refer to a meta-analysis. The figures show the individual level data for the specific metabolites; thus, we did not include any forest plots summarizing the site specific and combined effect estimates.

### L-Dopa treatment but not dopamine agonist treatment increases L-Dopa-dependent metabolites in IPD

First, we explored differences in metabolites comparing all IPD patients (n=140) and HC. Here, we identified five metabolites, i.e., 3-methoxytyramine, Methyldopa, Putrescine, (+/-)-16-Hydroxydocosahexaenoic acid (16-HDoHE), and Ornithine, as the only significant metabolites if corrected for multiple testing (**Table 2, Supplementary Table 4**). We observed a heterogenous distribution within the IPD group in particular for 3-Methoxytyramine and Methyldopa, as these metabolites were predominantly increased in L-Dopa treated individuals (in the following referred to as L-Dopa^positive^), but not similarly in untreated IPD patients and individuals only treated with dopamine agonists (**Figure 2**). Concluding that L-Dopa treatment is responsible for these striking differences between IPD patients and HC, we continued analyzing the L-Dopa^positive^ and the L-Dopa^negative^ group separately, to elucidate differences in the metabolome not caused by dopaminergic treatment. The demographical and clinical data of the two IPD subgroups are likewise displayed in **Table 1**.

**Figure 2.**
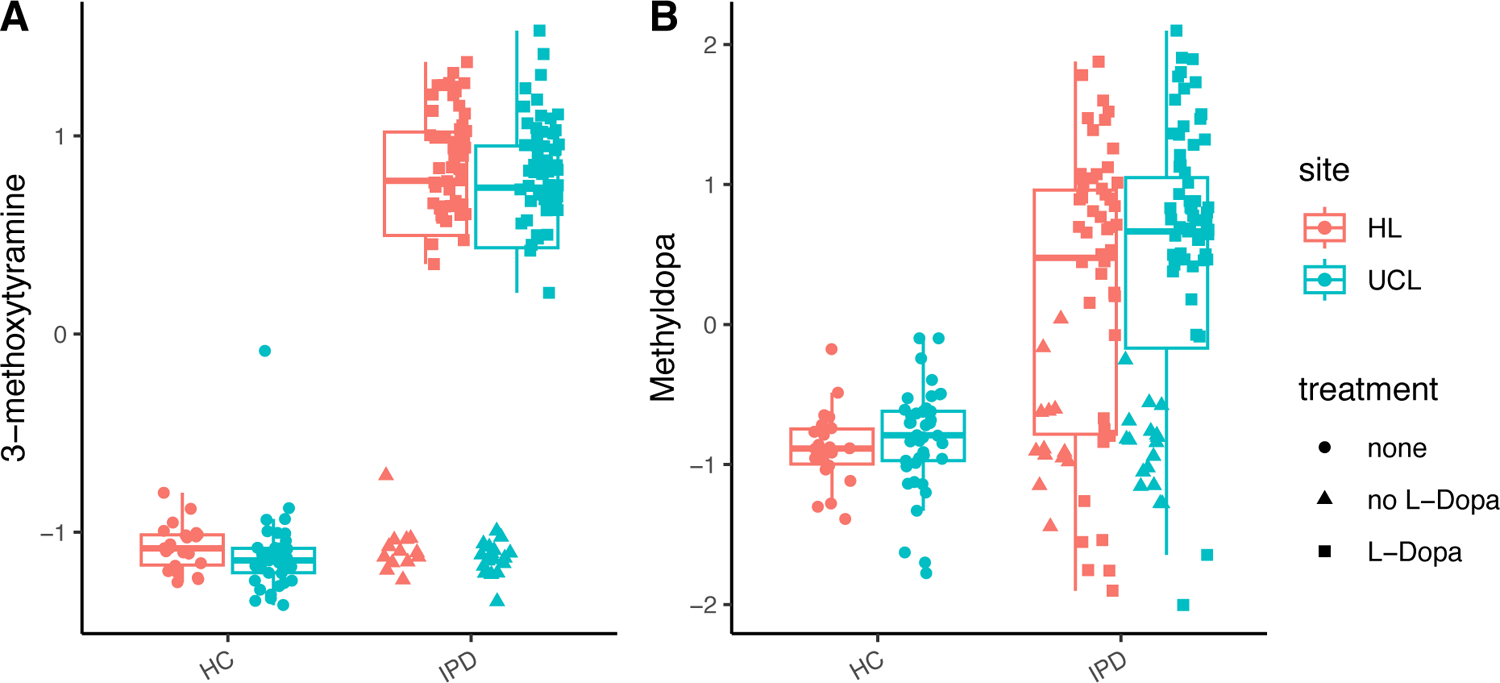
Levels of 3-methoxytyramine (A), methyldopa (B) are increased in idiopathic Parkinsons’s disease (IPD) patients treated with L-Dopa (L-Dopa^positive^, squares) but not elevated in untreated patients (dots) and patients with agonist treatment only (L-Dopa^negative^, triangles). HC = healthy controls; Data were analyzed as described in the statistic section. No significance levels are depicted in the figure, as data uncorrected for site effects is shown, while statistical calculations were performed by meta-analysis as described in the methods part.

**Table 2.**
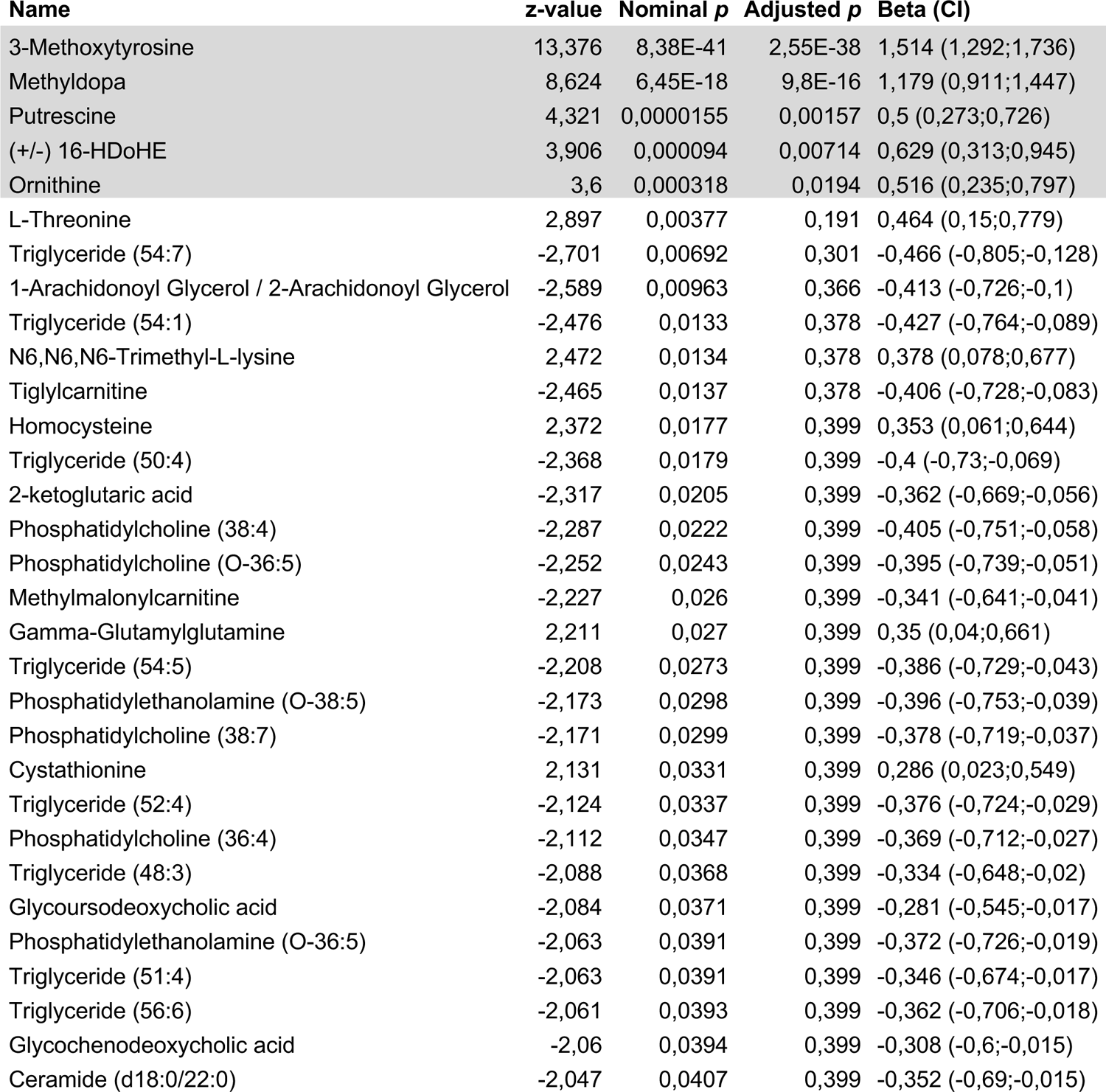
Metabolites differently expressed in IPD patients (n = 140) compared to healthy controls (HC; n = 64). Shown are the results of the fixed effect meta-analysis with a nominal p-value below *p* < 0.05: regression coefficient beta (CI: 95% confidence interval), nominal and adjusted p-values. Metabolites with an adjusted *p* < 0.05 are displayed with a grey background. The complete table with all metabolites, can be found as Supplementary Table 4.

| Name | z-value | Nominal p | Adjusted p | Beta (CI) |
| --- | --- | --- | --- | --- |
| 3-Methoxytyrosine | 13,376 | 8,38E-41 | 2,55E-38 | 1,514 (1,292;1,736) |
| Methyldopa | 8,624 | 6,45E-18 | 9,8E-16 | 1,179 (0,911;1,447) |
| Putrescine | 4,321 | 0,0000155 | 0,00157 | 0,5 (0,273;0,726) |
| (+/-) 16-HDoHE | 3,906 | 0,000094 | 0,00714 | 0,629 (0,313;0,945) |
| Ornithine | 3,6 | 0,000318 | 0,0194 | 0,516 (0,235;0,797) |
| L-Threonine | 2,897 | 0,00377 | 0,191 | 0,464 (0,15;0,779) |
| Triglyceride (54:7) | -2,701 | 0,00692 | 0,301 | -0,466 (-0,805;-0,128) |
| 1-Arachidonoyl Glycerol / 2-Arachidonoyl Glycerol | -2,589 | 0,00963 | 0,366 | -0,413 (-0,726;-0,1) |
| Triglyceride (54:1) | -2,476 | 0,0133 | 0,378 | -0,427 (-0,764;-0,089) |
| N6,N6,N6-Trimethyl-L-lysine | 2,472 | 0,0134 | 0,378 | 0,378 (0,078;0,677) |
| Tiglylcarnitine | -2,465 | 0,0137 | 0,378 | -0,406 (-0,728;-0,083) |
| Homocysteine | 2,372 | 0,0177 | 0,399 | 0,353 (0,061;0,644) |
| Triglyceride (50:4) | -2,368 | 0,0179 | 0,399 | -0,4 (-0,73;-0,069) |
| 2-ketoglutaric acid | -2,317 | 0,0205 | 0,399 | -0,362 (-0,669;-0,056) |
| Phosphatidylcholine (38:4) | -2,287 | 0,0222 | 0,399 | -0,405 (-0,751;-0,058) |
| Phosphatidylcholine (O-36:5) | -2,252 | 0,0243 | 0,399 | -0,395 (-0,739;-0,051) |
| Methylmalonylcarnitine | -2,227 | 0,026 | 0,399 | -0,341 (-0,641;-0,041) |
| Gamma-Glutamylglutamine | 2,211 | 0,027 | 0,399 | 0,35 (0,04;0,661) |
| Triglyceride (54:5) | -2,208 | 0,0273 | 0,399 | -0,386 (-0,729;-0,043) |
| Phosphatidylethanolamine (O-38:5) | -2,173 | 0,0298 | 0,399 | -0,396 (-0,753;-0,039) |
| Phosphatidylcholine (38:7) | -2,171 | 0,0299 | 0,399 | -0,378 (-0,719;-0,037) |
| Cystathionine | 2,131 | 0,0331 | 0,399 | 0,286 (0,023;0,549) |
| Triglyceride (52:4) | -2,124 | 0,0337 | 0,399 | -0,376 (-0,724;-0,029) |
| Phosphatidylcholine (36:4) | -2,112 | 0,0347 | 0,399 | -0,369 (-0,712;-0,027) |
| Triglyceride (48:3) | -2,088 | 0,0368 | 0,399 | -0,334 (-0,648;-0,02) |
| Glycoursodeoxycholic acid | -2,084 | 0,0371 | 0,399 | -0,281 (-0,545;-0,017) |
| Phosphatidylethanolamine (O-36:5) | -2,063 | 0,0391 | 0,399 | -0,372 (-0,726;-0,019) |
| Triglyceride (51:4) | -2,063 | 0,0391 | 0,399 | -0,346 (-0,674;-0,017) |
| Triglyceride (56:6) | -2,061 | 0,0393 | 0,399 | -0,362 (-0,706;-0,018) |
| Glycochenodeoxycholic acid | -2,06 | 0,0394 | 0,399 | -0,308 (-0,6;-0,015) |
| Ceramide (d18:0/22:0) | -2,047 | 0,0407 | 0,399 | -0,352 (-0,69;-0,015) |

Investigating the L-Dopa^positive^ group separately, we identified the same five metabolites already mentioned above, i.e., Methoxytyramine, Methyldopa, 16-HDoHE, Ornithine, and Putrescine, to be significantly elevated patients compared to HC after adjusting for multiple testing (**Figure 3, Table 3, Supplementary Table 5**). As expected, the L-Dopa downstream metabolites^25^ Methoxytyramine and Methyldopa (p < .0001) were highly elevated. Despite also accounting for dopaminergic treatment other than L-Dopa, the LEDD was associated with the levels of both metabolites in the L-Dopa^positive^ group (Methoxythyramine: beta = 0.55 [CI: 0.42 - 0.66, p < .0001]; Methyldopa 0.39 [CI: 0.24 - 0.52, p < .0001]).

**Figure 3:**
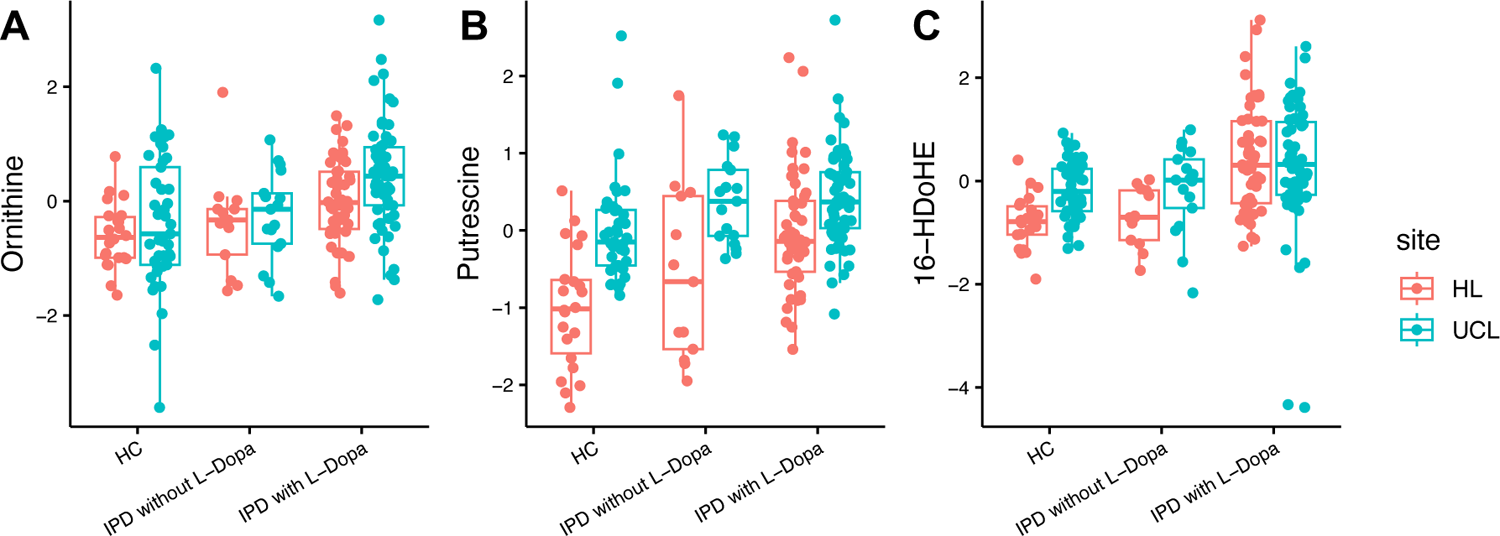
(+/-)-16-HDoHE (A), ornithine (B), and putrescine (C) are increased in idiopathic Parkinsons’s disease (IPD) patients treated with L-Dopa (L-Dopa^positive^) but not elevated in untreated patients and patients with agonist treatment only (L-Dopa^negative^). HC = healthy controls; Data were analyzed as described in the statistic section. No significance levels are depicted in the figure, as data uncorrected for site effects is shown, while statistical calculations were performed by meta-analysis as described in the methods part.

**Table 3.**
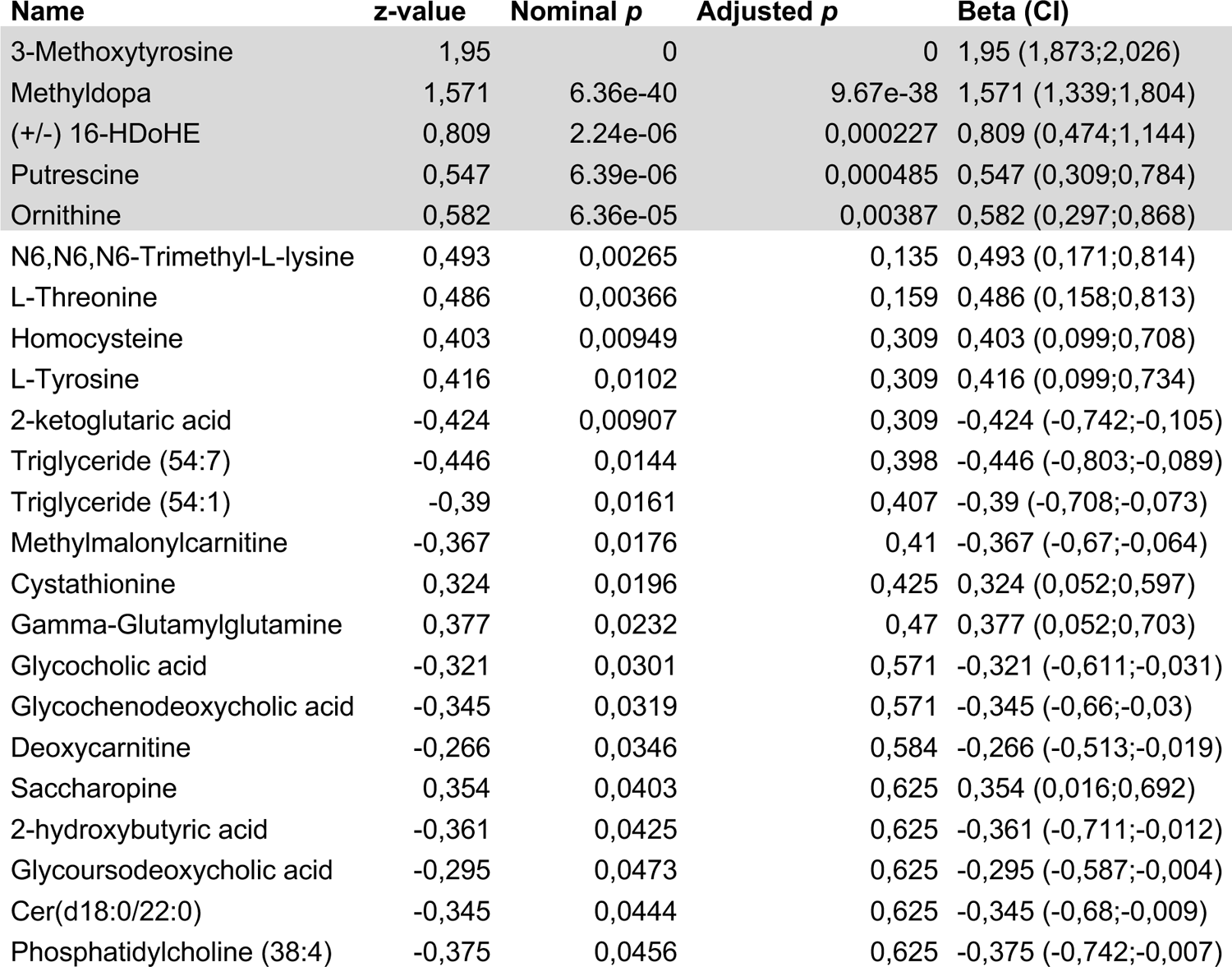
Metabolites differently expressed in IPD patients treated with L-Dopa (Dopa^positive^; n = 110) compared to healthy controls (HC; n = 64). Shown are the results of the fixed effect meta-analysis with a nominal p-value below *p* < 0.05: regression coefficient beta (CI: 95% confidence interval), nominal and adjusted p-values. Metabolites with an adjusted *p* < 0.05 are displayed with a grey background. The complete table with all metabolites can be found as Supplementary Table 5.

### Differences between L-Dopa^negative^ IPD patients and HCs

When comparing L-Dopa^negative^ IPD patients to HC there were no significant differences in metabolite concentration if only adjusted p-values were considered. Of note, the most strikingly elevated metabolites in L-Dopa^positive^ individuals, i.e. Methoxytyramine and Methyldopa, were not at all increased in L-Dopa^negative^ patients compared to controls (**Figure 2**). Regarding 16-HDoHE, Ornithine, and Putrescine there was no difference in L-Dopa^negative^ patients compared to HC after adjusting for multiple testing. However, Putrescine displayed a nominal p-value of 0.026. (adjusted p = 0.56, beta: 0.386; CI [0.046-0.725]) (**Figure 3**).

Among the top hits with nominal p-values < 0.05 in L-Dopa^negative^ IPD patients, which did not show up in L-Dopa^positive^ individuals, we found several membrane lipids such as lysophosphatidic acids (16:0, 14:0), phosphatidylcholines (36:4, 38:7 and 0-36:5), and phosphoethanolamines (34:2, 36:4). Furthermore, we found the endocannabinoids Arachidonoyl glycerol (1-AG/2-AG) and Linoleoyl glycerol (1-LG/2-LG) to be lowered in L-Dopa^negative^ IPD patients. Of note, our methods did not allow us to distinguish between 1-AG and 2-AG and 1-LG and 2-LG respectively. In addition, we found triglycerides (50:1, 52:1, 54:7, and 56:6), Glutathione, Citric acid, and Thromboxane B2 to be elevated. Three acetyl-carnitines were elevated (Hexadecenoylcarnitine, Oleoylcarnitine, Tetradecenoylcarnitine), whereas one was lowered (Tiglylcarnitine) (**Table 4**; **Supplementary Table 6**).

**Table 4.**
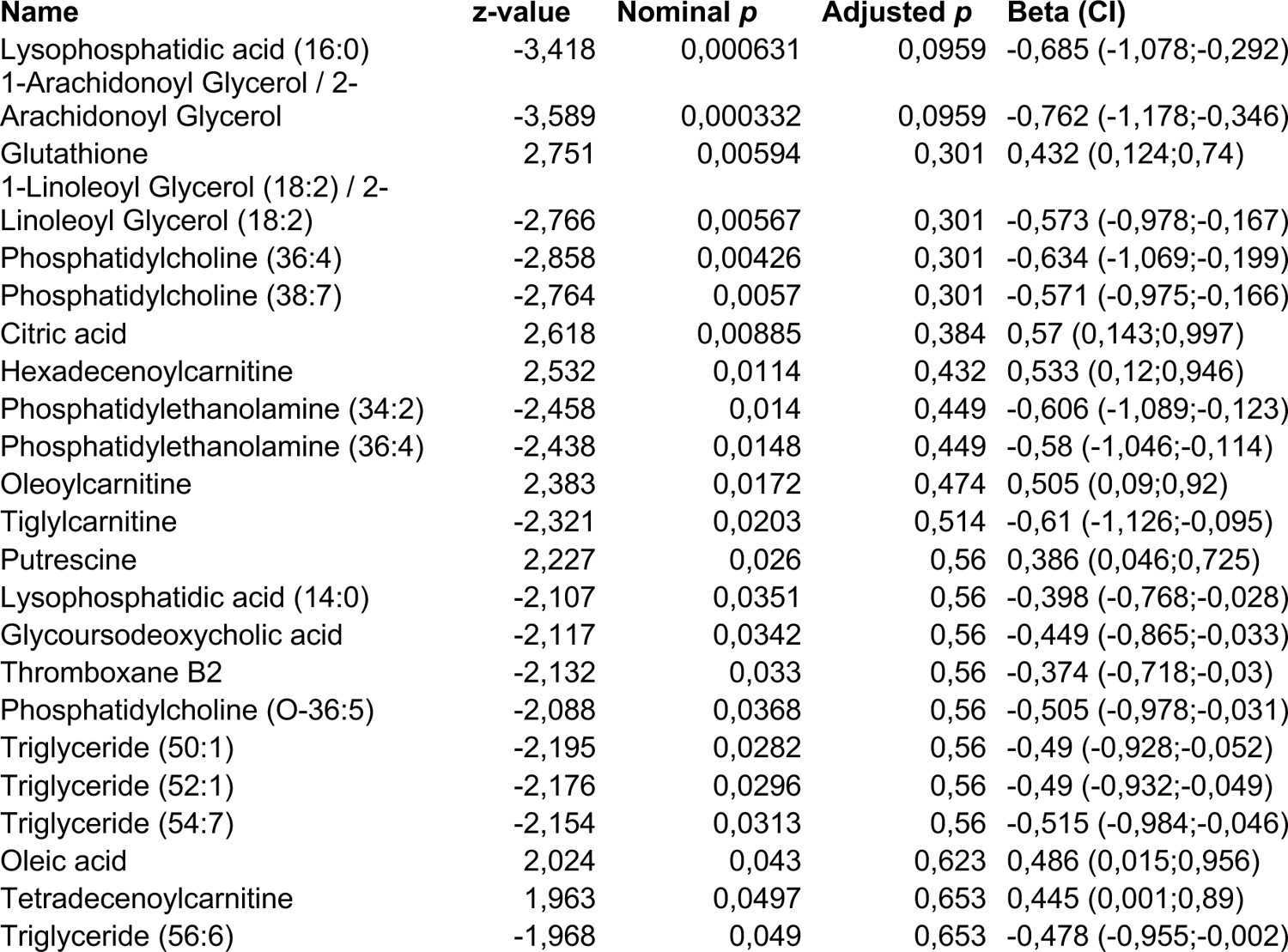
Metabolites differently expressed in IPD patients without L-Dopa treatment (L-Dopa^negative^; n = 30) compared to healthy controls (HC; n = 64). Shown are the results of the fixed effect meta-analysis with a nominal p-value below 0.05, regression coefficient (CI: 95% confidence interval), nominal and adjusted p-values, sorted by adjusted p-value. The complete table with all metabolites can be found as Supplementary Table 6.

| Name | z-value | Nominal p | Adjusted p | Beta (CI) |
| --- | --- | --- | --- | --- |
| Lysophosphatidic acid (16:0) | -3,418 | 0,000631 | 0,0959 | -0,685 (-1,078;-0,292) |
| 1-Arachidonoyl Glycerol / 2-Arachidonoyl Glycerol | -3,589 | 0,000332 | 0,0959 | -0,762 (-1,178;-0,346) |
| Glutathione | 2,751 | 0,00594 | 0,301 | 0,432 (0,124;0,74) |
| 1-Linoleoyl Glycerol (18:2) / 2-Linoleoyl Glycerol (18:2) | -2,766 | 0,00567 | 0,301 | -0,573 (-0,978;-0,167) |
| Phosphatidylcholine (36:4) | -2,858 | 0,00426 | 0,301 | -0,634 (-1,069;-0,199) |
| Phosphatidylcholine (38:7) | -2,764 | 0,0057 | 0,301 | -0,571 (-0,975;-0,166) |
| Citric acid | 2,618 | 0,00885 | 0,384 | 0,57 (0,143;0,997) |
| Hexadecenoylcarnitine | 2,532 | 0,0114 | 0,432 | 0,533 (0,12;0,946) |
| Phosphatidylethanolamine (34:2) | -2,458 | 0,014 | 0,449 | -0,606 (-1,089;-0,123) |
| Phosphatidylethanolamine (36:4) | -2,438 | 0,0148 | 0,449 | -0,58 (-1,046;-0,114) |
| Oleoylcarnitine | 2,383 | 0,0172 | 0,474 | 0,505 (0,09;0,92) |
| Tiglylcarnitine | -2,321 | 0,0203 | 0,514 | -0,61 (-1,126;-0,095) |
| Putrescine | 2,227 | 0,026 | 0,56 | 0,386 (0,046;0,725) |
| Lysophosphatidic acid (14:0) | -2,107 | 0,0351 | 0,56 | -0,398 (-0,768;-0,028) |
| Glycoursodeoxycholic acid | -2,117 | 0,0342 | 0,56 | -0,449 (-0,865;-0,033) |
| Thromboxane B2 | -2,132 | 0,033 | 0,56 | -0,374 (-0,718;-0,03) |
| Phosphatidylcholine (O-36:5) | -2,088 | 0,0368 | 0,56 | -0,505 (-0,978;-0,031) |
| Triglyceride (50:1) | -2,195 | 0,0282 | 0,56 | -0,49 (-0,928;-0,052) |
| Triglyceride (52:1) | -2,176 | 0,0296 | 0,56 | -0,49 (-0,932;-0,049) |
| Triglyceride (54:7) | -2,154 | 0,0313 | 0,56 | -0,515 (-0,984;-0,046) |
| Oleic acid | 2,024 | 0,043 | 0,623 | 0,486 (0,015;0,956) |
| Tetradecenoylcarnitine | 1,963 | 0,0497 | 0,653 | 0,445 (0,001;0,89) |
| Triglyceride (56:6) | -1,968 | 0,049 | 0,653 | -0,478 (-0,955;-0,002) |

### Disease duration and disease severity in L-Dopa^negative^ IPD patients are associated with alterations in different metabolite classes

We conducted regression analyses to investigate whether there was an association between the discovered altered metabolites and disease duration and disease severity in the L-Dopa^negative^ group.

We found higher Putrescine, Glutathione, and Citric acid levels in L-Dopa^negative^ patients compared to HC. Our association analyses revealed a negative correlation of all three metabolites with disease severity but no association with disease duration (**Figure 4**). In contrast, lower levels of endocannabinoids (2-AG and 2-LG) were associated with longer disease duration but had no association with disease severity (**Figure 4**).

**Figure 4.**
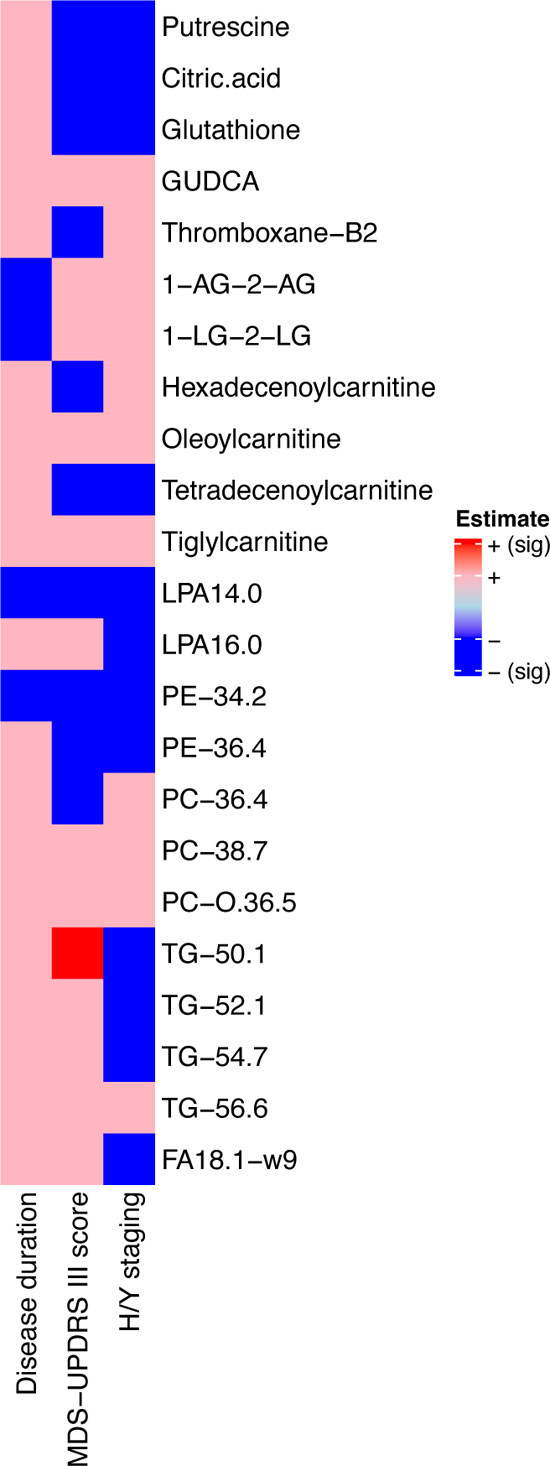
Heatmap of metabolite association with disease duration, and disease severity measured by MDS-UPDRS III and Hoehn und Yahr staging in IPD not treated with L-Dopa (L-Dopa^negative^) patients. Metabolites that displayed a nominal *p* < 0.05 in pairwise comparison between healthy controls and L-Dopa^negative^ patients. Significant influences (*p* < 0.05) are displayed in dark red and blue, trends are marked in light red and blue respectively.

Regarding carnitines, we found an inconclusive picture, with an observed negative correlation between Hexadoconylcarnitine and Tetradecenoylcarnitine with MDS-UPDRS III score, and Tetradecenoylcarnitine with H/Y score. Otherwise, both Oleolylcarnitine and Tigylcarnitine did not show any association with either MDS-UPDRS III or H/Y scores (**Figure 4**).

With respect to lipids, only one of three phosphatidylcholines showed a negative correlation with the UPDRS III score and all phosphatidylcholines did not show an association with disease duration. Lysophosphatidic acid 16:0, the top nominal p-value metabolite, was significantly associated with higher MDS-UPDRS III score but not H/Y stage or disease duration. In contrast, Lysophosphatidic acid 14:0 was negatively correlated with disease duration and severity (**Figure 4**). Both phosphatidylethanolamines negatively correlated with disease severity but only Phosphatidylethanolamine 34.2 was negatively correlated with disease duration. Three triglycerides did not correlate with disease duration or the MDS-UPDRS III score, but one showed a positive correlation. Three out of four negatively correlated with H/Y stage (**Figure 4**). There were no associations between levels of the bile acid Glycoursodeoxycholic acid and disease duration or severity.

*In the mitoPD group, hydroxyeicosatetraenoic acids (HETEs) are elevated which were not found to be altered in IPD patients besides likewise increased L-Dopa-dependent metabolites* All mitoPD patients were treated with L-Dopa, thus investigating a L-Dopa^negative^ group was not possible. Again, the L-Dopa-dependent metabolites Methyldopa (beta 1,95 [CI 1,873 - 2,026]; p < .0001) and Methoxytyramine (beta 1,571 [1,339 - 1,804]; p < .0001) were elevated in the mitoPD group compared to HC after adjusting for multiple testing. Within metabolites with nominal p-values < 0.05, we found among the top hits 16-HDoHE, L-Tyrosine, Putrescine, and Ornithine, but also 5-HETE, 8-HETE, 11-HETE, and 15(S)-HETrE (**Supplementary Table 7**). Correlation analyses in mitoPD patients were not performed due to the small number of patients.

## Discussion

Our study demonstrates (i) that L-Dopa but not agonist treatment significantly impacts the metabolomic profile in PD and (ii) supports a role of specific lipids and endocannabinoids independent from dopaminergic therapy to differentiate between PD patients and HC. Our approach of differentiating between L-Dopa and agonist treatment allows new insights into the PD metabolome, implicating previous studies’ interpretation and future metabolomics study’ design in PD.

### L-Dopa treatment

Many previous PD metabolomic studies have highlighted several metabolites as promising candidates for differentiating HC from PD patients. However, by differentiating between L-Dopa and agonist treatment/no dopaminergic medication, we found several of these metabolites were increased by L-Dopa treatment itself, but not by agonist treatment.

Thus, we have addressed a major challenge faced in studies of the PD metabolism, in that the majority of patients included are treated with L-Dopa. Not surprisingly, Methyldopa and Methoxytyramine, both breakdown products of L-Dopa, were highly elevated in L-Dopa-treated IPD and mitoPD patients. They were neither elevated in IPD patients without L-Dopa treatment nor in patients with L-Dopa agonist treatment only. As expected, the LEDD correlated with levels of Methyldopa and Methoxytyramine in the blood.

In addition, we found 16-HDoHE, a very long-chain fatty acid, to be significantly elevated in L-Dopa-treated patients. 16-HDoHE is a type of hydroxyeicosatetraenoic acid (HETE) that is derived from arachidonic acid.^26^ Both arachidonic acid and docosahexaenoic acids are polyunsaturated fatty acids present in phospholipids of membranes of the body’s cells, and are abundant in the brain. HETEs are markers of oxidative damage and are increased in IPD.^27^ α-syn has been reported to immediately change its structure in the presence of both arachidonic acid and docosahexaenoic acids to its α-helical conformation. Upon prolonged exposure to docosahexaenoic acids, α-syn gradually assembles into amyloid-like fibrils, with the docosahexaenoic acid being part of the aggregate.^28,8^

However, 16-HDoHE did not cluster with its downstream products or other arachidonic acid derivatives like other hydroxydocosahexaenoic acids in our study. It had similar levels in the plasma of non-L-dopa-treated participants and HC. Therefore, the role of 16-HDoHE in the PD metabolome is unclear based on our data. Otherwise, HETEs were among the top hits of altered metabolites investigating mitoPD patients, which are, however, entirely treated with L-Dopa, potentially supporting the role of L-dopa treatment on this specific metabolite pathway.

### Polyamine metabolism

Alterations in the ornithine metabolism, its downstream metabolite Urea,^29^ and polyamine-metabolites such as Putrescine and Spermidine have been repeatedly suggested as a marker of PD state and severity,^30,31^ even when controlling for LEDD.^32^ However, only one of the studies^31^ did include a small group (n=7) of non-L-Dopa treated probands. In our study, Ornithine and Putrescine were also elevated in the L-Dopa^positive^ group. Regarding non-L-Dopa treated individuals, only Putrescine was elevated when compared with HC, and only when considering non-adjusted p-values. Thus, our data implies that polyamine levels in PD are strongly influenced by L-Dopa treatment. In keeping with this suggested crosslink between L-Dopa-treatment and urea metabolism, Putrescine is significantly elevated in rat brains and livers after oral administration of L-Dopa.^33^ We revealed a negative correlation between Putrescine levels and disease duration and severity (MDS-UPDRS III and H/Y stage) in L-Dopa^negative^ patients. This finding could indicate that the disease and the L-Dopa treatment influence the polyamine metabolism. It is tempting to speculate that there is some compensatory upregulation of the polyamine metabolism at the beginning of the disease that decreases in advanced stages. However, there was no association with disease duration. Therefore, the biological underpinnings of a potential relationship between L-Dopa-treatment, Parkinson’s disease and polyamine metabolism needs further investigation.

### Endocannabinoid metabolism

2-AG is the brain’s most abundant endocannabinoid and the primary ligand to the cannabinoid type 1 (CB1) receptor. Another prominent cannabinoid is 2-LG, a partial agonist to the CB1 receptor.^34^ Both regulate diverse neural functions and are fundamental to synaptic plasticity. Endocannabinoids are released in the synapse via a synuclein-dependent mechanism, which is not functional in conditions with misfolded α-syn.^35^ Homozygous loss-of-function mutations in 2-AG synthase diacylglycerol lipase β that produce 2-AG and 2-LG, have been linked to early onset autosomal recessive parkinsonism^36^. In addition, increasing 2-AG levels is neuroprotective in the MPTP-mouse model of PD.^37^

In agreement with these findings and another small study that measured only 2-AG,^38^ we found markedly reduced levels of 2-AG and 2-LG in L-Dopa^negative^ patients. Also, longer disease duration was associated with lower levels of both endocannabinoids in L-Dopa^negative^ patients, suggesting a decline throughout the disease. In mitoPD patients, endocannabinoid levels were not reduced, which could indicate a less severe disruption of the endocannabinoid system, which might be because there is less α-syn involved. The treatment of PD patients with medical cannabis has so far yielded controversial results and needs further investigation with endocannabinoids as potential biomarkers.^39^

### Lipid metabolism and crosslink between fatty acids and alpha-synuclein

Many studies have indicated that fatty acid metabolism is altered in IPD patients, and several membrane-lipids such as sphingolipids and glycerophospholipids^11^ have been consistently reported to be decreased in PD.^2,8^ Of note, several genes in the metabolomic pathway of these lipids are associated with Parkinson’s disease (PD), such as *PLA2G6*, *SMPD1,* and *GBA*.^11^ There is also a well-established interaction between α-syn and Docosahexaenoic acids, as discussed above.^8^ α-syn also interacts with other lipids, such as sphingolipids, and its pathway follows the neurotoxic process after the aggregation of α-syn.^40^

We found several lipids decreased in our study comparing L-Dopa^negative^ with HC. Most of these metabolites belonged to membrane-associated lipids such as sphingolipids, glycerophospholipids, and triglycerides. In detail, we identified lower levels of Lysophosphatidic acid 14:0 and 16:0 that are upstream metabolites forming various other glycerophospholipids and three phosphatidylcholines (PC 36.4, PC O36.5, PC 38.7). PLA2G6 converts lysophosphatidylcholines into phosphatidylcholines, and lipid analysis of brain tissues has revealed that the acyl-chain length of phospholipids is shortened by *PLA2G6* loss, which causes endoplasmic reticulum stress through membrane lipid disequilibrium that in turn leads to dopaminergic neurodegeneration.^41^ Lysophosphatidylcholines and lysophosphatidylethanolamine have been found to strongly inhibit α-syn aggregation.^42^ We also identified lower levels in two phosphatidylethanolamines (34:2 and 36:4), which are downstream metabolites of phosphatidylcholines, which confirms previous findings.^11^

We detected several triglycerides to be lowered in L-Dopa^negative^ patients, and higher H/Y stages were associated with lower levels of triglycerides. The relationship between triglycerides and PD is controversial, as a recent meta-analysis claimed a protective effect of elevated levels of triglycerides in PD.^43^ In contrast another meta-analysis in the same year found no effect.^44^ Because the number of complex lipids is lowered, as discussed above, low levels of simple lipids such as triglycerides could also result from transformation into complex lipids.

Together, the evidence provided by our data, demonstrating alterations in a cluster of lipids acting in a common pathway, excluding the influence of L-dopa-treatment, underscores the potential pathophysiological role of these metabolites in PD, mediated by a possible crosslink with α-syn.

### Other metabolites

Acylcarnitines transport acyl-groups (organic acids and fatty acids) from the cytoplasm into the mitochondria, where they are broken down to produce energy via beta-oxidation. We found three acylcarnitines to be increased (Hexadecenoylcarnitine, Tetradecenoylcarnitine, and Oleoylcarnitine) and one (Tiglylcarnitine) to be lowered in L-Dopa^negative^ patients. Previous studies had suggested lower levels of free and total carnitine levels in older individuals^3^ and in healthy individuals compared to IPD patients. However, no drug naïve patients had been included.^45,46^ Our analyses revealed no clear correlation of acylcarnitines with disease severity and duration.

Furthermore, we found Glutathione, the most abundant and significant antioxidant in the human body, to be increased in L-Dopa^negative^ patients. This finding contrasts with previous research that found decreased levels of Glutathione in the substantia nigra of IPD patients.^47^ A recent study suggested that an increase in plasma Glutathione was associated with less increase in PD probability,^48^ which aligns with our finding of a negative correlation between glutathione levels and clinical severity. However, clinical trials had not shown glutathione treatment to be superior to placebo regarding the alleviation of PD symptoms.^49,50^

We found reduced Glycoursodeoxycholic acid levels in L-Dopa^negative^ patients by nominal p-values. While Glycoursodeoxycholic acid has not yet been linked to PD, it was found to be increased in patients with type 2 diabetes and has been discussed as a marker for hyperglycemia.^51^ Given the high prevalence of type 2 diabetes among individuals with PD,^52^ we consider this discovery highly credible.

Finally, Citric acid was found to be elevated in L-Dopa^negative^ patients by nominal p-value. No study has yet suggested Citric acid to be elevated in PD, but we found a strong connection of its elevation with age at examination, that has widely been reported. We also found a negative correlation with disease severity that could support a protective role of higher Citric acid levels. Accumulation of Citric acid, the starting point of the citric acid circle, could also indicate a shift from oxidative phosphorylation to increased glycolysis due to mitochondrial dysfunction, which we previously described.^53,54^

### Mito-PD

Only one study investigated the metabolome in *PRKN*-linked PD^55^ and no investigations addressing patients with *PINK1*-linked PD have been available thus far. We combined these two autosomal recessively inherited forms of PD, as the proteins encoded by these genes act in a common biochemical pathway, mainly involved in the degradation of damaged mitochondria,^6^ but also links to alterations of innate immune system have been described to be associated with Parkin and Pink1 dysfunction.^56^ Importantly, we were able to partly reproduce the differences described in the only other metabolomics study investigating *PRKN*-linked PD patients, where an elevation of fatty acids and oxidized lipids and a decrease of antioxidants, caffeine, and benzoate-related metabolites were reported. We found several hydroxyeicosatetraenoic acids (5-HETE, 8-HETE, 11-HETE, and 15(S)-HETrE) elevated at nominal p-values. These metabolites were not found to be different in the comparison between IPD and HC. As explained above, this might reflect different pathways of oxidative dysfunction in these subsets of PD patients known to display mitochondrial alterations.

As all *PRKN/PINK1*-linked PD patients in this study were treated with L-Dopa, conclusions drawn from these analyses are limited due to the significant effect of L-Dopa treatment on the metabolome. Methoxytyrosine and Methyldopa were the only metabolites that were significantly different between the mitoPD group and HC. Furthermore, most metabolites with nominal p-values < 0.05, such as (+/-) 16-HDoHE, L-Tyrosine, Putrescine, and Ornithine, were identified as L-Dopa-dependent within our study. Together, investigating the mitoPD group did not allow us to draw definite conclusions, as all patients were treated with L-Dopa, and the sample size was too small.

### Strengths and Limitations

The strengths of our study consist of considering the substantial effect of L-Dopa medication and investigating two independent samples of probands recruited with the same detailed protocol to ensure identical pre-analytical handling. Nonetheless, we observed site-dependent effects. Thus, we applied a strict statistical approach to correct for this bias, as well as for multiple testing. Furthermore, we used state-of-the-art large-scale comprehensive metabolomic platforms to measure over 300 metabolites and identify differences in single metabolites rather than in clusters.

Limitations include the cross-sectional study design and the relatively small sample size regarding patients with *PRKN-/PINK1*-linked disease, making correlations with disease severity impossible. Moreover, no group of patients with pathogenic biallelic variants in *PRKN* and *PINK1* without L-Dopa treatment was available, preventing deeper insight into the effects of dopaminergic medication on the metabolome in this group of PD patients.

## Conclusions

We investigated a broad range of more than 300 metabolites that covered all major biochemical pathways. Adjusting for multiple testing, only metabolites impacted by L-dopa treatment significantly differed, highlighting the dominant effect of L-dopa on the PD metabolome. However, several metabolites had nominal p-values < .005 comparing HCs and non-L-dopa treated PD patients, which we consider relevant as (i) applying strict statistical correction for multiple testing might mitigate all but significant effects if such a magnitude of tests are performed, (ii) several of these metabolites have been consistently reproduced in other studies as we recently reviewed,^2^ (iii) several of these metabolites are belonged to the same biological pathways, making a role in PD metabolism more plausible, and (iv) the relevant metabolites were associated with disease state and progression, further highlighting their biological impact.

Overall, our study underscores the large impact of L-Dopa treatment, on metabolomic studies in PD. Thus we highlight the risk of misinterpreting differences between IPD patients and healthy controls as disease-specific, when in fact they may relate simply to L-Dopa treatment. Thus, the impact of dopaminergic treatment should be considered in all future metabolomics studies. This might also be the case for biomarker studies dealing with PD in general, as with recent findings implicate that L-Dopa treatment even impacts PD imaging biomarkers.^57^ Furthermore, our associative analyses revealed the evidence of different metabolite-trajectories throughout the disease; however, longitudinal metabolomic studies are warranted to assess relevant metabolite changes over the disease course.

Finally, we found evidence for the role of specific glycerophospholipids and endocannabinoids in disease progression and severity, supporting recent findings that lipid pathway alterations influence PD pathogenesis.

## Ethics and Consent

The study was approved by the ethics committees at the University of Lübeck, Lübeck, Germany and University College London, London, United Kingdom. All participants gave written informed consent.

## Data Availability

All results after statistical analyses are available within the article and its supplementary material. The raw data that support the findings of this study are available from the corresponding author, upon reasonable request.

