## Supplement for "The role of dopaminergic medication, lipid, and endocannabinoid pathway alterations in idiopathic and *PRKN/PINK1*-mediated Parkinson’s disease – a large-scale targeted metabolomics study"

*contributed equally

^#^Corresponding authors:

Supplementary Methods

| **Analysis** | **Method** | **Dependent variable and covariates** | **Individuals and metabolites** |
| --- | --- | --- | --- |
| Site differences | Linear regression for each metabolite separately  Adjustment of P-values using Benjamini-Hochberg approach | Dependent variable: Transformed metabolite intensity Independent variables: age at examination, sex, Diagnosis.group | 204 controls and IPD patients  304 metabolites |
| Association with disease status | For each metabolite and pairwise comparison:  Analysis separately for each site: - linear regression  Fixed effect meta-analysis and adjustment of P-values using Benjamini-Hochberg approach | Dependent variable: Transformed metabolite intensity, ratios or sums  Independent variables: Diagnosis.group, Sex, AAE | 64 controls  140 IPD patients  304 metabolites |
| Comparison control vs IPD patients with and without Levodopa | For each metabolite and pairwise comparison:  Analysis separately for each site:  - linear regression  Fixed effect meta-analysis and adjustment of P-values using Benjamini-Hochberg approach | Dependent variable: Transformed metabolite intensity, ratios or sums  Independent variables: treatment_group, sex, age at examination | 30 No levodopa  110 Levodopa  64 HC  304 metabolites |
| Association with age (In cases and controls) | For each metabolite analysis separately for each site:  Linear regression  Fixed effect meta-analysis of interaction term and adjustment of P-values using Benjamini-Hochberg approach | Dependent variable: Transformed metabolite intensity, ratios or sums  Independent variables: Age at examination, Sex, Diagnosis.group | 204 controls and IPD patients  304 metabolites |
| Association with UPDRS III total score (IPD patients without Levodopa) | For each metabolite and pairwise comparison:  Analysis separately for each site: Linear regression  Fixed effect meta-analysis and adjustment of P-values using Benjamini-Hochberg approach | Dependent variable: Transformed metabolite intensity, ratios or sums  Independent variables: MDS-UPDRS III Total Score, Sex | 29 IPD patients without Levodopa  304 metabolites |
| Association with disease duration (IPD patients without Levodopa) | For each metabolite and pairwise comparison:  Analysis separately for each site: Linear regression  Fixed effect meta-analysis and adjustment of P-values using Benjamini-Hochberg approach | Dependent variable: Transformed metabolite intensity, ratios or sums  Independent variables: Disease Duration, Sex, age at examination | 29 IPD patients without Levodopa  304 metabolites |
| Association with Hoehn and Yahr staging (IPD patients without Levodopa) | For each metabolite and pairwise comparison:  Analysis separately for each site:  Linear regression  Fixed effect meta-analysis and adjustment of P-values using Benjamini-Hochberg approach | Dependent variable: Transformed metabolite intensity, ratios or sums  Independent variables: HoehnandYahrStaging, Sex. | 29 IPD patients without Levodopa  304 metabolites |

Supplementary Tables

**Supplementary Table 1. Overview over all measured metabolites (n= 304) and their respective identifiers.** Metabolites are ordered alphabetically, NA: not avaliable

| **Metabolite Name** | **ChEBI ID** | **PubChem_ID** | **Lipidmaps** |
| --- | --- | --- | --- |
| (+/-) 10-HDoHE | 72640 | 11537494 | LMFA04000027 |
| (+/-) 11-HDoHE | 72794 | 11631564 | LMFA04000028 |
| (+/-) 14-HDoHE | 72647 | 11566378 | LMFA04000030 |
| (+/-) 16-HDoHE | 72613 | 11595378 | LMFA04000031 |
| (+/-) 8-HDoHE | 72610 | 11976798 | LMFA04000026 |
| 1-AG/2-AG | 34071;52392 | 16019980;5282280 | LMGL01010032;LMGL01010023 |
| 1-LG/2-LG | 75561;75457 | 5283469;5365676 | LMGL01010006;LMGL01010033 |
| 1-Methylhistidine | 70958 | NA | NA |
| 11-HETE | 72606 | 14123410 | LMFA03060085 |
| 11,12-DiHETrE | 63969 | 5283146 | LMFA03050008 |
| 12-HETE | 8447 | 13786989 | LMFA03060088 |
| 12,13-DiHODE | 88461 | 16061067 | LMFA02000046 |
| 12,13-DiHOME | 72665 | 10236635 | LMFA02000230 |
| 12,13-EpOME | 38229 | 5356421 | LMFA02000038 |
| 12(S)-HEPE | 72645 | 10041593 | LMFA03070031 |
| 13-HODE | 72639 | 5282947 | LMFA02000154 |
| 14,15-DiHETE | 88459 | 16061119 | LMFA03060077 |
| 14,15-DiHETrE | 63966 | 5283147 | LMFA03050010 |
| 15(S)-HETrE | 88348 | 5283145 | LMFA03050007 |
| 17,18-DiHETE | 88349 | 16061120 | LMFA03060078 |
| 19,20-DiHDPA | 72657 | 16061148 | LMFA04000043 |
| 2-ketoglutaric acid | 30915 | NA | NA |
| 2-Methylbutyroylcarnitine | 73026 | NA | NA |
| 3-Hydroxybutyric acid | 20067 | NA | NA |
| 3-Hydroxypropionic Acid | 33404 | NA | NA |
| 3-Methoxytyrosine | 1582 | NA | NA |
| 5-HETE | 28209 | 9862886 | LMFA03060084 |
| 5,6-DiHETrE | 63974 | 5283142 | LMFA03050004 |
| 8-HETE | 72643 | 11976122 | LMFA03060086 |
| 8,12-iPF2a IV | NA | 156593941 | NA |
| 8,9-DiHETrE | 63970 | 5283144 | LMFA03050006 |
| 8(9)-EpETrE | 34490 | 5283203 | LMFA03080003 |
| 9-HODE | 72651 | 5282944 | LMFA02000151 |
| 9-HOTrE | 80447 | 6439873 | LMFA02000024 |
| 9,10-DiHOME | 72663 | 9966640 | LMFA02000229 |
| 9,10,13-TriHOME | 34499 | 5282965 | LMFA02000168 |
| 9,12,13-TriHOME | 34506 | 9858729 | LMFA02000014 |
| Acetylcarnitine | 73024 | NA | NA |
| Anandamide | 2700 | 5281969 | LMFA08040001 |
| Betaine | 41139 | NA | NA |
| Butyrylcarnitine | 7676 | NA | NA |
| Carnitine | 17126 | NA | NA |
| CE(18:1) | NA | NA | NA |
| CE(18:2) | NA | NA | NA |
| CE(20:4) | NA | NA | NA |
| CE(22:6) | NA | NA | NA |
| Cer(d18:0/22:0) | NA | NA | NA |
| Cer(d18:0/23:0) | NA | NA | NA |
| Cer(d18:1/22:0) | NA | NA | NA |
| Cer(d18:1/23:0) | NA | NA | NA |
| Cer(d18:1/24:1) | NA | NA | NA |
| Cholic acid | 16359 | 221493 | LMST04010001 |
| Choline | 15354 | NA | NA |
| Citric acid | 30769 | NA | NA |
| Citrulline | 18211 | NA | NA |
| Cortisol | 17650 | 5754 | LMST02030001 |
| Cystathionine | 15901 | NA | NA |
| Cysteine | 15356 | NA | NA |
| Decanoylcarnitine | 68830 | NA | NA |
| Decenoylcarnitine | 86063 | NA | NA |
| Dehydroepiandrosteron | 85252 | 5283451 | LMFA08040009 |
| Deoxycarnitine | 16244 | NA | NA |
| Deoxycholic acid | 28834 | 222528 | LMST04010040 |
| DG(36:2) | NA | NA | NA |
| DG(36:3) | NA | NA | NA |
| DGLEA | 34488 | 5282272 | LMFA08040011 |
| DL-3-aminoisobutyric acid | 27389 | NA | NA |
| Dodecenoylcarnitine | 86065 | NA | NA |
| Ethanolamine | 16000 | NA | NA |
| FA 18(1) | 16196 | 445639 | LMFA01030002 |
| FA 18(2) | 17351 | 5280450 | LMFA01030120 |
| FA 22(5) w6 | 65136 | 6441454 | LMFA04000064 |
| FA 22(6) | 28125 | 445580 | LMFA01030185 |
| Gamma-aminobutyric acid | 16865 | NA | NA |
| Gamma-Glutamylglutamine | 73707 | NA | NA |
| Gamma-L-glutamyl-L-alanine | 50619 | NA | NA |
| Glutathione | 16856 | NA | NA |
| Glycine | 15428 | NA | NA |
| Glycochenodeoxycholic acid | 36274 | 12544 | LMST05030008 |
| Glycocholic acid | 17687 | 10140 | LMST05030001 |
| Glycodeoxycholic acid | 27471 | 3035026 | LMST05030006 |
| Glycolithocholic acid | 37998 | 115245 | LMST05030009 |
| Glycoursodeoxycholic acid | 89929 | 12310288 | LMST05030016 |
| Glycylglycine | 17201 | NA | NA |
| Hexadecenoylcarnitine | 86032 | NA | NA |
| Hexanoylcarnitine | 70749 | NA | NA |
| Homocitrulline | 17443 | NA | NA |
| Homocysteine | 17230 | NA | NA |
| Hydroxylysine | 86498 | NA | NA |
| Isobutyrylcarnitine | 73017 | NA | NA |
| Isocitrate | 30887 | NA | NA |
| Isovalerylcarnitine | 73025 | NA | NA |
| L-2-aminoadipic acid | 37024 | NA | NA |
| L-4-hydroxy-proline | 20392 | NA | NA |
| L-Alanine | 16449 | NA | NA |
| L-Alpha-aminobutyric acid | 35621 | NA | NA |
| L-Arginine | 29016 | NA | NA |
| L-Asparagine | 22653 | NA | NA |
| L-Glutamic acid | 18237 | NA | NA |
| L-Glutamine | 28300 | NA | NA |
| L-Histidine | 27570 | NA | NA |
| L-Homoserine | 30653 | NA | NA |
| L-Isoleucine | 24898 | NA | NA |
| L-Kynurenine | 28683 | NA | NA |
| L-Leucine | 25017 | NA | NA |
| L-Lysine | 25094 | NA | NA |
| L-Methionine | 16811 | NA | NA |
| L-Phenylalanine | 28044 | NA | NA |
| L-Proline | 26271 | NA | NA |
| L-Serine | 17822 | NA | NA |
| L-Threonine | 26986 | NA | NA |
| L-Tryptophan | 27897 | NA | NA |
| L-Tyrosine | 18186 | NA | NA |
| L-Valine | 27266 | NA | NA |
| Lactic acid | 422 | NA | NA |
| Lauroylcarnitine | 73054 | NA | NA |
| Linoleoyl ethanolamide | 64032 | 5283446 | LMFA08040004 |
| Linoleylcarnitine | 73072 | NA | NA |
| LPS 18(1) | 52649 | 9547099 | LMGP03050001 |
| Lysophosphatidic acid 14(0) | 62833 | 9547180 | LMGP10050007 |
| Lysophosphatidic acid 16(0) | 15799 | 6419701;53478599 | LMGP10050006;LMGP10050042 |
| Lysophosphatidic acid 16(1) | 75070 | 52929751 | LMGP10050016 |
| Lysophosphatidic acid 18(1) | 62837;78270 | 5311263;52929749 | LMGP10050008;LMGP10050014 |
| Lysophosphatidic acid 18(2) | 62834;74330 | 50990923;53478601 | LMGP10050017;LMGP10050044 |
| Lysophosphatidylcholine(14:0) | NA | NA | NA |
| Lysophosphatidylcholine(16:0) | NA | NA | NA |
| Lysophosphatidylcholine(16:1) | NA | NA | NA |
| Lysophosphatidylcholine(18:0) | NA | NA | NA |
| Lysophosphatidylcholine(18:1) | NA | NA | NA |
| Lysophosphatidylcholine(18:2) | NA | NA | NA |
| Lysophosphatidylcholine(18:3) | NA | NA | NA |
| Lysophosphatidylcholine(20:3) | NA | NA | NA |
| Lysophosphatidylcholine(20:4) | NA | NA | NA |
| Lysophosphatidylcholine(20:5) | NA | NA | NA |
| Lysophosphatidylcholine(22:6) | NA | NA | NA |
| Lysophosphatidylcholine(O-16:0) | NA | NA | NA |
| Lysophosphatidylcholine(O-16:1) | NA | NA | NA |
| Lysophosphatidylcholine(O-18:1) | NA | NA | NA |
| Lysophosphatidylethanolamine (16:0) | 73134;131743 | 9547069;53480922 | LMGP02050002;LMGP02050036 |
| Lysophosphatidylethanolamine (16:1) | 145277;145248 | 52925129;53480923 | LMGP02050010;LMGP02050037 |
| Lysophosphatidylethanolamine (20:4) | 64395 | 42607465;53480936;53480952;53480937 | LMGP02050009;LMGP02050051;LMGP02050067;LMGP02050052 |
| Lysophosphatidylethanolamine (20:5) | 145287;145263 | 52925146;53480938 | LMGP02050027;LMGP02050053 |
| Lysophosphatidylethanolamine(18:0) | NA | NA | NA |
| Lysophosphatidylethanolamine(18:1) | NA | NA | NA |
| Lysophosphatidylethanolamine(20:4) | NA | NA | NA |
| Lysophosphatidylethanolamine(22:6) | NA | NA | NA |
| Lysophosphatidylinositol 16(1) | NA | 52928604 | LMGP06050009 |
| Lysophosphatidylinositol 18(0) | 83054 | 42607495 | LMGP06050004 |
| Lysophosphatidylinositol 18(1) | 82753 | 42607496 | LMGP06050005 |
| Lysophosphatidylinositol 18(2) | NA | 52928605 | LMGP06050010 |
| Lysophosphatidylinositol 20(4) | NA | 42607497 | LMGP06050006 |
| Malic acid | 6650 | NA | NA |
| Malonylcarnitine | 73028 | NA | NA |
| Methionine sulfone | 132188 | NA | NA |
| Methyldopa | 167648 | NA | NA |
| Methylmalonylcarnitine | 73031 | NA | NA |
| Myristoilcarnitine | 73061 | NA | NA |
| N6,N6,N6-Trimethyl-L-lysine | 165870 | NA | NA |
| Nonaylcarnitine | 70997 | NA | NA |
| O-Acetyl-L-serine | 17981 | NA | NA |
| O-Anandamide | 418207 | 5712057 | LMFA00000014 |
| O-Phosphoethanolamine | 17553 | NA | NA |
| OA01 - 2-hydroxybutyric acid | 1148 | NA | NA |
| Octanoylcarnitine | 73039 | NA | NA |
| Octenoylcarnitine | 86052 | NA | NA |
| Oleoylcarnitine | 72689 | NA | NA |
| Ornithine | 18257 | NA | NA |
| Palmitoyl ethanolamide | 71464 | 4671 | LMFA08040013 |
| Palmitoylcarnitine | 73067 | NA | NA |
| Phosphatidylcholine(32:0) | NA | NA | NA |
| Phosphatidylcholine(32:1) | NA | NA | NA |
| Phosphatidylcholine(32:2) | NA | NA | NA |
| Phosphatidylcholine(34:1) | NA | NA | NA |
| Phosphatidylcholine(34:2) | NA | NA | NA |
| Phosphatidylcholine(34:3) | NA | NA | NA |
| Phosphatidylcholine(34:4) | NA | NA | NA |
| Phosphatidylcholine(36:1) | NA | NA | NA |
| Phosphatidylcholine(36:2) | NA | NA | NA |
| Phosphatidylcholine(36:3) | NA | NA | NA |
| Phosphatidylcholine(36:4) | NA | NA | NA |
| Phosphatidylcholine(36:5) | NA | NA | NA |
| Phosphatidylcholine(38:2) | NA | NA | NA |
| Phosphatidylcholine(38:3) | NA | NA | NA |
| Phosphatidylcholine(38:4) | NA | NA | NA |
| Phosphatidylcholine(38:5) | NA | NA | NA |
| Phosphatidylcholine(38:6) | NA | NA | NA |
| Phosphatidylcholine(38:7) | NA | NA | NA |
| Phosphatidylcholine(40:5) | NA | NA | NA |
| Phosphatidylcholine(40:6) | NA | NA | NA |
| Phosphatidylcholine(40:7) | NA | NA | NA |
| Phosphatidylcholine(40:8) | NA | NA | NA |
| Phosphatidylcholine(O-34:1) | NA | NA | NA |
| Phosphatidylcholine(O-34:2) | NA | NA | NA |
| Phosphatidylcholine(O-34:3) | NA | NA | NA |
| Phosphatidylcholine(O-36:3) | NA | NA | NA |
| Phosphatidylcholine(O-36:4) | NA | NA | NA |
| Phosphatidylcholine(O-36:5) | NA | NA | NA |
| Phosphatidylcholine(O-38:4) | NA | NA | NA |
| Phosphatidylcholine(O-38:5) | NA | NA | NA |
| Phosphatidylcholine(O-38:6) | NA | NA | NA |
| Phosphatidylcholine(O-44:5) | NA | NA | NA |
| Phosphatidylethanolamine(34:2) | NA | NA | NA |
| Phosphatidylethanolamine(36:4) | NA | NA | NA |
| Phosphatidylethanolamine(38:2) | NA | NA | NA |
| Phosphatidylethanolamine(38:4) | NA | NA | NA |
| Phosphatidylethanolamine(38:6) | NA | NA | NA |
| Phosphatidylethanolamine(O-36:5) | NA | NA | NA |
| Phosphatidylethanolamine(O-38:5) | NA | NA | NA |
| Phosphatidylethanolamine(O-38:7) | NA | NA | NA |
| Pimelylcarnitine | 86084 | NA | NA |
| Propionylcarnitine | 28867 | NA | NA |
| Prostaglandin E2 | 15551 | 5280360 | LMFA03010003 |
| Prostaglandin F2α | 15553 | 5280363 | LMFA03010002 |
| Putrescine | 17148 | NA | NA |
| Pyroglutamic acid | 16010 | NA | NA |
| S-Methylcysteine | 45658 | NA | NA |
| Saccharopine | 30768 | NA | NA |
| Sarcosine | 15611 | NA | NA |
| Serotonine | 28790 | NA | NA |
| Sphingomyelin(d18:1/14:0) | NA | NA | NA |
| Sphingomyelin(d18:1/15:0) | NA | NA | NA |
| Sphingomyelin(d18:1/16:0) | NA | NA | NA |
| Sphingomyelin(d18:1/16:1) | NA | NA | NA |
| Sphingomyelin(d18:1/18:0) | NA | NA | NA |
| Sphingomyelin(d18:1/18:1) | NA | NA | NA |
| Sphingomyelin(d18:1/18:2) | NA | NA | NA |
| Sphingomyelin(d18:1/20:0) | NA | NA | NA |
| Sphingomyelin(d18:1/20:1) | NA | NA | NA |
| Sphingomyelin(d18:1/21:0) | NA | NA | NA |
| Sphingomyelin(d18:1/22:0) | NA | NA | NA |
| Sphingomyelin(d18:1/22:1) | NA | NA | NA |
| Sphingomyelin(d18:1/23:0) | NA | NA | NA |
| Sphingomyelin(d18:1/23:1) | NA | NA | NA |
| Sphingomyelin(d18:1/24:0) | NA | NA | NA |
| Sphingomyelin(d18:1/24:1) | NA | NA | NA |
| Sphingomyelin(d18:1/24:2) | NA | NA | NA |
| Sphingomyelin(d18:1/25:0) | NA | NA | NA |
| Sphingomyelin(d18:1/25:1) | NA | NA | NA |
| Sphingosine 1-phosphate 18(0) | 16893 | 91486 | LMSP01050002 |
| Sphingosine 1-phosphate 18(1) | 37550 | 5283560 | LMSP01050001 |
| Sphingosine 1-phosphate 18(2) | NA | NA | NA |
| Stearoylcarnitine | 73074 | NA | NA |
| Symmetric dimethylarginine | 25682 | NA | NA |
| Taurine | 15891 | NA | NA |
| Taurochenodeoxycholic acid | 16525 | 387316 | LMST05040005 |
| Taurocholic acid | 28865 | 6675 | LMST05040001 |
| Taurodeoxycholic acid | 9410 | 2733768 | LMST05040013 |
| Taurolithocholic acid | 36259 | 439763 | LMST05040003 |
| Tetradecadienylcarnitine | 86069 | NA | NA |
| Tetradecenoylcarnitine | 86066 | NA | NA |
| TG(42:0) | NA | NA | NA |
| TG(42:1) | NA | NA | NA |
| TG(42:2) | NA | NA | NA |
| TG(44:0) | NA | NA | NA |
| TG(44:1) | NA | NA | NA |
| TG(44:2) | NA | NA | NA |
| TG(45:0) | NA | NA | NA |
| TG(46:0) | NA | NA | NA |
| TG(46:1) | NA | NA | NA |
| TG(46:2) | NA | NA | NA |
| TG(48:1) | NA | NA | NA |
| TG(48:2) | NA | NA | NA |
| TG(48:3) | NA | NA | NA |
| TG(50:1) | NA | NA | NA |
| TG(50:2) | NA | NA | NA |
| TG(50:3) | NA | NA | NA |
| TG(50:4) | NA | NA | NA |
| TG(51:1) | NA | NA | NA |
| TG(51:2) | NA | NA | NA |
| TG(51:3) | NA | NA | NA |
| TG(51:4) | NA | NA | NA |
| TG(52:1) | NA | NA | NA |
| TG(52:2) | NA | NA | NA |
| TG(52:3) | NA | NA | NA |
| TG(52:4) | NA | NA | NA |
| TG(53:1) | NA | NA | NA |
| TG(54:0) | NA | NA | NA |
| TG(54:1) | NA | NA | NA |
| TG(54:2) | NA | NA | NA |
| TG(54:3) | NA | NA | NA |
| TG(54:4) | NA | NA | NA |
| TG(54:5) | NA | NA | NA |
| TG(54:7) | NA | NA | NA |
| TG(55:1) | NA | NA | NA |
| TG(56:0) | NA | NA | NA |
| TG(56:1) | NA | NA | NA |
| TG(56:2) | NA | NA | NA |
| TG(56:3) | NA | NA | NA |
| TG(56:4) | NA | NA | NA |
| TG(56:5) | NA | NA | NA |
| TG(56:6) | NA | NA | NA |
| TG(56:7) | NA | NA | NA |
| TG(57:1) | NA | NA | NA |
| TG(58:2) | NA | NA | NA |
| TG(58:8) | NA | NA | NA |
| TG(58:9) | NA | NA | NA |
| TG(59:1) | NA | NA | NA |
| TG(60:1) | NA | NA | NA |
| TG(60:2) | NA | NA | NA |
| TG(60:3) | NA | NA | NA |
| Thromboxane B2 | 28728 | 5283137 | LMFA03030002 |
| Tiglylcarnitine | 71179 | NA | NA |
| Trimethylamine N-oxide | 15724 | NA | NA |
| Valerylcarnitine | 86050 | NA | NA |

**Supplementary Table 2. Influence of the recruitment site on metabolite levels for HC and all IPD patients combined (n = 204).** Shown are the results of a linear regression for each metabolite and site separately: regression coefficient (beta), standard error (SE), nominal and adjusted p-values.

| **Name** | **Mean all** | **Mean UL** | **Mean UCL** | **Beta** | **SE** | **Nominal *p*** | **Adjusted *p*** |
| --- | --- | --- | --- | --- | --- | --- | --- |
| Sphingosine 1-phosphate 18(1) | -0,045 | -1,071 | 0,689 | 1,731 | 0,077 | 2.22e-56 | 6.74e-54 |
| Sphingosine 1-phosphate 18(0) | -0,05 | -1,077 | 0,683 | 1,752 | 0,079 | 1.33e-55 | 2.02e-53 |
| 12-HETE | -0,044 | -1,079 | 0,696 | 1,728 | 0,08 | 7.17e-54 | 7.27e-52 |
| 12(S)-HEPE | -0,03 | -1,06 | 0,706 | 1,758 | 0,082 | 2.88e-53 | 2.19e-51 |
| (+/-) 14-HDoHE | -0,022 | -1,049 | 0,711 | 1,759 | 0,083 | 5.59e-53 | 3.4e-51 |
| (+/-) 10-HDoHE | -0,017 | -1,017 | 0,697 | 1,735 | 0,087 | 4.06e-49 | 2.06e-47 |
| O-Phosphoethanolamine | -0,039 | -1,023 | 0,673 | 1,652 | 0,087 | 5.09e-46 | 2.21e-44 |
| Taurine | -0,037 | -0,989 | 0,652 | 1,605 | 0,092 | 7.85e-42 | 2.98e-40 |
| (+/-) 11-HDoHE | -0,015 | -0,989 | 0,681 | 1,68 | 0,098 | 5.84e-41 | 1.97e-39 |
| Sphingosine 1-phosphate 18(2) | -0,051 | -0,945 | 0,587 | 1,475 | 0,089 | 1.59e-39 | 4.82e-38 |
| LPS 18(1) | -0,046 | -0,977 | 0,619 | 1,548 | 0,097 | 1.36e-37 | 3.75e-36 |
| 8(9)-EpETrE | -0,045 | -0,93 | 0,587 | 1,502 | 0,101 | 5.52e-34 | 1.4e-32 |
| Serotonine | -0,025 | -0,893 | 0,603 | 1,456 | 0,107 | 4.16e-30 | 9.72e-29 |
| 8-HETE | -0,079 | -0,865 | 0,483 | 1,267 | 0,107 | 7.83e-25 | 1.7e-23 |
| Isocitrate | -0,042 | -0,696 | 0,424 | 1,268 | 0,116 | 5.9e-22 | 1.2e-20 |
| 15(S)-HETrE | -0,068 | -0,793 | 0,45 | 1,193 | 0,118 | 1.06e-19 | 2.02e-18 |
| 13-HODE | -0,005 | -0,71 | 0,498 | 1,143 | 0,123 | 2.19e-17 | 3.91e-16 |
| Glutathione | -0,058 | -0,675 | 0,389 | 0,949 | 0,124 | 8.36e-13 | 1.41e-11 |
| Linoleylcarnitine | 0,012 | -0,534 | 0,401 | 0,911 | 0,125 | 8.23e-12 | 1.32e-10 |
| FA 18(2) | 0,031 | -0,487 | 0,402 | 0,893 | 0,132 | 1.38e-10 | 2.09e-09 |
| Putrescine | -0,005 | -0,365 | 0,256 | 0,718 | 0,11 | 6.49e-10 | 9.39e-09 |
| 9,10-DiHOME | 0,007 | -0,523 | 0,386 | 0,845 | 0,138 | 4.79e-09 | 6.61e-08 |
| 11-HETE | -0,051 | -0,535 | 0,295 | 0,779 | 0,135 | 3.11e-08 | 4.11e-07 |
| Prostaglandin E2 | 0,027 | -0,468 | 0,38 | 0,765 | 0,133 | 3.4e-08 | 4.31e-07 |
| Thromboxane B2 | 0,008 | -0,496 | 0,368 | 0,736 | 0,13 | 5.21e-08 | 6.33e-07 |
| TG(54:2) | -0,002 | 0,45 | -0,326 | -0,791 | 0,145 | 1.61e-07 | 1.88e-06 |
| Lysophosphatidylcholine(O-18:1) | -0,016 | -0,514 | 0,34 | 0,773 | 0,142 | 1.74e-07 | 1.96e-06 |
| Malic acid | -0,034 | -0,412 | 0,236 | 0,71 | 0,134 | 3.36e-07 | 3.64e-06 |
| 9,10,13-TriHOME | 0,016 | 0,462 | -0,302 | -0,718 | 0,141 | 8.24e-07 | 8.64e-06 |
| 9-HODE | 0,014 | -0,422 | 0,325 | 0,686 | 0,137 | 1.16e-06 | 1.17e-05 |
| Sarcosine | -0,021 | 0,356 | -0,293 | -0,669 | 0,134 | 1.39e-06 | 1.36e-05 |
| Prostaglandin F2α | -0,044 | -0,45 | 0,246 | 0,654 | 0,134 | 2.24e-06 | 2.12e-05 |
| 9,12,13-TriHOME | 0,003 | 0,392 | -0,275 | -0,659 | 0,141 | 5.1e-06 | 4.7e-05 |
| TG(52:1) | -0,005 | 0,396 | -0,293 | -0,677 | 0,146 | 6.54e-06 | 5.85e-05 |
| FA 22(6) | 0,023 | -0,308 | 0,26 | 0,659 | 0,143 | 7.68e-06 | 6.67e-05 |
| TG(56:3) | -0,003 | 0,393 | -0,286 | -0,695 | 0,151 | 8.11e-06 | 6.84e-05 |
| TG(51:1) | -0,016 | 0,41 | -0,32 | -0,665 | 0,145 | 8.45e-06 | 6.94e-05 |
| TG(52:2) | -0,023 | 0,359 | -0,296 | -0,656 | 0,147 | 1.39e-05 | 0,000111 |
| Phosphatidylcholine(O-38:4) | 0,03 | 0,409 | -0,241 | -0,655 | 0,148 | 1.72e-05 | 0,000134 |
| TG(51:2) | -0,014 | 0,404 | -0,313 | -0,64 | 0,145 | 1.78e-05 | 0,000135 |
| O-Acetyl-L-serine | -0,007 | -0,208 | 0,14 | 0,603 | 0,138 | 2.03e-05 | 0,000151 |
| Cortisol | 0,018 | 0,341 | -0,213 | -0,608 | 0,146 | 4.62e-05 | 0,000334 |
| TG(50:2) | -0,042 | 0,312 | -0,296 | -0,588 | 0,144 | 6.68e-05 | 0,000468 |
| Glycoursodeoxycholic acid | -0,054 | 0,226 | -0,254 | -0,535 | 0,131 | 6.78e-05 | 0,000468 |
| TG(50:1) | -0,026 | 0,323 | -0,276 | -0,591 | 0,145 | 6.92e-05 | 0,000468 |
| TG(54:3) | -0,005 | 0,325 | -0,241 | -0,611 | 0,152 | 8.83e-05 | 0,000584 |
| TG(48:1) | -0,032 | 0,319 | -0,283 | -0,56 | 0,143 | 0,000122 | 0,00079 |
| Phosphatidylcholine(36:1) | 0,018 | 0,334 | -0,208 | -0,542 | 0,138 | 0,000129 | 0,000815 |
| L-Glutamic acid | -0,015 | -0,294 | 0,187 | 0,563 | 0,145 | 0,000136 | 0,000841 |
| S-Methylcysteine | 0,026 | -0,29 | 0,254 | 0,585 | 0,151 | 0,000141 | 0,00086 |
| Lysophosphatidylethanolamine (16:1) | -0,099 | 0,221 | -0,328 | -0,523 | 0,138 | 0,000208 | 0,00124 |
| TG(53:1) | -0,002 | 0,346 | -0,25 | -0,572 | 0,152 | 0,000231 | 0,00135 |
| 3-Hydroxypropionic Acid | -0,061 | -0,311 | 0,118 | 0,549 | 0,147 | 0,000244 | 0,0014 |
| Lysophosphatidylinositol 18(1) | 0,016 | 0,333 | -0,21 | -0,55 | 0,15 | 0,000328 | 0,00184 |
| TG(56:4) | -0,012 | 0,296 | -0,232 | -0,546 | 0,153 | 0,000474 | 0,00262 |
| Lysophosphatidic acid 16(1) | -0,138 | 0,124 | -0,325 | -0,438 | 0,125 | 0,000541 | 0,00294 |
| TG(58:9) | 0,022 | -0,236 | 0,207 | 0,519 | 0,149 | 0,000622 | 0,00332 |
| CE(22:6) | 0,057 | -0,173 | 0,222 | 0,49 | 0,141 | 0,000645 | 0,00337 |
| TG(56:2) | -0,002 | 0,297 | -0,216 | -0,542 | 0,156 | 0,000654 | 0,00337 |
| Methylmalonylcarnitine | 0,033 | -0,246 | 0,233 | 0,468 | 0,137 | 0,000781 | 0,00396 |
| Lysophosphatidylinositol 16(1) | -0,092 | 0,14 | -0,258 | -0,49 | 0,144 | 8e-04 | 0,00399 |
| TG(54:1) | -0,003 | 0,32 | -0,235 | -0,521 | 0,153 | 0,000815 | 0,004 |
| TG(48:2) | -0,039 | 0,26 | -0,253 | -0,486 | 0,143 | 0,000833 | 0,00402 |
| Phosphatidylethanolamine(34:2) | -0,021 | 0,259 | -0,222 | -0,488 | 0,148 | 0,0012 | 0,00572 |
| TG(46:1) | -0,034 | 0,243 | -0,232 | -0,459 | 0,142 | 0,00141 | 0,00659 |
| Lysophosphatidic acid 18(1) | -0,046 | 0,175 | -0,204 | -0,434 | 0,135 | 0,0015 | 0,00693 |
| 14,15-DiHETE | 0,017 | -0,24 | 0,2 | 0,482 | 0,15 | 0,00156 | 0,00706 |
| Lysophosphatidylcholine(20:5) | 0,052 | -0,155 | 0,199 | 0,408 | 0,127 | 0,00161 | 0,00718 |
| Lysophosphatidylcholine(22:6) | 0,03 | -0,241 | 0,223 | 0,475 | 0,149 | 0,00174 | 0,00767 |
| Ornithine | -0,064 | -0,219 | 0,048 | 0,427 | 0,135 | 0,00187 | 0,00811 |
| 19,20-DiHDPA | 0,049 | -0,176 | 0,21 | 0,465 | 0,148 | 0,00191 | 0,00817 |
| 1-LG/2-LG | -0,027 | 0,219 | -0,204 | -0,442 | 0,142 | 0,00208 | 0,00877 |
| Phosphatidylcholine(38:3) | -0,075 | 0,181 | -0,259 | -0,45 | 0,147 | 0,00263 | 0,011 |
| Phosphatidylcholine(32:1) | -0,079 | 0,196 | -0,275 | -0,423 | 0,14 | 0,00288 | 0,0118 |
| Phosphatidylethanolamine(36:4) | -0,012 | 0,246 | -0,196 | -0,415 | 0,143 | 0,00406 | 0,0163 |
| Lysophosphatidic acid 14(0) | -0,091 | 0,199 | -0,297 | -0,366 | 0,126 | 0,00408 | 0,0163 |
| (+/-) 8-HDoHE | -0,017 | -0,213 | 0,123 | 0,445 | 0,156 | 0,00468 | 0,0185 |
| Lysophosphatidic acid 16(0) | -0,082 | 0,139 | -0,24 | -0,362 | 0,129 | 0,00548 | 0,0214 |
| Taurodeoxycholic acid | 0,008 | 0,219 | -0,142 | -0,392 | 0,142 | 0,00624 | 0,024 |
| Phosphatidylcholine(38:6) | 0,031 | -0,157 | 0,166 | 0,397 | 0,144 | 0,00648 | 0,0246 |
| Taurochenodeoxycholic acid | 0,013 | 0,223 | -0,138 | -0,397 | 0,145 | 0,00665 | 0,0247 |
| OA01 - 2-hydroxybutyric acid | -0,006 | -0,235 | 0,157 | 0,413 | 0,151 | 0,00666 | 0,0247 |
| Phosphatidylcholine(36:5) | 0,004 | -0,152 | 0,116 | 0,393 | 0,145 | 0,00722 | 0,0264 |
| Phosphatidylcholine(40:7) | 0,043 | -0,162 | 0,19 | 0,383 | 0,142 | 0,00761 | 0,0272 |
| Phosphatidylcholine(O-36:4) | 0,021 | 0,214 | -0,116 | -0,422 | 0,157 | 0,00764 | 0,0272 |
| L-Tyrosine | -0,045 | 0,224 | -0,24 | -0,375 | 0,139 | 0,00769 | 0,0272 |
| Lauroylcarnitine | 0,042 | 0,291 | -0,135 | -0,392 | 0,149 | 0,00924 | 0,0323 |
| Phosphatidylcholine(34:1) | 0,053 | 0,112 | 0,01 | -0,1 | 0,038 | 0,00951 | 0,0325 |
| Lysophosphatidic acid 18(2) | -0,032 | 0,125 | -0,144 | -0,368 | 0,141 | 0,00957 | 0,0325 |
| L-Arginine | -0,038 | 0,2 | -0,211 | -0,383 | 0,147 | 0,00963 | 0,0325 |
| TG(51:3) | -0,008 | 0,243 | -0,187 | -0,386 | 0,15 | 0,0111 | 0,0371 |
| TG(50:3) | -0,041 | 0,192 | -0,208 | -0,383 | 0,15 | 0,0116 | 0,0382 |
| FA 22(5) w6 | -0,041 | -0,267 | 0,121 | 0,361 | 0,143 | 0,0124 | 0,0405 |
| L-Asparagine | -0,028 | -0,276 | 0,152 | 0,352 | 0,141 | 0,0134 | 0,0431 |
| Lysophosphatidylethanolamine (20:5) | 0,011 | -0,113 | 0,099 | 0,363 | 0,146 | 0,0135 | 0,0431 |
| Methyldopa | -0,042 | -0,098 | -0,002 | 0,311 | 0,125 | 0,0136 | 0,0431 |
| Lysophosphatidylethanolamine (20:4) | -0,034 | 0,197 | -0,199 | -0,361 | 0,146 | 0,0139 | 0,0435 |
| Phosphatidylcholine(40:8) | 0,067 | -0,125 | 0,204 | 0,31 | 0,126 | 0,0148 | 0,0458 |
| 9-HOTrE | 0,004 | -0,235 | 0,175 | 0,362 | 0,149 | 0,0159 | 0,0488 |
| CE(18:1) | 0,018 | 0,207 | -0,117 | -0,37 | 0,153 | 0,0164 | 0,05 |
| Lysophosphatidylinositol 20(4) | -0,042 | 0,147 | -0,178 | -0,344 | 0,144 | 0,0178 | 0,0536 |
| TG(56:5) | -0,021 | 0,185 | -0,169 | -0,369 | 0,158 | 0,0202 | 0,0599 |
| L-Tryptophan | -0,028 | 0,071 | -0,1 | -0,333 | 0,142 | 0,0203 | 0,0599 |
| Phosphatidylcholine(38:2) | -0,043 | 0,147 | -0,179 | -0,347 | 0,149 | 0,0208 | 0,0609 |
| Myristoilcarnitine | 0,024 | 0,316 | -0,185 | -0,343 | 0,148 | 0,0212 | 0,0612 |
| Glycochenodeoxycholic acid | -0,022 | 0,122 | -0,125 | -0,314 | 0,135 | 0,0213 | 0,0612 |
| Sphingomyelin(d18:1/18:0) | -0,026 | 0,15 | -0,153 | -0,329 | 0,142 | 0,0216 | 0,0615 |
| 1-AG/2-AG | -0,025 | 0,191 | -0,179 | -0,331 | 0,144 | 0,0225 | 0,0633 |
| TG(48:3) | -0,038 | 0,15 | -0,173 | -0,329 | 0,146 | 0,0252 | 0,0703 |
| 17,18-DiHETE | 0,036 | -0,088 | 0,126 | 0,327 | 0,147 | 0,027 | 0,0747 |
| DL-3-aminoisobutyric acid | -0,021 | 0,197 | -0,178 | -0,323 | 0,145 | 0,0274 | 0,0752 |
| Phosphatidylcholine(36:3) | -0,021 | 0,139 | -0,136 | -0,328 | 0,148 | 0,028 | 0,0761 |
| L-Kynurenine | -0,057 | 0,176 | -0,226 | -0,331 | 0,15 | 0,0285 | 0,0762 |
| TG(52:3) | -0,019 | 0,159 | -0,147 | -0,343 | 0,155 | 0,0286 | 0,0762 |
| TG(46:2) | -0,039 | 0,139 | -0,167 | -0,309 | 0,142 | 0,0301 | 0,0797 |
| Cer(d18:1/23:0) | 0,02 | 0,216 | -0,12 | -0,319 | 0,146 | 0,0308 | 0,0807 |
| Linoleoyl ethanolamide | -0,017 | -0,224 | 0,131 | 0,311 | 0,146 | 0,0348 | 0,0905 |
| 5-HETE | -0,098 | -0,28 | 0,031 | 0,3 | 0,143 | 0,0367 | 0,0945 |
| TG(44:1) | -0,019 | 0,149 | -0,139 | -0,289 | 0,142 | 0,0436 | 0,111 |
| DGLEA | -0,077 | 0,108 | -0,21 | -0,296 | 0,146 | 0,044 | 0,112 |
| Lysophosphatidylcholine(16:1) | -0,065 | 0,08 | -0,168 | -0,301 | 0,149 | 0,0449 | 0,113 |
| 1-Methylhistidine | 0,04 | 0,244 | -0,108 | -0,288 | 0,143 | 0,0452 | 0,113 |
| Betaine | 0,052 | -0,11 | 0,167 | 0,287 | 0,144 | 0,0478 | 0,118 |
| TG(54:7) | 0,025 | -0,144 | 0,146 | 0,292 | 0,154 | 0,0587 | 0,144 |
| Citric acid | -0,025 | -0,032 | -0,02 | 0,246 | 0,13 | 0,0607 | 0,147 |
| Lysophosphatidylcholine(20:3) | -0,045 | 0,063 | -0,123 | -0,28 | 0,149 | 0,061 | 0,147 |
| Lysophosphatidylethanolamine(22:6) | 0,016 | -0,088 | 0,09 | 0,27 | 0,143 | 0,0618 | 0,147 |
| Gamma-aminobutyric acid | -0,059 | -0,24 | 0,071 | 0,264 | 0,141 | 0,0623 | 0,147 |
| Cer(d18:1/22:0) | -0,015 | 0,145 | -0,129 | -0,302 | 0,161 | 0,0625 | 0,147 |
| Choline | -0,023 | -0,086 | 0,022 | 0,264 | 0,143 | 0,0671 | 0,156 |
| (+/-) 16-HDoHE | 0,018 | -0,055 | 0,071 | 0,272 | 0,148 | 0,0674 | 0,156 |
| TG(56:6) | -0,007 | 0,147 | -0,117 | -0,29 | 0,158 | 0,0682 | 0,156 |
| Phosphatidylcholine(40:6) | 0,028 | -0,068 | 0,097 | 0,265 | 0,145 | 0,0684 | 0,156 |
| Lactic acid | -0,033 | -0,149 | 0,05 | 0,276 | 0,151 | 0,0695 | 0,158 |
| Phosphatidylcholine(36:2) | 0,017 | 0,126 | -0,061 | -0,273 | 0,153 | 0,0766 | 0,173 |
| Anandamide | -0,11 | 0,01 | -0,196 | -0,265 | 0,15 | 0,0785 | 0,175 |
| Carnitine | -0,003 | 0,188 | -0,139 | -0,246 | 0,14 | 0,0798 | 0,177 |
| L-4-hydroxy-proline | -0,061 | 0,101 | -0,179 | -0,251 | 0,143 | 0,0809 | 0,178 |
| TG(58:8) | 0,016 | -0,08 | 0,085 | 0,267 | 0,154 | 0,0842 | 0,184 |
| O-Anandamide | -0,088 | 0,027 | -0,171 | -0,26 | 0,151 | 0,0872 | 0,189 |
| Sphingomyelin(d18:1/15:0) | 0,008 | 0,234 | -0,154 | -0,247 | 0,144 | 0,0881 | 0,189 |
| Lysophosphatidylcholine(18:1) | 0,063 | 0,161 | -0,008 | -0,259 | 0,151 | 0,0882 | 0,189 |
| Lysophosphatidylethanolamine(18:0) | 0,029 | 0,223 | -0,11 | -0,251 | 0,147 | 0,0889 | 0,189 |
| Decanoylcarnitine | 0,047 | 0,23 | -0,083 | -0,251 | 0,148 | 0,0917 | 0,193 |
| L-Isoleucine | 0,005 | 0,129 | -0,085 | -0,239 | 0,141 | 0,0922 | 0,193 |
| Cer(d18:1/24:1) | -0,028 | 0,118 | -0,133 | -0,263 | 0,157 | 0,0948 | 0,197 |
| Ethanolamine | -0,071 | -0,03 | -0,101 | -0,228 | 0,137 | 0,0971 | 0,201 |
| Cysteine | -0,036 | 0,229 | -0,228 | -0,218 | 0,133 | 0,101 | 0,208 |
| Lysophosphatidylinositol 18(0) | -0,06 | 0,174 | -0,228 | -0,22 | 0,134 | 0,102 | 0,208 |
| Glycodeoxycholic acid | -0,029 | 0,09 | -0,115 | -0,248 | 0,152 | 0,104 | 0,209 |
| TG(55:1) | -0,018 | 0,126 | -0,121 | -0,267 | 0,164 | 0,104 | 0,209 |
| Decenoylcarnitine | 0,035 | -0,098 | 0,13 | 0,245 | 0,151 | 0,105 | 0,209 |
| Sphingomyelin(d18:1/23:0) | 0,006 | 0,12 | -0,076 | -0,248 | 0,152 | 0,105 | 0,209 |
| Propionylcarnitine | 0,001 | 0,197 | -0,139 | -0,218 | 0,134 | 0,106 | 0,209 |
| TG(46:0) | -0,015 | 0,124 | -0,114 | -0,241 | 0,148 | 0,107 | 0,209 |
| 5,6-DiHETrE | 0,033 | 0,137 | -0,042 | -0,233 | 0,145 | 0,11 | 0,215 |
| Tetradecenoylcarnitine | 0,022 | 0,162 | -0,077 | -0,239 | 0,15 | 0,111 | 0,216 |
| TG(54:4) | 0,002 | 0,103 | -0,07 | -0,246 | 0,157 | 0,12 | 0,231 |
| Phosphatidylethanolamine(38:2) | 0,039 | 0,21 | -0,084 | -0,222 | 0,144 | 0,125 | 0,238 |
| 12,13-DiHOME | 0,022 | -0,153 | 0,148 | 0,227 | 0,147 | 0,125 | 0,238 |
| 8,9-DiHETrE | -0,008 | 0,075 | -0,067 | -0,215 | 0,142 | 0,132 | 0,25 |
| Tiglylcarnitine | 0,046 | -0,026 | 0,097 | 0,221 | 0,147 | 0,136 | 0,255 |
| TG(50:4) | -0,023 | 0,101 | -0,113 | -0,225 | 0,15 | 0,136 | 0,255 |
| Cer(d18:0/23:0) | 0,012 | 0,143 | -0,081 | -0,223 | 0,15 | 0,139 | 0,258 |
| TG(56:1) | -0,002 | 0,119 | -0,089 | -0,242 | 0,163 | 0,141 | 0,26 |
| Stearoylcarnitine | 0,06 | 0,253 | -0,079 | -0,205 | 0,141 | 0,147 | 0,269 |
| Deoxycholic acid | -0,034 | 0,086 | -0,12 | -0,219 | 0,151 | 0,148 | 0,27 |
| Cer(d18:0/22:0) | -0,008 | 0,1 | -0,085 | -0,216 | 0,154 | 0,161 | 0,291 |
| TG(60:1) | -0,014 | 0,07 | -0,074 | -0,217 | 0,155 | 0,162 | 0,291 |
| Lysophosphatidylethanolamine(20:4) | 0,003 | 0,108 | -0,073 | -0,213 | 0,152 | 0,164 | 0,294 |
| TG(60:2) | -0,016 | 0,087 | -0,09 | -0,217 | 0,156 | 0,166 | 0,294 |
| Phosphatidylcholine(O-34:2) | 0,015 | 0,145 | -0,078 | -0,231 | 0,166 | 0,166 | 0,294 |
| L-Glutamine | 0,019 | 0,143 | -0,071 | -0,206 | 0,149 | 0,168 | 0,295 |
| Sphingomyelin(d18:1/14:0) | -0,002 | 0,18 | -0,133 | -0,203 | 0,15 | 0,179 | 0,313 |
| Hexanoylcarnitine | 0,034 | 0,209 | -0,091 | -0,202 | 0,15 | 0,181 | 0,314 |
| Phosphatidylcholine(34:3) | -0,047 | 0,059 | -0,123 | -0,199 | 0,149 | 0,183 | 0,315 |
| TG(44:0) | -0,02 | 0,086 | -0,096 | -0,198 | 0,148 | 0,183 | 0,315 |
| Dodecenoylcarnitine | 0,001 | 0,126 | -0,089 | -0,193 | 0,146 | 0,186 | 0,317 |
| L-Homoserine | 0,009 | -0,078 | 0,071 | 0,207 | 0,157 | 0,188 | 0,32 |
| L-2-aminoadipic acid | -0,007 | -0,061 | 0,032 | 0,179 | 0,137 | 0,192 | 0,323 |
| Deoxycarnitine | 0,038 | -0,04 | 0,094 | 0,156 | 0,119 | 0,192 | 0,323 |
| Phosphatidylcholine(O-34:1) | 0,028 | 0,137 | -0,049 | -0,187 | 0,145 | 0,199 | 0,332 |
| TG(42:0) | -0,014 | 0,081 | -0,081 | -0,194 | 0,152 | 0,202 | 0,335 |
| L-Lysine | -0,017 | -0,117 | 0,055 | 0,192 | 0,152 | 0,209 | 0,342 |
| Phosphatidylethanolamine(38:4) | 0,03 | 0,192 | -0,086 | -0,181 | 0,144 | 0,209 | 0,342 |
| TG(58:2) | 0,002 | 0,08 | -0,054 | -0,201 | 0,16 | 0,211 | 0,342 |
| Valerylcarnitine | -0,001 | 0,173 | -0,124 | -0,178 | 0,142 | 0,212 | 0,342 |
| Phosphatidylcholine(38:5) | 0,02 | -0,083 | 0,094 | 0,189 | 0,151 | 0,212 | 0,342 |
| TG(51:4) | -0,013 | 0,126 | -0,112 | -0,188 | 0,151 | 0,214 | 0,344 |
| Sphingomyelin(d18:1/21:0) | -0,007 | 0,121 | -0,098 | -0,187 | 0,15 | 0,215 | 0,344 |
| FA 18(1) | -0,03 | -0,093 | 0,015 | 0,17 | 0,146 | 0,244 | 0,388 |
| Sphingomyelin(d18:1/18:2) | -0,017 | -0,133 | 0,066 | 0,205 | 0,176 | 0,246 | 0,389 |
| TG(59:1) | -0,061 | 0,01 | -0,111 | -0,188 | 0,162 | 0,248 | 0,389 |
| Lysophosphatidylcholine(O-16:0) | 0 | -0,149 | 0,107 | 0,178 | 0,154 | 0,249 | 0,389 |
| TG(60:3) | 0,019 | 0,112 | -0,047 | -0,184 | 0,159 | 0,25 | 0,389 |
| Lysophosphatidylcholine(14:0) | -0,023 | 0,109 | -0,118 | -0,176 | 0,155 | 0,257 | 0,398 |
| TG(42:1) | -0,024 | 0,043 | -0,072 | -0,158 | 0,144 | 0,275 | 0,425 |
| 3-Hydroxybutyric acid | 0,028 | -0,051 | 0,085 | 0,166 | 0,155 | 0,284 | 0,436 |
| DG(36:2) | -0,024 | 0,049 | -0,077 | -0,167 | 0,156 | 0,285 | 0,436 |
| Oleoylcarnitine | -0,015 | -0,07 | 0,025 | 0,156 | 0,147 | 0,287 | 0,436 |
| 12,13-DiHODE | 0,03 | -0,079 | 0,107 | 0,153 | 0,143 | 0,288 | 0,436 |
| Hydroxylysine | -0,085 | 0,069 | -0,197 | -0,158 | 0,152 | 0,299 | 0,449 |
| L-Alpha-aminobutyric acid | 0,068 | -0,035 | 0,143 | 0,154 | 0,148 | 0,3 | 0,449 |
| Sphingomyelin(d18:1/20:0) | -0,026 | 0,04 | -0,073 | -0,156 | 0,151 | 0,302 | 0,449 |
| Tetradecadienylcarnitine | 0,039 | -0,082 | 0,125 | 0,152 | 0,147 | 0,303 | 0,449 |
| TG(44:2) | -0,029 | 0,051 | -0,087 | -0,144 | 0,143 | 0,315 | 0,464 |
| Octanoylcarnitine | 0,045 | 0,175 | -0,048 | -0,15 | 0,151 | 0,321 | 0,47 |
| Butyrylcarnitine | -0,028 | 0,06 | -0,091 | -0,147 | 0,147 | 0,322 | 0,47 |
| Lysophosphatidylcholine(16:0) | 0,015 | 0,044 | -0,007 | -0,15 | 0,152 | 0,325 | 0,472 |
| Isovalerylcarnitine | 0,064 | 0,121 | 0,024 | -0,132 | 0,135 | 0,327 | 0,472 |
| Lysophosphatidylcholine(O-16:1) | 0,016 | -0,119 | 0,112 | 0,151 | 0,154 | 0,33 | 0,472 |
| Homocysteine | -0,015 | 0,153 | -0,137 | -0,135 | 0,139 | 0,331 | 0,472 |
| Phosphatidylcholine(34:2) | -0,005 | 0,042 | -0,038 | -0,148 | 0,152 | 0,331 | 0,472 |
| Phosphatidylcholine(O-38:6) | 0,047 | -0,025 | 0,099 | 0,148 | 0,154 | 0,34 | 0,483 |
| Sphingomyelin(d18:1/25:0) | 0,02 | 0,088 | -0,029 | -0,149 | 0,157 | 0,345 | 0,488 |
| TG(56:7) | 0,009 | -0,055 | 0,055 | 0,147 | 0,158 | 0,353 | 0,497 |
| Pimelylcarnitine | 0,023 | -0,08 | 0,096 | 0,129 | 0,141 | 0,362 | 0,505 |
| Sphingomyelin(d18:1/24:2) | -0,011 | -0,128 | 0,072 | 0,139 | 0,152 | 0,364 | 0,505 |
| Phosphatidylcholine(O-38:5) | 0,028 | 0,072 | -0,004 | -0,146 | 0,16 | 0,364 | 0,505 |
| Dehydroepiandrosteron | 0,039 | 0,023 | 0,05 | 0,13 | 0,145 | 0,37 | 0,512 |
| Taurocholic acid | 0 | 0,064 | -0,045 | -0,129 | 0,144 | 0,373 | 0,513 |
| L-Leucine | 0,013 | 0,055 | -0,018 | -0,119 | 0,136 | 0,383 | 0,524 |
| Lysophosphatidylcholine(18:0) | 0,044 | 0,072 | 0,024 | -0,135 | 0,156 | 0,387 | 0,528 |
| Phosphatidylethanolamine(O-36:5) | 0,034 | -0,072 | 0,111 | 0,136 | 0,158 | 0,391 | 0,528 |
| Phosphatidylethanolamine(O-38:7) | 0,044 | -0,006 | 0,079 | 0,129 | 0,151 | 0,391 | 0,528 |
| Cystathionine | -0,006 | 0,186 | -0,145 | -0,104 | 0,121 | 0,393 | 0,528 |
| Lysophosphatidylcholine(18:3) | -0,005 | 0,065 | -0,055 | -0,14 | 0,171 | 0,414 | 0,554 |
| TG(54:5) | -0,007 | 0,022 | -0,028 | -0,127 | 0,156 | 0,416 | 0,554 |
| Pyroglutamic acid | -0,032 | -0,087 | 0,008 | 0,123 | 0,153 | 0,424 | 0,563 |
| Hexadecenoylcarnitine | -0,016 | 0,121 | -0,114 | -0,116 | 0,148 | 0,435 | 0,572 |
| Lysophosphatidylethanolamine (16:0) | 0,015 | -0,017 | 0,038 | 0,116 | 0,148 | 0,435 | 0,572 |
| Sphingomyelin(d18:1/22:0) | -0,004 | -0,009 | -0,001 | -0,117 | 0,153 | 0,444 | 0,582 |
| Sphingomyelin(d18:1/25:1) | 0,014 | 0,105 | -0,05 | -0,11 | 0,147 | 0,454 | 0,592 |
| Sphingomyelin(d18:1/18:1) | -0,025 | 0,019 | -0,056 | -0,1 | 0,138 | 0,473 | 0,614 |
| Sphingomyelin(d18:1/20:1) | -0,044 | -0,151 | 0,033 | 0,103 | 0,148 | 0,487 | 0,627 |
| 14,15-DiHETrE | -0,033 | -0,032 | -0,034 | -0,102 | 0,146 | 0,487 | 0,627 |
| L-Histidine | -0,012 | -0,037 | 0,006 | -0,104 | 0,152 | 0,492 | 0,631 |
| Isobutyrylcarnitine | 0,072 | 0,106 | 0,047 | 0,094 | 0,144 | 0,514 | 0,657 |
| Palmitoyl ethanolamide | -0,04 | 0,046 | -0,101 | -0,097 | 0,152 | 0,522 | 0,664 |
| Lysophosphatidylcholine(18:2) | 0,064 | 0,032 | 0,087 | -0,096 | 0,15 | 0,525 | 0,664 |
| Phosphatidylcholine(O-36:5) | 0,024 | 0,054 | 0,003 | -0,096 | 0,155 | 0,537 | 0,678 |
| Nonaylcarnitine | 0,038 | 0,173 | -0,059 | -0,088 | 0,144 | 0,545 | 0,682 |
| L-Alanine | -0,053 | -0,086 | -0,029 | 0,095 | 0,157 | 0,546 | 0,682 |
| Citrulline | -0,012 | 0,096 | -0,091 | -0,09 | 0,149 | 0,549 | 0,682 |
| Symmetric dimethylarginine | 0,037 | 0,158 | -0,05 | -0,086 | 0,144 | 0,549 | 0,682 |
| L-Methionine | -0,053 | -0,15 | 0,017 | 0,077 | 0,131 | 0,557 | 0,687 |
| Trimethylamine N-oxide | -0,014 | 0,048 | -0,058 | 0,088 | 0,15 | 0,559 | 0,687 |
| L-Valine | 0,045 | 0,085 | 0,016 | -0,084 | 0,145 | 0,56 | 0,687 |
| CE(18:2) | 0,03 | -0,046 | 0,084 | 0,087 | 0,15 | 0,564 | 0,689 |
| TG(52:4) | -0,013 | 0,003 | -0,025 | -0,087 | 0,157 | 0,581 | 0,703 |
| Phosphatidylcholine(O-36:3) | 0,048 | 0,102 | 0,009 | -0,08 | 0,145 | 0,581 | 0,703 |
| Lysophosphatidylcholine(20:4) | 0,031 | -0,079 | 0,109 | 0,082 | 0,149 | 0,582 | 0,703 |
| Acetylcarnitine | 0,004 | 0,073 | -0,046 | -0,08 | 0,146 | 0,586 | 0,703 |
| L-Threonine | 0,015 | -0,1 | 0,098 | 0,079 | 0,145 | 0,587 | 0,703 |
| Methionine sulfone | 0,028 | 0,053 | 0,011 | 0,077 | 0,148 | 0,604 | 0,72 |
| TG(57:1) | -0,002 | -0,068 | 0,045 | 0,076 | 0,151 | 0,615 | 0,73 |
| Phosphatidylcholine(32:2) | -0,042 | 0,014 | -0,082 | -0,074 | 0,151 | 0,626 | 0,74 |
| Phosphatidylcholine(O-44:5) | 0,047 | -0,001 | 0,081 | 0,075 | 0,155 | 0,63 | 0,742 |
| 2-ketoglutaric acid | -0,073 | -0,104 | -0,051 | 0,067 | 0,141 | 0,634 | 0,744 |
| Phosphatidylcholine(40:5) | -0,036 | 0,009 | -0,068 | -0,073 | 0,154 | 0,637 | 0,744 |
| 3-Methoxytyrosine | -0,049 | 0,076 | -0,14 | 0,048 | 0,104 | 0,646 | 0,752 |
| Glycylglycine | -0,021 | -0,059 | 0,006 | 0,062 | 0,146 | 0,673 | 0,781 |
| TG(56:0) | 0,022 | -0,032 | 0,06 | 0,065 | 0,159 | 0,683 | 0,789 |
| Phosphatidylcholine(O-34:3) | 0,044 | -0,027 | 0,096 | 0,058 | 0,149 | 0,696 | 0,802 |
| Saccharopine | -0,046 | -0,109 | -0,001 | 0,055 | 0,145 | 0,703 | 0,807 |
| Sphingomyelin(d18:1/24:1) | -0,005 | -0,034 | 0,016 | -0,057 | 0,156 | 0,715 | 0,816 |
| Sphingomyelin(d18:1/22:1) | -0,004 | -0,006 | -0,003 | -0,055 | 0,151 | 0,716 | 0,816 |
| Phosphatidylcholine(38:7) | -0,005 | -0,07 | 0,041 | 0,052 | 0,155 | 0,736 | 0,833 |
| Lysophosphatidylethanolamine(18:1) | -0,027 | 0,017 | -0,058 | -0,049 | 0,146 | 0,737 | 0,833 |
| Octenoylcarnitine | -0,012 | -0,042 | 0,01 | -0,047 | 0,15 | 0,753 | 0,845 |
| 2-Methylbutyroylcarnitine | 0,043 | 0,133 | -0,021 | -0,042 | 0,135 | 0,755 | 0,845 |
| Malonylcarnitine | -0,007 | -0,024 | 0,006 | 0,047 | 0,152 | 0,756 | 0,845 |
| Gamma-Glutamylglutamine | -0,057 | -0,118 | -0,013 | 0,044 | 0,143 | 0,759 | 0,845 |
| L-Serine | 0,001 | -0,059 | 0,045 | -0,044 | 0,147 | 0,763 | 0,846 |
| Phosphatidylcholine(38:4) | 0,007 | -0,044 | 0,044 | 0,042 | 0,158 | 0,79 | 0,869 |
| Phosphatidylcholine(36:4) | 0 | -0,055 | 0,04 | 0,041 | 0,155 | 0,791 | 0,869 |
| Phosphatidylcholine(34:4) | -0,039 | -0,002 | -0,066 | -0,039 | 0,146 | 0,792 | 0,869 |
| TG(45:0) | -0,016 | -0,004 | -0,024 | -0,039 | 0,155 | 0,799 | 0,871 |
| Sphingomyelin(d18:1/16:0) | 0,021 | 0,004 | 0,033 | -0,04 | 0,16 | 0,801 | 0,871 |
| 11,12-DiHETrE | -0,03 | -0,078 | 0,004 | -0,036 | 0,145 | 0,802 | 0,871 |
| Glycocholic acid | -0,028 | -0,035 | -0,024 | -0,034 | 0,137 | 0,807 | 0,873 |
| Palmitoylcarnitine | -0,01 | 0,082 | -0,077 | -0,034 | 0,148 | 0,819 | 0,882 |
| Sphingomyelin(d18:1/16:1) | -0,004 | 0,001 | -0,007 | -0,035 | 0,152 | 0,821 | 0,882 |
| 12,13-EpOME | 0,013 | -0,015 | 0,034 | 0,033 | 0,162 | 0,841 | 0,898 |
| Sphingomyelin(d18:1/23:1) | 0,003 | 0,035 | -0,019 | -0,028 | 0,143 | 0,846 | 0,898 |
| TG(42:2) | -0,029 | -0,036 | -0,025 | -0,028 | 0,146 | 0,849 | 0,898 |
| CE(20:4) | 0,014 | -0,01 | 0,032 | 0,03 | 0,159 | 0,85 | 0,898 |
| L-Phenylalanine | -0,047 | -0,045 | -0,048 | -0,026 | 0,141 | 0,851 | 0,898 |
| Homocitrulline | 0,017 | 0,09 | -0,036 | 0,026 | 0,15 | 0,862 | 0,904 |
| 8,12-iPF2a IV | -0,102 | -0,166 | -0,057 | 0,023 | 0,131 | 0,863 | 0,904 |
| DG(36:3) | -0,012 | -0,025 | -0,003 | -0,023 | 0,153 | 0,879 | 0,915 |
| Glycolithocholic acid | -0,047 | -0,077 | -0,026 | 0,023 | 0,15 | 0,879 | 0,915 |
| Lysophosphatidylinositol 18(2) | -0,024 | -0,112 | 0,039 | 0,02 | 0,148 | 0,891 | 0,924 |
| TG(54:0) | 0,021 | 0,031 | 0,013 | -0,02 | 0,161 | 0,9 | 0,928 |
| Cholic acid | -0,043 | -0,02 | -0,059 | 0,018 | 0,143 | 0,903 | 0,928 |
| Gamma-L-glutamyl-L-alanine | -0,04 | -0,019 | -0,055 | -0,018 | 0,149 | 0,903 | 0,928 |
| N6,N6,N6-Trimethyl-L-lysine | -0,019 | 0,016 | -0,044 | -0,016 | 0,138 | 0,908 | 0,929 |
| Sphingomyelin(d18:1/24:0) | 0,002 | -0,074 | 0,056 | 0,017 | 0,154 | 0,911 | 0,929 |
| Phosphatidylethanolamine(O-38:5) | 0,033 | 0,009 | 0,051 | 0,014 | 0,16 | 0,93 | 0,943 |
| Phosphatidylethanolamine(38:6) | 0,036 | 0,08 | 0,005 | -0,012 | 0,138 | 0,931 | 0,943 |
| Taurolithocholic acid | -0,012 | -0,024 | -0,004 | -0,01 | 0,148 | 0,946 | 0,956 |
| Phosphatidylcholine(32:0) | 0,012 | 0,048 | -0,013 | -0,007 | 0,15 | 0,962 | 0,968 |
| L-Proline | -0,048 | -0,044 | -0,051 | 0,002 | 0,143 | 0,989 | 0,993 |
| Glycine | -0,028 | -0,04 | -0,019 | -0,001 | 0,141 | 0,995 | 0,995 |

**Supplementary Table 3. Influence of age at examination on metabolites levels for HC and all IPD patients combined (n = 204).** Shown are the results of the fixed effect meta-analysis: regression coefficient (CI: 95% confidence interval), nominal and adjusted p-values.

| **Name** | **z-value** | **Nominal *p*** | **Adjusted *p*** | **Beta (CI)** |
| --- | --- | --- | --- | --- |
| O-Acetyl-L-serine | 5,994 | 2.05e-09 | 6.24e-07 | 0,032 (0,021;0,042) |
| Cystathionine | 4,981 | 6.31e-07 | 9.59e-05 | 0,023 (0,014;0,032) |
| Cysteine | 4,769 | 1.85e-06 | 0,000187 | 0,024 (0,014;0,034) |
| 3-Methoxytyrosine | 4,278 | 1.88e-05 | 0,00143 | 0,017 (0,009;0,024) |
| Citric acid | 4,207 | 2.59e-05 | 0,00155 | 0,021 (0,011;0,03) |
| Sphingomyelin(d18:1/15:0) | 4,169 | 3.07e-05 | 0,00155 | 0,023 (0,012;0,033) |
| Sphingomyelin(d18:1/22:0) | -3,865 | 0,000111 | 0,00482 | -0,022 (-0,033;-0,011) |
| Isobutyrylcarnitine | 3,743 | 0,000182 | 0,00614 | 0,02 (0,01;0,031) |
| Lysophosphatidylinositol 18(0) | 3,763 | 0,000168 | 0,00614 | 0,019 (0,009;0,029) |
| L-Tryptophan | -3,664 | 0,000248 | 0,00717 | -0,02 (-0,03;-0,009) |
| Trimethylamine N-oxide | 3,653 | 0,000259 | 0,00717 | 0,021 (0,01;0,032) |
| Lysophosphatidylethanolamine (20:5) | 3,625 | 0,000289 | 0,00732 | 0,02 (0,009;0,031) |
| Lysophosphatidic acid 14(0) | 3,581 | 0,000342 | 0,00799 | 0,017 (0,008;0,027) |
| Sphingomyelin(d18:1/24:0) | -3,562 | 0,000368 | 0,00799 | -0,02 (-0,032;-0,009) |
| Nonaylcarnitine | 3,524 | 0,000425 | 0,00819 | 0,019 (0,009;0,03) |
| Isocitrate | 3,52 | 0,000431 | 0,00819 | 0,015 (0,007;0,024) |
| Homocitrulline | 3,458 | 0,000544 | 0,00918 | 0,02 (0,009;0,031) |
| Lysophosphatidylcholine(18:2) | -3,47 | 0,000521 | 0,00918 | -0,02 (-0,031;-0,008) |
| Ethanolamine | -3,312 | 0,000925 | 0,0148 | -0,017 (-0,027;-0,007) |
| Choline | 3,283 | 0,00103 | 0,0156 | 0,018 (0,007;0,029) |
| (+/-) 16-HDoHE | 3,195 | 0,0014 | 0,0202 | 0,018 (0,007;0,028) |
| Phosphatidylcholine(36:5) | 3,175 | 0,0015 | 0,0207 | 0,017 (0,007;0,028) |
| Myristoilcarnitine | 3,152 | 0,00162 | 0,0215 | 0,017 (0,006;0,027) |
| Lysophosphatidylinositol 18(2) | -3,103 | 0,00191 | 0,0242 | -0,017 (-0,029;-0,006) |
| L-Threonine | -3,075 | 0,00211 | 0,0256 | -0,017 (-0,028;-0,006) |
| CE(22:6) | 2,993 | 0,00276 | 0,0323 | 0,016 (0,005;0,026) |
| Methionine sulfone | 2,974 | 0,00294 | 0,0331 | 0,017 (0,006;0,028) |
| Lysophosphatidylcholine(20:3) | -2,925 | 0,00344 | 0,0374 | -0,016 (-0,027;-0,005) |
| 8-HETE | -2,885 | 0,00392 | 0,0411 | -0,012 (-0,02;-0,004) |
| Lysophosphatidylcholine(20:4) | -2,827 | 0,0047 | 0,0476 | -0,016 (-0,027;-0,005) |
| Symmetric dimethylarginine | 2,789 | 0,00529 | 0,0519 | 0,015 (0,005;0,026) |
| L-Histidine | -2,71 | 0,00674 | 0,0622 | -0,016 (-0,027;-0,004) |
| Tiglylcarnitine | 2,709 | 0,00675 | 0,0622 | 0,015 (0,004;0,026) |
| L-Serine | -2,698 | 0,00698 | 0,0624 | -0,015 (-0,026;-0,004) |
| Glutathione | -2,652 | 0,00801 | 0,0696 | -0,012 (-0,021;-0,003) |
| Homocysteine | 2,629 | 0,00856 | 0,0723 | 0,014 (0,004;0,024) |
| Methyldopa | 2,59 | 0,00959 | 0,0782 | 0,012 (0,003;0,021) |
| Lysophosphatidylinositol 16(1) | -2,566 | 0,0103 | 0,0782 | -0,014 (-0,025;-0,003) |
| Dehydroepiandrosteron | 2,583 | 0,00979 | 0,0782 | 0,014 (0,003;0,025) |
| Phosphatidylethanolamine(38:2) | 2,57 | 0,0102 | 0,0782 | 0,014 (0,003;0,025) |
| Sphingosine 1-phosphate 18(2) | -2,521 | 0,0117 | 0,085 | -0,008 (-0,015;-0,002) |
| 14,15-DiHETrE | -2,52 | 0,0117 | 0,085 | -0,014 (-0,025;-0,003) |
| Sphingomyelin(d18:1/14:0) | 2,506 | 0,0122 | 0,0864 | 0,014 (0,003;0,025) |
| Ornithine | 2,481 | 0,0131 | 0,0904 | 0,013 (0,003;0,023) |
| 11,12-DiHETrE | -2,469 | 0,0135 | 0,0914 | -0,014 (-0,024;-0,003) |
| Phosphatidylethanolamine(38:4) | 2,459 | 0,0139 | 0,092 | 0,013 (0,003;0,023) |
| Propionylcarnitine | 2,397 | 0,0165 | 0,102 | 0,012 (0,002;0,022) |
| Stearoylcarnitine | 2,401 | 0,0164 | 0,102 | 0,012 (0,002;0,022) |
| Sphingomyelin(d18:1/20:1) | -2,405 | 0,0162 | 0,102 | -0,014 (-0,025;-0,002) |
| Phosphatidylcholine(40:6) | 2,387 | 0,017 | 0,103 | 0,013 (0,002;0,024) |
| Lysophosphatidylethanolamine(18:0) | 2,379 | 0,0174 | 0,103 | 0,013 (0,002;0,024) |
| Lysophosphatidylethanolamine(22:6) | 2,363 | 0,0181 | 0,106 | 0,013 (0,002;0,023) |
| Valerylcarnitine | 2,345 | 0,019 | 0,107 | 0,013 (0,002;0,023) |
| Lysophosphatidylcholine(18:1) | -2,33 | 0,0198 | 0,107 | -0,013 (-0,024;-0,002) |
| Sphingomyelin(d18:1/24:1) | -2,331 | 0,0197 | 0,107 | -0,014 (-0,025;-0,002) |
| TG(59:1) | -2,34 | 0,0193 | 0,107 | -0,014 (-0,026;-0,002) |
| 17,18-DiHETE | 2,295 | 0,0217 | 0,116 | 0,013 (0,002;0,024) |
| Palmitoylcarnitine | 2,21 | 0,0271 | 0,141 | 0,012 (0,001;0,022) |
| Thromboxane-B2 | -2,206 | 0,0274 | 0,141 | -0,011 (-0,02;-0,001) |
| 2-Methylbutyroylcarnitine | 2,179 | 0,0293 | 0,148 | 0,011 (0,001;0,021) |
| Citrulline | 2,161 | 0,0307 | 0,151 | 0,012 (0,001;0,023) |
| Hexadecenoylcarnitine | 2,164 | 0,0305 | 0,151 | 0,012 (0,001;0,022) |
| Phosphatidylcholine(38:6) | 2,154 | 0,0313 | 0,151 | 0,012 (0,001;0,022) |
| L-Methionine | -2,146 | 0,0319 | 0,151 | -0,011 (-0,02;-0,001) |
| Hydroxylysine | 2,076 | 0,0379 | 0,176 | 0,012 (0,001;0,023) |
| 13-HODE | -2,073 | 0,0382 | 0,176 | -0,009 (-0,018;-0,001) |
| FA 22(6) | 2,009 | 0,0446 | 0,197 | 0,011 (0;0,022) |
| 12,13-DiHOME | -2,019 | 0,0435 | 0,197 | -0,011 (-0,022;0) |
| Lysophosphatidylcholine(16:0) | -2,008 | 0,0447 | 0,197 | -0,011 (-0,023;0) |
| 8,9-DiHETrE | -1,985 | 0,0472 | 0,205 | -0,011 (-0,021;0) |
| Sphingomyelin(d18:1/20:0) | -1,977 | 0,048 | 0,206 | -0,011 (-0,022;0) |
| 9,10-DiHOME | -1,953 | 0,0509 | 0,215 | -0,01 (-0,02;0) |
| Phosphatidylethanolamine(38:6) | 1,929 | 0,0537 | 0,224 | 0,01 (0;0,02) |
| 12-HETE | -1,886 | 0,0593 | 0,244 | -0,004 (-0,009;0) |
| Putrescine | 1,868 | 0,0618 | 0,248 | 0,007 (0;0,015) |
| TG(54:5) | -1,867 | 0,0619 | 0,248 | -0,011 (-0,022;0,001) |
| Hexanoylcarnitine | 1,86 | 0,0629 | 0,248 | 0,011 (-0,001;0,022) |
| 8,12-iPF2a IV | -1,848 | 0,0646 | 0,249 | -0,009 (-0,019;0,001) |
| TG(54:4) | -1,847 | 0,0648 | 0,249 | -0,011 (-0,022;0,001) |
| Octenoylcarnitine | -1,832 | 0,0669 | 0,254 | -0,01 (-0,022;0,001) |
| 19,20-DiHDPA | 1,819 | 0,0688 | 0,258 | 0,01 (-0,001;0,021) |
| Phosphatidylcholine(40:7) | 1,81 | 0,0703 | 0,261 | 0,01 (-0,001;0,02) |
| Octanoylcarnitine | 1,794 | 0,0729 | 0,267 | 0,01 (-0,001;0,021) |
| TG(58:8) | 1,784 | 0,0744 | 0,269 | 0,01 (-0,001;0,022) |
| (+/-) 8-HDoHE | 1,764 | 0,0777 | 0,278 | 0,01 (-0,001;0,022) |
| L-Glutamic acid | 1,758 | 0,0788 | 0,279 | 0,009 (-0,001;0,019) |
| Lysophosphatidylcholine(16:1) | -1,714 | 0,0866 | 0,302 | -0,01 (-0,021;0,001) |
| O-Anandamide | -1,708 | 0,0876 | 0,303 | -0,01 (-0,021;0,001) |
| Decanoylcarnitine | 1,691 | 0,0908 | 0,307 | 0,009 (-0,001;0,02) |
| 15(S)-HETrE | -1,692 | 0,0907 | 0,307 | -0,007 (-0,016;0,001) |
| O-Phosphoethanolamine | -1,671 | 0,0947 | 0,31 | -0,003 (-0,007;0,001) |
| Lysophosphatidylcholine(18:0) | -1,666 | 0,0957 | 0,31 | -0,01 (-0,021;0,002) |
| Phosphatidylcholine(32:0) | 1,671 | 0,0947 | 0,31 | 0,009 (-0,002;0,021) |
| TG(51:2) | 1,667 | 0,0954 | 0,31 | 0,009 (-0,002;0,019) |
| Lysophosphatidylinositol 20(4) | -1,64 | 0,101 | 0,32 | -0,009 (-0,02;0,002) |
| Sphingomyelin(d18:1/18:0) | -1,641 | 0,101 | 0,32 | -0,009 (-0,019;0,002) |
| Sphingomyelin(d18:1/22:1) | -1,624 | 0,104 | 0,326 | -0,009 (-0,02;0,002) |
| TG(58:9) | 1,621 | 0,105 | 0,326 | 0,009 (-0,002;0,02) |
| TG(54:3) | -1,613 | 0,107 | 0,328 | -0,009 (-0,02;0,002) |
| Gamma-Glutamylglutamine | -1,603 | 0,109 | 0,331 | -0,009 (-0,019;0,002) |
| L-Kynurenine | 1,582 | 0,114 | 0,335 | 0,009 (-0,002;0,02) |
| Carnitine | 1,592 | 0,111 | 0,335 | 0,008 (-0,002;0,019) |
| Phosphatidylethanolamine(O-38:7) | 1,584 | 0,113 | 0,335 | 0,009 (-0,002;0,02) |
| Anandamide | -1,572 | 0,116 | 0,336 | -0,009 (-0,02;0,002) |
| Sphingomyelin(d18:1/25:1) | 1,575 | 0,115 | 0,336 | 0,009 (-0,002;0,019) |
| L-2-aminoadipic acid | 1,554 | 0,12 | 0,336 | 0,008 (-0,002;0,018) |
| L-Leucine | -1,552 | 0,121 | 0,336 | -0,008 (-0,018;0,002) |
| Lysophosphatidylethanolamine (16:0) | 1,557 | 0,119 | 0,336 | 0,009 (-0,002;0,02) |
| TG(51:1) | 1,559 | 0,119 | 0,336 | 0,008 (-0,002;0,018) |
| 1-Methylhistidine | 1,53 | 0,126 | 0,348 | 0,008 (-0,002;0,019) |
| 9-HOTrE | -1,495 | 0,135 | 0,369 | -0,008 (-0,019;0,003) |
| Sphingomyelin(d18:1/23:1) | 1,477 | 0,14 | 0,379 | 0,008 (-0,003;0,019) |
| Saccharopine | -1,469 | 0,142 | 0,382 | -0,008 (-0,019;0,003) |
| S-Methylcysteine | 1,453 | 0,146 | 0,386 | 0,008 (-0,003;0,02) |
| Cholic acid | 1,453 | 0,146 | 0,386 | 0,008 (-0,003;0,019) |
| 5,6-DiHETrE | -1,435 | 0,151 | 0,394 | -0,008 (-0,018;0,003) |
| 9-HODE | -1,43 | 0,153 | 0,394 | -0,007 (-0,017;0,003) |
| Phosphatidylcholine(O-36:4) | -1,43 | 0,153 | 0,394 | -0,008 (-0,02;0,003) |
| L-Asparagine | -1,402 | 0,161 | 0,411 | -0,008 (-0,018;0,003) |
| L-Isoleucine | -1,372 | 0,17 | 0,425 | -0,007 (-0,018;0,003) |
| L-Tyrosine | 1,371 | 0,17 | 0,425 | 0,007 (-0,003;0,018) |
| Lysophosphatidylcholine(O-16:1) | -1,378 | 0,168 | 0,425 | -0,008 (-0,019;0,003) |
| TG(60:1) | -1,356 | 0,175 | 0,433 | -0,008 (-0,019;0,003) |
| TG(52:4) | -1,351 | 0,177 | 0,433 | -0,008 (-0,019;0,004) |
| DL-3-aminoisobutyric acid | 1,322 | 0,186 | 0,439 | 0,007 (-0,003;0,018) |
| Isovalerylcarnitine | -1,327 | 0,184 | 0,439 | -0,007 (-0,017;0,003) |
| Lysophosphatidylcholine(O-16:0) | -1,326 | 0,185 | 0,439 | -0,007 (-0,018;0,004) |
| Lysophosphatidylcholine(O-18:1) | -1,33 | 0,183 | 0,439 | -0,007 (-0,017;0,003) |
| Phosphatidylcholine(36:2) | -1,33 | 0,184 | 0,439 | -0,008 (-0,019;0,004) |
| Lysophosphatidic acid 18(2) | -1,299 | 0,194 | 0,446 | -0,007 (-0,017;0,004) |
| Sphingomyelin(d18:1/18:1) | -1,308 | 0,191 | 0,446 | -0,007 (-0,017;0,003) |
| TG(58:2) | -1,301 | 0,193 | 0,446 | -0,008 (-0,019;0,004) |
| CE(18:1) | -1,283 | 0,2 | 0,453 | -0,007 (-0,019;0,004) |
| Lysophosphatidylcholine(20:5) | 1,286 | 0,198 | 0,453 | 0,006 (-0,003;0,015) |
| Lysophosphatidic acid 16(0) | 1,258 | 0,208 | 0,469 | 0,006 (-0,003;0,015) |
| Prostaglandin E2 | -1,249 | 0,212 | 0,473 | -0,006 (-0,016;0,004) |
| Phosphatidylcholine(36:3) | -1,223 | 0,221 | 0,485 | -0,007 (-0,018;0,004) |
| Phosphatidylcholine(38:7) | -1,231 | 0,218 | 0,485 | -0,007 (-0,018;0,004) |
| TG(51:4) | 1,222 | 0,222 | 0,485 | 0,007 (-0,004;0,017) |
| Taurine | -1,186 | 0,236 | 0,504 | -0,003 (-0,009;0,002) |
| Sphingomyelin(d18:1/24:2) | -1,184 | 0,237 | 0,504 | -0,007 (-0,018;0,004) |
| TG(52:3) | -1,187 | 0,235 | 0,504 | -0,007 (-0,018;0,004) |
| TG(54:2) | -1,182 | 0,237 | 0,504 | -0,006 (-0,017;0,004) |
| Glycine | -1,114 | 0,265 | 0,552 | -0,006 (-0,016;0,004) |
| Sphingomyelin(d18:1/23:0) | -1,116 | 0,264 | 0,552 | -0,006 (-0,017;0,005) |
| TG(56:3) | -1,119 | 0,263 | 0,552 | -0,006 (-0,017;0,005) |
| TG(60:2) | -1,107 | 0,268 | 0,555 | -0,006 (-0,018;0,005) |
| TG(56:6) | -1,1 | 0,271 | 0,557 | -0,006 (-0,018;0,005) |
| Acetylcarnitine | 1,08 | 0,28 | 0,568 | 0,006 (-0,005;0,017) |
| Malic acid | 1,055 | 0,291 | 0,568 | 0,005 (-0,005;0,015) |
| Linoleoyl ethanolamide | -1,059 | 0,29 | 0,568 | -0,006 (-0,016;0,005) |
| Lysophosphatidylcholine(18:3) | -1,058 | 0,29 | 0,568 | -0,007 (-0,019;0,006) |
| Phosphatidylcholine(36:4) | -1,056 | 0,291 | 0,568 | -0,006 (-0,017;0,005) |
| Sphingomyelin(d18:1/16:0) | -1,056 | 0,291 | 0,568 | -0,006 (-0,018;0,005) |
| Sphingomyelin(d18:1/21:0) | 1,056 | 0,291 | 0,568 | 0,006 (-0,005;0,017) |
| TG(56:5) | -1,079 | 0,281 | 0,568 | -0,006 (-0,017;0,005) |
| Oleoylcarnitine | 1,035 | 0,301 | 0,569 | 0,006 (-0,005;0,016) |
| Cortisol | -1,029 | 0,303 | 0,569 | -0,006 (-0,016;0,005) |
| Lysophosphatidylethanolamine(20:4) | -1,034 | 0,301 | 0,569 | -0,006 (-0,017;0,005) |
| Phosphatidylcholine(O-38:6) | 1,034 | 0,301 | 0,569 | 0,006 (-0,005;0,018) |
| Phosphatidylcholine(O-44:5) | -1,044 | 0,297 | 0,569 | -0,006 (-0,018;0,005) |
| TG(51:3) | 1,032 | 0,302 | 0,569 | 0,005 (-0,005;0,016) |
| Lauroylcarnitine | 1,015 | 0,31 | 0,575 | 0,006 (-0,005;0,016) |
| Tetradecadienylcarnitine | -1,016 | 0,31 | 0,575 | -0,006 (-0,016;0,005) |
| Deoxycarnitine | 0,999 | 0,318 | 0,585 | 0,004 (-0,004;0,012) |
| Phosphatidylcholine(38:4) | -0,986 | 0,324 | 0,594 | -0,006 (-0,017;0,006) |
| Decenoylcarnitine | 0,952 | 0,341 | 0,597 | 0,005 (-0,006;0,017) |
| 3-Hydroxybutyric acid | 0,941 | 0,347 | 0,597 | 0,006 (-0,006;0,017) |
| Lysophosphatidic acid 18(1) | -0,955 | 0,339 | 0,597 | -0,005 (-0,015;0,005) |
| LPS 18(1) | -0,94 | 0,347 | 0,597 | -0,003 (-0,009;0,003) |
| (+/-) 10-HDoHE | 0,968 | 0,333 | 0,597 | 0,003 (-0,003;0,009) |
| Glycolithocholic acid | -0,958 | 0,338 | 0,597 | -0,005 (-0,016;0,006) |
| Prostaglandin F2α | -0,941 | 0,347 | 0,597 | -0,004 (-0,014;0,005) |
| Cer(d18:1/23:0) | 0,956 | 0,339 | 0,597 | 0,005 (-0,006;0,016) |
| Phosphatidylcholine(34:2) | -0,94 | 0,347 | 0,597 | -0,005 (-0,017;0,006) |
| Phosphatidylcholine(38:3) | -0,942 | 0,346 | 0,597 | -0,005 (-0,016;0,006) |
| TG(60:3) | -0,949 | 0,343 | 0,597 | -0,005 (-0,017;0,006) |
| 11-HETE | -0,923 | 0,356 | 0,605 | -0,005 (-0,015;0,005) |
| TG(52:2) | -0,922 | 0,356 | 0,605 | -0,005 (-0,016;0,006) |
| Lysophosphatidylethanolamine (20:4) | 0,905 | 0,365 | 0,615 | 0,005 (-0,006;0,016) |
| TG(56:4) | -0,904 | 0,366 | 0,615 | -0,005 (-0,016;0,006) |
| Phosphatidylcholine(O-38:5) | -0,9 | 0,368 | 0,615 | -0,005 (-0,017;0,006) |
| 14,15-DiHETE | 0,884 | 0,377 | 0,626 | 0,005 (-0,006;0,016) |
| 1-AG/2-AG | 0,865 | 0,387 | 0,64 | 0,005 (-0,006;0,015) |
| Pyroglutamic acid | 0,835 | 0,404 | 0,664 | 0,005 (-0,006;0,016) |
| TG(48:1) | 0,825 | 0,409 | 0,669 | 0,004 (-0,006;0,015) |
| Taurochenodeoxycholic acid | -0,816 | 0,415 | 0,674 | -0,004 (-0,015;0,006) |
| Lysophosphatidylcholine(14:0) | 0,81 | 0,418 | 0,676 | 0,005 (-0,007;0,016) |
| Serotonine | -0,788 | 0,431 | 0,689 | -0,003 (-0,009;0,004) |
| 3-Hydroxypropionic Acid | 0,784 | 0,433 | 0,689 | 0,004 (-0,006;0,014) |
| Sphingosine 1-phosphate 18(1) | -0,783 | 0,434 | 0,689 | -0,002 (-0,006;0,003) |
| Glycodeoxycholic acid | -0,778 | 0,437 | 0,689 | -0,004 (-0,016;0,007) |
| Glycoursodeoxycholic acid | -0,776 | 0,438 | 0,689 | -0,003 (-0,012;0,005) |
| FA 18(1) | 0,766 | 0,444 | 0,693 | 0,004 (-0,007;0,015) |
| 12,13-EpOME | -0,765 | 0,444 | 0,693 | -0,005 (-0,016;0,007) |
| Phosphatidylcholine(38:5) | 0,758 | 0,449 | 0,696 | 0,004 (-0,007;0,015) |
| L-Valine | -0,748 | 0,454 | 0,696 | -0,004 (-0,015;0,007) |
| 8(9)-EpETrE | -0,752 | 0,452 | 0,696 | -0,002 (-0,008;0,004) |
| TG(56:7) | 0,746 | 0,456 | 0,696 | 0,004 (-0,007;0,016) |
| Lactic acid | 0,74 | 0,459 | 0,698 | 0,004 (-0,007;0,014) |
| Sphingosine 1-phosphate 18(0) | 0,713 | 0,476 | 0,713 | 0,002 (-0,003;0,006) |
| Phosphatidylethanolamine(36:4) | 0,719 | 0,472 | 0,713 | 0,004 (-0,007;0,014) |
| Sphingomyelin(d18:1/25:0) | -0,716 | 0,474 | 0,713 | -0,004 (-0,016;0,007) |
| Deoxycholic acid | -0,7 | 0,484 | 0,721 | -0,004 (-0,015;0,007) |
| Betaine | 0,691 | 0,49 | 0,723 | 0,004 (-0,007;0,014) |
| Taurolithocholic acid | -0,69 | 0,49 | 0,723 | -0,004 (-0,015;0,007) |
| Phosphatidylcholine(O-38:4) | -0,684 | 0,494 | 0,725 | -0,004 (-0,015;0,007) |
| Gamma-L-glutamyl-L-alanine | -0,656 | 0,512 | 0,727 | -0,004 (-0,014;0,007) |
| Sarcosine | -0,655 | 0,513 | 0,727 | -0,003 (-0,013;0,007) |
| FA 22(5) w6 | -0,668 | 0,504 | 0,727 | -0,004 (-0,014;0,007) |
| (+/-) 14-HDoHE | 0,659 | 0,51 | 0,727 | 0,001 (-0,003;0,005) |
| Phosphatidylcholine(32:1) | 0,657 | 0,511 | 0,727 | 0,003 (-0,007;0,014) |
| Sphingomyelin(d18:1/16:1) | -0,652 | 0,514 | 0,727 | -0,004 (-0,015;0,008) |
| TG(44:1) | 0,655 | 0,512 | 0,727 | 0,003 (-0,006;0,012) |
| TG(46:2) | 0,663 | 0,507 | 0,727 | 0,003 (-0,006;0,012) |
| TG(54:0) | -0,644 | 0,519 | 0,731 | -0,004 (-0,016;0,008) |
| Lysophosphatidylinositol 18(1) | -0,634 | 0,526 | 0,733 | -0,004 (-0,015;0,008) |
| TG(46:1) | 0,637 | 0,524 | 0,733 | 0,003 (-0,006;0,013) |
| Linoleylcarnitine | -0,626 | 0,531 | 0,733 | -0,003 (-0,012;0,006) |
| Phosphatidylcholine(36:1) | -0,627 | 0,531 | 0,733 | -0,003 (-0,013;0,007) |
| Phosphatidylcholine(O-34:3) | -0,623 | 0,533 | 0,733 | -0,004 (-0,015;0,008) |
| Phosphatidylcholine(34:4) | 0,619 | 0,536 | 0,734 | 0,003 (-0,007;0,014) |
| DGLEA | 0,586 | 0,558 | 0,761 | 0,003 (-0,008;0,014) |
| L-Alpha-aminobutyric acid | -0,572 | 0,568 | 0,761 | -0,003 (-0,014;0,008) |
| Malonylcarnitine | 0,561 | 0,575 | 0,761 | 0,003 (-0,008;0,014) |
| 2-ketoglutaric acid | 0,569 | 0,57 | 0,761 | 0,003 (-0,007;0,013) |
| Phosphatidylcholine(32:2) | 0,559 | 0,576 | 0,761 | 0,003 (-0,008;0,014) |
| Phosphatidylcholine(O-34:1) | 0,566 | 0,572 | 0,761 | 0,003 (-0,008;0,014) |
| TG(54:1) | 0,579 | 0,562 | 0,761 | 0,003 (-0,008;0,015) |
| TG(56:2) | -0,568 | 0,57 | 0,761 | -0,003 (-0,013;0,007) |
| TG(48:2) | 0,545 | 0,586 | 0,768 | 0,003 (-0,007;0,013) |
| TG(48:3) | 0,547 | 0,585 | 0,768 | 0,003 (-0,007;0,013) |
| Glycylglycine | -0,53 | 0,596 | 0,774 | -0,003 (-0,013;0,008) |
| (+/-) 11-HDoHE | 0,526 | 0,599 | 0,774 | 0,002 (-0,005;0,008) |
| TG(42:0) | 0,527 | 0,598 | 0,774 | 0,002 (-0,006;0,011) |
| Phosphatidylcholine(40:8) | -0,518 | 0,604 | 0,779 | -0,002 (-0,012;0,007) |
| Taurocholic acid | 0,51 | 0,61 | 0,783 | 0,003 (-0,008;0,013) |
| Dodecenoylcarnitine | 0,497 | 0,619 | 0,788 | 0,003 (-0,008;0,013) |
| TG(44:0) | 0,496 | 0,62 | 0,788 | 0,002 (-0,006;0,011) |
| 1-LG/2-LG | -0,492 | 0,623 | 0,789 | -0,003 (-0,013;0,008) |
| Sphingomyelin(d18:1/18:2) | 0,482 | 0,63 | 0,795 | 0,003 (-0,009;0,015) |
| L-Phenylalanine | -0,435 | 0,663 | 0,795 | -0,002 (-0,013;0,008) |
| Pimelylcarnitine | -0,422 | 0,673 | 0,795 | -0,002 (-0,013;0,008) |
| FA 18(2) | 0,413 | 0,68 | 0,795 | 0,002 (-0,008;0,012) |
| 12,13-DiHODE | -0,436 | 0,663 | 0,795 | -0,002 (-0,013;0,008) |
| 12(S)-HEPE | 0,452 | 0,652 | 0,795 | 0,001 (-0,003;0,005) |
| 5-HETE | -0,467 | 0,641 | 0,795 | -0,003 (-0,013;0,008) |
| Glycochenodeoxycholic acid | -0,407 | 0,684 | 0,795 | -0,002 (-0,012;0,008) |
| Palmitoyl ethanolamide | 0,442 | 0,659 | 0,795 | 0,003 (-0,009;0,014) |
| Cer(d18:0/22:0) | -0,47 | 0,639 | 0,795 | -0,003 (-0,014;0,009) |
| DG(36:3) | -0,417 | 0,676 | 0,795 | -0,002 (-0,012;0,008) |
| Phosphatidylcholine(34:1) | -0,412 | 0,681 | 0,795 | -0,001 (-0,003;0,002) |
| Phosphatidylcholine(38:2) | -0,427 | 0,67 | 0,795 | -0,002 (-0,013;0,009) |
| Phosphatidylcholine(O-36:3) | -0,438 | 0,661 | 0,795 | -0,002 (-0,012;0,008) |
| TG(42:2) | -0,423 | 0,673 | 0,795 | -0,002 (-0,012;0,008) |
| TG(44:2) | 0,441 | 0,659 | 0,795 | 0,002 (-0,007;0,011) |
| TG(46:0) | 0,413 | 0,679 | 0,795 | 0,002 (-0,007;0,011) |
| TG(50:1) | -0,424 | 0,672 | 0,795 | -0,002 (-0,013;0,008) |
| TG(52:1) | -0,457 | 0,647 | 0,795 | -0,002 (-0,013;0,008) |
| TG(53:1) | 0,405 | 0,685 | 0,795 | 0,002 (-0,008;0,012) |
| TG(56:1) | 0,45 | 0,653 | 0,795 | 0,002 (-0,007;0,011) |
| TG(57:1) | -0,433 | 0,665 | 0,795 | -0,002 (-0,013;0,008) |
| L-Homoserine | 0,379 | 0,704 | 0,814 | 0,002 (-0,009;0,014) |
| Phosphatidylethanolamine(34:2) | 0,348 | 0,728 | 0,838 | 0,002 (-0,009;0,013) |
| Lysophosphatidylethanolamine(18:1) | -0,338 | 0,735 | 0,843 | -0,002 (-0,011;0,008) |
| L-4-hydroxy-proline | 0,33 | 0,741 | 0,844 | 0,002 (-0,009;0,012) |
| Lysophosphatidic acid 16(1) | 0,331 | 0,74 | 0,844 | 0,002 (-0,008;0,011) |
| 9,12,13-TriHOME | -0,322 | 0,747 | 0,844 | -0,002 (-0,012;0,009) |
| Cer(d18:0/23:0) | 0,325 | 0,745 | 0,844 | 0,002 (-0,009;0,013) |
| Butyrylcarnitine | -0,295 | 0,768 | 0,861 | -0,002 (-0,013;0,009) |
| Phosphatidylcholine(O-36:5) | -0,295 | 0,768 | 0,861 | -0,002 (-0,013;0,01) |
| L-Arginine | 0,261 | 0,794 | 0,884 | 0,001 (-0,009;0,012) |
| CE(18:2) | -0,264 | 0,792 | 0,884 | -0,001 (-0,013;0,01) |
| TG(42:1) | -0,248 | 0,804 | 0,889 | -0,001 (-0,011;0,008) |
| TG(55:1) | -0,249 | 0,804 | 0,889 | -0,002 (-0,013;0,01) |
| Phosphatidylcholine(40:5) | -0,21 | 0,833 | 0,915 | -0,001 (-0,012;0,01) |
| TG(50:2) | -0,211 | 0,833 | 0,915 | -0,001 (-0,011;0,009) |
| Phosphatidylethanolamine(O-36:5) | -0,206 | 0,836 | 0,915 | -0,001 (-0,013;0,01) |
| TG(56:0) | 0,178 | 0,859 | 0,936 | 0,001 (-0,01;0,012) |
| L-Proline | -0,169 | 0,866 | 0,936 | -0,001 (-0,011;0,01) |
| DG(36:2) | 0,172 | 0,864 | 0,936 | 0,001 (-0,009;0,011) |
| TG(45:0) | -0,154 | 0,878 | 0,946 | -0,001 (-0,011;0,009) |
| L-Alanine | 0,13 | 0,897 | 0,95 | 0,001 (-0,01;0,012) |
| Methylmalonylcarnitine | 0,138 | 0,89 | 0,95 | 0,001 (-0,009;0,011) |
| 9,10,13-TriHOME | 0,132 | 0,895 | 0,95 | 0,001 (-0,01;0,011) |
| CE(20:4) | 0,132 | 0,895 | 0,95 | 0,001 (-0,011;0,013) |
| TG(50:4) | -0,137 | 0,891 | 0,95 | -0,001 (-0,012;0,01) |
| Cer(d18:1/22:0) | 0,118 | 0,906 | 0,957 | 0,001 (-0,01;0,011) |
| Gamma-aminobutyric acid | -0,093 | 0,926 | 0,968 | 0 (-0,009;0,008) |
| N6,N6,N6-Trimethyl-L-lysine | -0,098 | 0,922 | 0,968 | -0,001 (-0,011;0,01) |
| Taurodeoxycholic acid | 0,089 | 0,929 | 0,968 | 0 (-0,01;0,011) |
| Lysophosphatidylcholine(22:6) | 0,088 | 0,93 | 0,968 | 0,001 (-0,011;0,012) |
| Phosphatidylcholine(O-34:2) | 0,078 | 0,938 | 0,969 | 0 (-0,011;0,011) |
| TG(50:3) | -0,078 | 0,938 | 0,969 | 0 (-0,011;0,01) |
| TG(54:7) | 0,075 | 0,94 | 0,969 | 0 (-0,011;0,012) |
| L-Glutamine | -0,07 | 0,944 | 0,97 | 0 (-0,011;0,011) |
| Phosphatidylcholine(34:3) | 0,066 | 0,948 | 0,97 | 0 (-0,011;0,011) |
| Cer(d18:1/24:1) | 0,048 | 0,962 | 0,981 | 0 (-0,011;0,011) |
| Lysophosphatidylethanolamine (16:1) | 0,043 | 0,966 | 0,982 | 0 (-0,01;0,011) |
| L-Lysine | -0,022 | 0,983 | 0,992 | 0 (-0,012;0,011) |
| Hydroxybutyric acid | 0,026 | 0,98 | 0,992 | 0 (-0,011;0,011) |
| Tetradecenoylcarnitine | 0,004 | 0,997 | 0,997 | 0 (-0,011;0,011) |
| Glycocholic acid | -0,011 | 0,991 | 0,997 | 0 (-0,01;0,01) |
| Phosphatidylethanolamine(O-38:5) | -0,007 | 0,995 | 0,997 | 0 (-0,012;0,012) |

**Supplementary Table 4. Metabolites differently expressed in all IPD patients (n = 140) compared to healthy controls (n = 64).** Shown are the results of the fixed effect meta-analysis: regression coefficient (CI: 95% confidence interval), nominal and adjusted p-values.

| **Name** | **z-value** | **Nominal *p*** | **Adjusted *p*** | **Beta (CI)** |
| --- | --- | --- | --- | --- |
| 3-Methoxytyrosine | 13,376 | 8,38E-41 | 2,55E-38 | 1,514 (1,292;1,736) |
| Methyldopa | 8,624 | 6,45E-18 | 9,8E-16 | 1,179 (0,911;1,447) |
| Putrescine | 4,321 | 0,0000155 | 0,00157 | 0,5 (0,273;0,726) |
| (+/-) 16-HDoHE | 3,906 | 0,000094 | 0,00714 | 0,629 (0,313;0,945) |
| Ornithine | 3,6 | 0,000318 | 0,0194 | 0,516 (0,235;0,797) |
| L-Threonine | 2,897 | 0,00377 | 0,191 | 0,464 (0,15;0,779) |
| TG(54:7) | -2,701 | 0,00692 | 0,301 | -0,466 (-0,805;-0,128) |
| 1-AG/2-AG | -2,589 | 0,00963 | 0,366 | -0,413 (-0,726;-0,1) |
| TG(54:1) | -2,476 | 0,0133 | 0,378 | -0,427 (-0,764;-0,089) |
| N6,N6,N6-Trimethyl-L-lysine | 2,472 | 0,0134 | 0,378 | 0,378 (0,078;0,677) |
| Tiglylcarnitine | -2,465 | 0,0137 | 0,378 | -0,406 (-0,728;-0,083) |
| Homocysteine | 2,372 | 0,0177 | 0,399 | 0,353 (0,061;0,644) |
| TG(50:4) | -2,368 | 0,0179 | 0,399 | -0,4 (-0,73;-0,069) |
| 2-ketoglutaric acid | -2,317 | 0,0205 | 0,399 | -0,362 (-0,669;-0,056) |
| Phosphatidylcholine(38:4) | -2,287 | 0,0222 | 0,399 | -0,405 (-0,751;-0,058) |
| Phosphatidylcholine(O-36:5) | -2,252 | 0,0243 | 0,399 | -0,395 (-0,739;-0,051) |
| Methylmalonylcarnitine | -2,227 | 0,026 | 0,399 | -0,341 (-0,641;-0,041) |
| Gamma-Glutamylglutamine | 2,211 | 0,027 | 0,399 | 0,35 (0,04;0,661) |
| TG(54:5) | -2,208 | 0,0273 | 0,399 | -0,386 (-0,729;-0,043) |
| Phosphatidylethanolamine(O-38:5) | -2,173 | 0,0298 | 0,399 | -0,396 (-0,753;-0,039) |
| Phosphatidylcholine(38:7) | -2,171 | 0,0299 | 0,399 | -0,378 (-0,719;-0,037) |
| Cystathionine | 2,131 | 0,0331 | 0,399 | 0,286 (0,023;0,549) |
| TG(52:4) | -2,124 | 0,0337 | 0,399 | -0,376 (-0,724;-0,029) |
| Phosphatidylcholine(36:4) | -2,112 | 0,0347 | 0,399 | -0,369 (-0,712;-0,027) |
| TG(48:3) | -2,088 | 0,0368 | 0,399 | -0,334 (-0,648;-0,02) |
| Glycoursodeoxycholic acid | -2,084 | 0,0371 | 0,399 | -0,281 (-0,545;-0,017) |
| Phosphatidylethanolamine(O-36:5) | -2,063 | 0,0391 | 0,399 | -0,372 (-0,726;-0,019) |
| TG(51:4) | -2,063 | 0,0391 | 0,399 | -0,346 (-0,674;-0,017) |
| TG(56:6) | -2,061 | 0,0393 | 0,399 | -0,362 (-0,706;-0,018) |
| Glycochenodeoxycholic acid | -2,06 | 0,0394 | 0,399 | -0,308 (-0,6;-0,015) |
| Cer(d18:0/22:0) | -2,047 | 0,0407 | 0,399 | -0,352 (-0,69;-0,015) |
| 1-LG/2-LG | -1,946 | 0,0517 | 0,466 | -0,307 (-0,615;0,002) |
| Saccharopine | 1,934 | 0,0531 | 0,466 | 0,312 (-0,004;0,629) |
| CE(20:4) | -1,929 | 0,0537 | 0,466 | -0,351 (-0,708;0,006) |
| OA01 - 2-hydroxybutyric acid | -1,917 | 0,0552 | 0,466 | -0,317 (-0,642;0,007) |
| Glycine | 1,9 | 0,0574 | 0,466 | 0,297 (-0,009;0,604) |
| TG(51:3) | -1,878 | 0,0604 | 0,466 | -0,309 (-0,633;0,014) |
| Phosphatidylcholine(O-38:5) | -1,877 | 0,0605 | 0,466 | -0,346 (-0,706;0,015) |
| TG(50:3) | -1,873 | 0,0611 | 0,466 | -0,315 (-0,644;0,015) |
| Phosphatidylcholine(40:5) | -1,871 | 0,0613 | 0,466 | -0,32 (-0,655;0,015) |
| TG(51:2) | -1,852 | 0,064 | 0,474 | -0,3 (-0,617;0,017) |
| TG(52:1) | -1,832 | 0,067 | 0,476 | -0,297 (-0,615;0,021) |
| L-Tyrosine | 1,819 | 0,069 | 0,476 | 0,281 (-0,022;0,584) |
| TG(48:2) | -1,818 | 0,069 | 0,476 | -0,291 (-0,604;0,023) |
| TG(56:7) | -1,807 | 0,0708 | 0,476 | -0,325 (-0,677;0,028) |
| TG(52:3) | -1,799 | 0,072 | 0,476 | -0,315 (-0,658;0,028) |
| TG(48:1) | -1,785 | 0,0743 | 0,481 | -0,285 (-0,599;0,028) |
| Phosphatidylcholine(O-36:4) | -1,749 | 0,0803 | 0,509 | -0,314 (-0,667;0,038) |
| TG(58:2) | -1,709 | 0,0875 | 0,541 | -0,305 (-0,655;0,045) |
| Glycocholic acid | -1,69 | 0,0911 | 0,541 | -0,256 (-0,554;0,041) |
| Lysophosphatidic acid 16(0) | -1,689 | 0,0912 | 0,541 | -0,243 (-0,525;0,039) |
| Isobutyrylcarnitine | -1,682 | 0,0925 | 0,541 | -0,271 (-0,588;0,045) |
| TG(51:1) | -1,644 | 0,1 | 0,574 | -0,259 (-0,568;0,05) |
| L-2-aminoadipic acid | -1,611 | 0,107 | 0,577 | -0,242 (-0,536;0,052) |
| TG(50:1) | -1,607 | 0,108 | 0,577 | -0,261 (-0,58;0,057) |
| Sphingomyelin(d18:1/15:0) | -1,596 | 0,111 | 0,577 | -0,265 (-0,591;0,061) |
| 9-HOTrE | 1,585 | 0,113 | 0,577 | 0,26 (-0,062;0,582) |
| Phosphatidylcholine(34:4) | -1,585 | 0,113 | 0,577 | -0,262 (-0,585;0,062) |
| Phosphatidylcholine(38:5) | -1,577 | 0,115 | 0,577 | -0,272 (-0,611;0,066) |
| TG(54:4) | -1,575 | 0,115 | 0,577 | -0,279 (-0,627;0,068) |
| Sphingomyelin(d18:1/18:1) | 1,555 | 0,12 | 0,577 | 0,246 (-0,064;0,557) |
| Phosphatidylethanolamine(38:4) | -1,55 | 0,121 | 0,577 | -0,251 (-0,569;0,066) |
| Lysophosphatidic acid 14(0) | -1,541 | 0,123 | 0,577 | -0,218 (-0,496;0,059) |
| Phosphatidylcholine(O-38:6) | -1,542 | 0,123 | 0,577 | -0,272 (-0,618;0,074) |
| Sphingomyelin(d18:1/20:1) | 1,544 | 0,123 | 0,577 | 0,264 (-0,071;0,599) |
| Sphingomyelin(d18:1/23:0) | -1,532 | 0,126 | 0,577 | -0,262 (-0,596;0,073) |
| Deoxycarnitine | -1,522 | 0,128 | 0,577 | -0,183 (-0,418;0,053) |
| TG(50:2) | -1,518 | 0,129 | 0,577 | -0,244 (-0,559;0,071) |
| Citric acid | 1,498 | 0,134 | 0,591 | 0,217 (-0,067;0,501) |
| TG(52:2) | -1,45 | 0,147 | 0,622 | -0,237 (-0,558;0,083) |
| L-Homoserine | 1,447 | 0,148 | 0,622 | 0,241 (-0,085;0,567) |
| TG(46:2) | -1,44 | 0,15 | 0,622 | -0,211 (-0,498;0,076) |
| TG(54:2) | -1,426 | 0,154 | 0,622 | -0,23 (-0,545;0,086) |
| TG(56:1) | -1,425 | 0,154 | 0,622 | -0,212 (-0,504;0,08) |
| Palmitoylcarnitine | 1,42 | 0,155 | 0,622 | 0,231 (-0,088;0,55) |
| TG(56:5) | -1,423 | 0,155 | 0,622 | -0,249 (-0,592;0,094) |
| DG(36:2) | -1,413 | 0,158 | 0,622 | -0,224 (-0,536;0,087) |
| Phosphatidylethanolamine(38:2) | -1,401 | 0,161 | 0,626 | -0,23 (-0,552;0,092) |
| TG(60:3) | -1,396 | 0,163 | 0,626 | -0,246 (-0,592;0,099) |
| Choline | -1,381 | 0,167 | 0,629 | -0,222 (-0,536;0,093) |
| Hexadecenoylcarnitine | 1,377 | 0,169 | 0,629 | 0,222 (-0,094;0,539) |
| Sphingomyelin(d18:1/18:2) | -1,373 | 0,17 | 0,629 | -0,269 (-0,654;0,115) |
| Gamma-aminobutyric acid | -1,358 | 0,174 | 0,637 | -0,188 (-0,46;0,083) |
| TG(57:1) | -1,353 | 0,176 | 0,637 | -0,229 (-0,561;0,103) |
| Phosphatidylcholine(40:7) | -1,313 | 0,189 | 0,655 | -0,212 (-0,528;0,104) |
| TG(42:0) | -1,312 | 0,19 | 0,655 | -0,187 (-0,467;0,092) |
| FA 22(5) w6 | -1,306 | 0,192 | 0,655 | -0,209 (-0,523;0,105) |
| Phosphatidylcholine(38:2) | -1,3 | 0,194 | 0,655 | -0,22 (-0,552;0,112) |
| Phosphatidylcholine(38:3) | -1,295 | 0,195 | 0,655 | -0,212 (-0,533;0,109) |
| 3-Hydroxypropionic Acid | -1,293 | 0,196 | 0,655 | -0,179 (-0,451;0,093) |
| TG(44:1) | -1,294 | 0,196 | 0,655 | -0,186 (-0,468;0,096) |
| Thromboxane B2 | -1,283 | 0,199 | 0,657 | -0,186 (-0,469;0,098) |
| Serotonine | -1,277 | 0,202 | 0,657 | -0,127 (-0,322;0,068) |
| TG(46:1) | -1,272 | 0,203 | 0,657 | -0,194 (-0,494;0,105) |
| Phosphatidylethanolamine(O-38:7) | -1,266 | 0,205 | 0,657 | -0,219 (-0,557;0,12) |
| 15(S)-HETrE | 1,228 | 0,219 | 0,69 | 0,159 (-0,094;0,412) |
| TG(54:3) | -1,208 | 0,227 | 0,69 | -0,206 (-0,541;0,128) |
| TG(58:9) | -1,2 | 0,23 | 0,69 | -0,203 (-0,533;0,128) |
| Phosphatidylcholine(36:3) | -1,177 | 0,239 | 0,69 | -0,195 (-0,52;0,13) |
| Ethanolamine | 1,172 | 0,241 | 0,69 | 0,176 (-0,118;0,47) |
| Cer(d18:1/23:0) | -1,172 | 0,241 | 0,69 | -0,196 (-0,525;0,132) |
| Phosphatidylcholine(32:0) | -1,168 | 0,243 | 0,69 | -0,202 (-0,541;0,137) |
| Methionine sulfone | -1,162 | 0,245 | 0,69 | -0,19 (-0,511;0,131) |
| Sphingosine 1-phosphate 18(2) | 1,159 | 0,247 | 0,69 | 0,115 (-0,079;0,308) |
| Hexanoylcarnitine | 1,155 | 0,248 | 0,69 | 0,193 (-0,135;0,52) |
| Sphingomyelin(d18:1/18:0) | 1,152 | 0,249 | 0,69 | 0,186 (-0,131;0,503) |
| Isocitrate | -1,149 | 0,251 | 0,69 | -0,149 (-0,404;0,105) |
| 8(9)-EpETrE | 1,138 | 0,255 | 0,69 | 0,105 (-0,076;0,287) |
| TG(42:1) | -1,139 | 0,255 | 0,69 | -0,172 (-0,469;0,124) |
| Lysophosphatidylinositol 16(1) | 1,133 | 0,257 | 0,69 | 0,183 (-0,133;0,499) |
| Lysophosphatidylinositol 18(1) | 1,134 | 0,257 | 0,69 | 0,192 (-0,139;0,523) |
| Taurocholic acid | -1,135 | 0,257 | 0,69 | -0,181 (-0,495;0,132) |
| O-Acetyl-L-serine | -1,13 | 0,259 | 0,69 | -0,172 (-0,47;0,126) |
| Phosphatidylcholine(40:6) | -1,129 | 0,259 | 0,69 | -0,187 (-0,511;0,137) |
| TG(60:1) | -1,113 | 0,266 | 0,696 | -0,192 (-0,531;0,146) |
| Cer(d18:1/22:0) | -1,103 | 0,27 | 0,696 | -0,176 (-0,49;0,137) |
| 17,18-DiHETE | 1,1 | 0,271 | 0,696 | 0,18 (-0,14;0,5) |
| Lysophosphatidylethanolamine (16:1) | 1,091 | 0,275 | 0,696 | 0,169 (-0,135;0,473) |
| Octanoylcarnitine | 1,074 | 0,283 | 0,696 | 0,178 (-0,147;0,504) |
| Lysophosphatidylcholine(O-18:1) | -1,065 | 0,287 | 0,696 | -0,172 (-0,487;0,144) |
| TG(44:2) | -1,064 | 0,287 | 0,696 | -0,159 (-0,451;0,133) |
| TG(55:1) | -1,063 | 0,288 | 0,696 | -0,198 (-0,562;0,167) |
| TG(42:2) | -1,052 | 0,293 | 0,696 | -0,165 (-0,473;0,142) |
| Lysophosphatidylinositol 20(4) | 1,049 | 0,294 | 0,696 | 0,166 (-0,145;0,477) |
| Phosphatidylethanolamine(36:4) | -1,05 | 0,294 | 0,696 | -0,168 (-0,482;0,146) |
| Sphingomyelin(d18:1/23:1) | -1,049 | 0,294 | 0,696 | -0,173 (-0,495;0,15) |
| Decanoylcarnitine | 1,048 | 0,295 | 0,696 | 0,171 (-0,149;0,491) |
| Phosphatidylcholine(38:6) | -1,047 | 0,295 | 0,696 | -0,173 (-0,497;0,151) |
| TG(60:2) | -1,036 | 0,3 | 0,696 | -0,183 (-0,528;0,163) |
| 11-HETE | 1,031 | 0,303 | 0,696 | 0,153 (-0,138;0,444) |
| Nonaylcarnitine | -1,026 | 0,305 | 0,696 | -0,165 (-0,479;0,15) |
| 12,13-DiHOME | 1,026 | 0,305 | 0,696 | 0,167 (-0,152;0,486) |
| 2-Methylbutyroylcarnitine | -1,02 | 0,308 | 0,696 | -0,153 (-0,446;0,141) |
| Phosphatidylcholine(34:3) | -1,017 | 0,309 | 0,696 | -0,173 (-0,507;0,161) |
| Tetradecenoylcarnitine | 1,012 | 0,311 | 0,696 | 0,167 (-0,157;0,492) |
| Phosphatidylethanolamine(34:2) | -1,013 | 0,311 | 0,696 | -0,171 (-0,503;0,16) |
| Propionylcarnitine | -1 | 0,317 | 0,7 | -0,15 (-0,444;0,144) |
| FA 22(6) | -0,998 | 0,318 | 0,7 | -0,161 (-0,477;0,155) |
| FA 18(1) | 0,992 | 0,321 | 0,7 | 0,162 (-0,159;0,483) |
| 1-Methylhistidine | -0,985 | 0,325 | 0,7 | -0,156 (-0,467;0,155) |
| Dodecenoylcarnitine | 0,985 | 0,325 | 0,7 | 0,159 (-0,157;0,475) |
| TG(44:0) | -0,981 | 0,327 | 0,7 | -0,137 (-0,41;0,137) |
| Myristoilcarnitine | 0,968 | 0,333 | 0,705 | 0,156 (-0,16;0,473) |
| Lysophosphatidylethanolamine(18:0) | -0,967 | 0,334 | 0,705 | -0,163 (-0,494;0,168) |
| Sphingomyelin(d18:1/21:0) | -0,95 | 0,342 | 0,711 | -0,164 (-0,501;0,174) |
| TG(56:3) | -0,947 | 0,344 | 0,711 | -0,157 (-0,483;0,168) |
| Oleoylcarnitine | 0,941 | 0,347 | 0,711 | 0,153 (-0,166;0,472) |
| Pyroglutamic acid | -0,937 | 0,349 | 0,711 | -0,158 (-0,487;0,172) |
| LPS 18(1) | -0,937 | 0,349 | 0,711 | -0,092 (-0,284;0,1) |
| 11,12-DiHETrE | -0,927 | 0,354 | 0,718 | -0,147 (-0,458;0,164) |
| Deoxycholic acid | 0,918 | 0,359 | 0,722 | 0,154 (-0,174;0,482) |
| DGLEA | -0,909 | 0,363 | 0,722 | -0,149 (-0,47;0,172) |
| TG(46:0) | -0,909 | 0,363 | 0,722 | -0,137 (-0,433;0,159) |
| DG(36:3) | -0,905 | 0,366 | 0,722 | -0,145 (-0,46;0,169) |
| Phosphatidylcholine(40:8) | -0,894 | 0,371 | 0,725 | -0,129 (-0,411;0,153) |
| TG(53:1) | -0,891 | 0,373 | 0,725 | -0,141 (-0,45;0,169) |
| Homocitrulline | -0,888 | 0,374 | 0,725 | -0,148 (-0,475;0,179) |
| 12,13-EpOME | 0,863 | 0,388 | 0,741 | 0,154 (-0,195;0,503) |
| Trimethylamine N-oxide | 0,857 | 0,391 | 0,741 | 0,143 (-0,185;0,472) |
| 9,10-DiHOME | 0,855 | 0,393 | 0,741 | 0,131 (-0,169;0,43) |
| TG(56:4) | -0,852 | 0,394 | 0,741 | -0,148 (-0,488;0,192) |
| Sphingosine 1-phosphate 18(0) | -0,845 | 0,398 | 0,741 | -0,06 (-0,199;0,079) |
| Glycylglycine | 0,841 | 0,4 | 0,741 | 0,134 (-0,178;0,445) |
| 8,12-iPF2a IV | -0,842 | 0,4 | 0,741 | -0,119 (-0,397;0,159) |
| Betaine | -0,838 | 0,402 | 0,741 | -0,129 (-0,432;0,173) |
| Lysophosphatidylcholine(O-16:0) | -0,823 | 0,411 | 0,748 | -0,142 (-0,482;0,197) |
| Lysophosphatidylcholine(O-16:1) | -0,822 | 0,411 | 0,748 | -0,145 (-0,491;0,201) |
| Lauroylcarnitine | 0,812 | 0,417 | 0,751 | 0,134 (-0,189;0,457) |
| Cer(d18:0/23:0) | -0,806 | 0,42 | 0,751 | -0,138 (-0,474;0,198) |
| Sphingomyelin(d18:1/16:0) | -0,806 | 0,42 | 0,751 | -0,147 (-0,504;0,21) |
| Phosphatidylcholine(O-44:5) | 0,8 | 0,424 | 0,751 | 0,142 (-0,206;0,491) |
| Lysophosphatidylcholine(16:1) | 0,794 | 0,427 | 0,751 | 0,136 (-0,199;0,471) |
| Phosphatidylethanolamine(38:6) | -0,793 | 0,428 | 0,751 | -0,125 (-0,432;0,183) |
| L-Kynurenine | -0,783 | 0,433 | 0,755 | -0,131 (-0,457;0,196) |
| 9,12,13-TriHOME | 0,781 | 0,435 | 0,755 | 0,122 (-0,185;0,43) |
| Phosphatidylcholine(36:5) | -0,777 | 0,437 | 0,755 | -0,129 (-0,454;0,196) |
| Lysophosphatidylcholine(16:0) | -0,762 | 0,446 | 0,761 | -0,133 (-0,475;0,209) |
| Phosphatidylcholine(32:2) | -0,756 | 0,45 | 0,761 | -0,13 (-0,466;0,206) |
| 5,6-DiHETrE | -0,752 | 0,452 | 0,761 | -0,121 (-0,435;0,194) |
| Stearoylcarnitine | 0,743 | 0,457 | 0,761 | 0,114 (-0,186;0,414) |
| Taurodeoxycholic acid | -0,744 | 0,457 | 0,761 | -0,117 (-0,426;0,191) |
| 9,10,13-TriHOME | 0,74 | 0,459 | 0,761 | 0,114 (-0,187;0,414) |
| Lysophosphatidylcholine(18:0) | -0,734 | 0,463 | 0,761 | -0,131 (-0,48;0,218) |
| Prostaglandin E2 | -0,73 | 0,465 | 0,761 | -0,108 (-0,399;0,182) |
| Phosphatidylcholine(36:2) | -0,731 | 0,465 | 0,761 | -0,125 (-0,46;0,21) |
| Lysophosphatidylcholine(20:4) | -0,729 | 0,466 | 0,761 | -0,123 (-0,455;0,208) |
| Lysophosphatidylethanolamine(18:1) | 0,717 | 0,473 | 0,769 | 0,105 (-0,182;0,391) |
| Sarcosine | 0,712 | 0,477 | 0,771 | 0,106 (-0,186;0,399) |
| S-Methylcysteine | -0,705 | 0,481 | 0,774 | -0,116 (-0,439;0,207) |
| Lysophosphatidylinositol 18(2) | 0,7 | 0,484 | 0,774 | 0,116 (-0,209;0,441) |
| TG(56:2) | -0,688 | 0,491 | 0,779 | -0,107 (-0,413;0,198) |
| L-4-hydroxy-proline | 0,681 | 0,496 | 0,779 | 0,108 (-0,203;0,42) |
| L-Methionine | -0,68 | 0,496 | 0,779 | -0,099 (-0,385;0,186) |
| TG(58:8) | -0,679 | 0,497 | 0,779 | -0,119 (-0,461;0,224) |
| L-Glutamine | 0,67 | 0,503 | 0,784 | 0,11 (-0,212;0,432) |
| Citrulline | -0,664 | 0,507 | 0,786 | -0,11 (-0,435;0,215) |
| CE(18:2) | -0,654 | 0,513 | 0,792 | -0,111 (-0,445;0,222) |
| Lysophosphatidylcholine(14:0) | -0,629 | 0,529 | 0,809 | -0,111 (-0,456;0,235) |
| Sphingomyelin(d18:1/25:1) | -0,624 | 0,532 | 0,809 | -0,103 (-0,427;0,221) |
| TG(56:0) | -0,624 | 0,532 | 0,809 | -0,106 (-0,439;0,227) |
| 8,9-DiHETrE | -0,603 | 0,546 | 0,826 | -0,093 (-0,393;0,208) |
| Lysophosphatidylethanolamine (20:4) | -0,592 | 0,554 | 0,834 | -0,095 (-0,412;0,221) |
| (+/-) 8-HDoHE | 0,587 | 0,557 | 0,835 | 0,103 (-0,241;0,446) |
| Lysophosphatidylcholine(18:1) | 0,581 | 0,561 | 0,836 | 0,101 (-0,238;0,439) |
| Linoleoyl ethanolamide | -0,574 | 0,566 | 0,839 | -0,094 (-0,413;0,226) |
| Cholic acid | -0,559 | 0,576 | 0,85 | -0,09 (-0,404;0,225) |
| Acetylcarnitine | -0,555 | 0,579 | 0,85 | -0,091 (-0,413;0,231) |
| Taurolithocholic acid | -0,55 | 0,583 | 0,852 | -0,092 (-0,419;0,235) |
| Taurochenodeoxycholic acid | 0,544 | 0,586 | 0,853 | 0,088 (-0,23;0,406) |
| Dehydroepiandrosteron | -0,531 | 0,595 | 0,862 | -0,086 (-0,405;0,232) |
| Sphingomyelin(d18:1/14:0) | -0,525 | 0,6 | 0,863 | -0,091 (-0,429;0,248) |
| L-Glutamic acid | -0,522 | 0,602 | 0,863 | -0,08 (-0,38;0,22) |
| L-Alpha-aminobutyric acid | -0,502 | 0,615 | 0,873 | -0,082 (-0,404;0,239) |
| L-Leucine | -0,5 | 0,617 | 0,873 | -0,075 (-0,369;0,219) |
| L-Tryptophan | -0,495 | 0,621 | 0,873 | -0,078 (-0,386;0,231) |
| 9-HODE | 0,494 | 0,621 | 0,873 | 0,075 (-0,223;0,374) |
| Glutathione | -0,487 | 0,626 | 0,873 | -0,065 (-0,327;0,197) |
| Palmitoyl ethanolamide | 0,487 | 0,626 | 0,873 | 0,083 (-0,251;0,417) |
| Phosphatidylcholine(O-34:2) | 0,481 | 0,631 | 0,874 | 0,079 (-0,242;0,399) |
| Phosphatidylcholine(32:1) | 0,474 | 0,636 | 0,874 | 0,076 (-0,238;0,391) |
| FA 18(2) | -0,466 | 0,641 | 0,874 | -0,068 (-0,354;0,218) |
| Phosphatidylcholine(34:2) | -0,465 | 0,642 | 0,874 | -0,08 (-0,419;0,258) |
| Prostaglandin F2α | 0,458 | 0,647 | 0,874 | 0,061 (-0,201;0,324) |
| Lactic acid | -0,455 | 0,649 | 0,874 | -0,073 (-0,389;0,242) |
| 13-HODE | 0,454 | 0,65 | 0,874 | 0,061 (-0,204;0,327) |
| 19,20-DiHDPA | 0,454 | 0,65 | 0,874 | 0,073 (-0,242;0,387) |
| L-Alanine | -0,447 | 0,655 | 0,877 | -0,075 (-0,403;0,254) |
| L-Histidine | -0,443 | 0,658 | 0,877 | -0,075 (-0,405;0,255) |
| Sphingomyelin(d18:1/24:1) | -0,42 | 0,675 | 0,895 | -0,075 (-0,423;0,273) |
| Sphingomyelin(d18:1/16:1) | -0,411 | 0,681 | 0,896 | -0,071 (-0,412;0,269) |
| 5-HETE | 0,41 | 0,682 | 0,896 | 0,066 (-0,249;0,38) |
| Gamma-L-glutamyl-L-alanine | 0,404 | 0,686 | 0,896 | 0,065 (-0,251;0,382) |
| Lysophosphatidylcholine(18:2) | 0,404 | 0,687 | 0,896 | 0,069 (-0,267;0,405) |
| Cortisol | -0,397 | 0,692 | 0,899 | -0,061 (-0,365;0,242) |
| Sphingomyelin(d18:1/25:0) | -0,391 | 0,696 | 0,9 | -0,07 (-0,42;0,281) |
| 12,13-DiHODE | -0,381 | 0,703 | 0,905 | -0,061 (-0,377;0,254) |
| Sphingomyelin(d18:1/20:0) | 0,374 | 0,708 | 0,908 | 0,064 (-0,271;0,398) |
| Carnitine | 0,369 | 0,712 | 0,91 | 0,058 (-0,248;0,363) |
| L-Lysine | 0,356 | 0,722 | 0,915 | 0,06 (-0,27;0,39) |
| Lysophosphatidylethanolamine(22:6) | -0,353 | 0,724 | 0,915 | -0,058 (-0,381;0,265) |
| Lysophosphatidic acid 18(1) | 0,351 | 0,726 | 0,915 | 0,053 (-0,244;0,35) |
| 3-Hydroxybutyric acid | -0,347 | 0,729 | 0,915 | -0,06 (-0,4;0,28) |
| Lysophosphatidic acid 18(2) | -0,344 | 0,731 | 0,915 | -0,054 (-0,36;0,253) |
| Phosphatidylcholine(O-34:3) | -0,335 | 0,738 | 0,917 | -0,057 (-0,39;0,276) |
| Lysophosphatidylethanolamine (20:5) | 0,333 | 0,739 | 0,917 | 0,054 (-0,264;0,372) |
| CE(18:1) | -0,329 | 0,742 | 0,917 | -0,057 (-0,396;0,282) |
| Taurine | -0,316 | 0,752 | 0,925 | -0,026 (-0,189;0,137) |
| Cer(d18:1/24:1) | -0,31 | 0,756 | 0,927 | -0,055 (-0,401;0,291) |
| 14,15-DiHETE | 0,306 | 0,76 | 0,928 | 0,05 (-0,273;0,374) |
| Sphingomyelin(d18:1/22:0) | -0,296 | 0,767 | 0,931 | -0,05 (-0,384;0,283) |
| Phosphatidylcholine(36:1) | 0,294 | 0,769 | 0,931 | 0,046 (-0,262;0,355) |
| L-Valine | -0,275 | 0,783 | 0,941 | -0,044 (-0,358;0,27) |
| Tetradecadienylcarnitine | 0,275 | 0,783 | 0,941 | 0,045 (-0,275;0,364) |
| Decenoylcarnitine | 0,266 | 0,79 | 0,943 | 0,044 (-0,28;0,368) |
| TG(54:0) | -0,262 | 0,793 | 0,943 | -0,048 (-0,41;0,313) |
| CE(22:6) | -0,257 | 0,797 | 0,943 | -0,041 (-0,357;0,274) |
| L-Asparagine | -0,256 | 0,798 | 0,943 | -0,04 (-0,347;0,267) |
| Cysteine | -0,253 | 0,801 | 0,943 | -0,037 (-0,322;0,248) |
| Malic acid | -0,237 | 0,813 | 0,954 | -0,035 (-0,327;0,256) |
| Lysophosphatidylcholine(20:5) | 0,224 | 0,823 | 0,955 | 0,033 (-0,253;0,318) |
| Symmetric dimethylarginine | 0,218 | 0,828 | 0,955 | 0,035 (-0,276;0,345) |
| Sphingomyelin(d18:1/24:0) | -0,212 | 0,832 | 0,955 | -0,037 (-0,374;0,301) |
| Valerylcarnitine | -0,205 | 0,837 | 0,955 | -0,033 (-0,346;0,28) |
| L-Phenylalanine | -0,205 | 0,838 | 0,955 | -0,032 (-0,339;0,275) |
| Glycolithocholic acid | -0,203 | 0,839 | 0,955 | -0,034 (-0,361;0,294) |
| Lysophosphatidylethanolamine(20:4) | -0,198 | 0,843 | 0,955 | -0,034 (-0,37;0,302) |
| L-Proline | -0,196 | 0,844 | 0,955 | -0,031 (-0,339;0,277) |
| 12-HETE | 0,196 | 0,845 | 0,955 | 0,014 (-0,125;0,153) |
| DL-3-aminoisobutyric acid | 0,193 | 0,847 | 0,955 | 0,031 (-0,284;0,346) |
| Sphingomyelin(d18:1/24:2) | 0,188 | 0,851 | 0,955 | 0,033 (-0,31;0,376) |
| L-Serine | 0,18 | 0,857 | 0,955 | 0,029 (-0,29;0,349) |
| Lysophosphatidylcholine(22:6) | 0,177 | 0,859 | 0,955 | 0,03 (-0,307;0,367) |
| Hydroxylysine | -0,172 | 0,863 | 0,955 | -0,029 (-0,361;0,302) |
| Glycodeoxycholic acid | -0,17 | 0,865 | 0,955 | -0,028 (-0,356;0,3) |
| Phosphatidylcholine(O-38:4) | -0,167 | 0,867 | 0,955 | -0,028 (-0,358;0,302) |
| Butyrylcarnitine | -0,167 | 0,868 | 0,955 | -0,027 (-0,349;0,294) |
| O-Phosphoethanolamine | -0,163 | 0,87 | 0,955 | -0,01 (-0,136;0,115) |
| Phosphatidylcholine(O-34:1) | 0,15 | 0,881 | 0,963 | 0,025 (-0,301;0,351) |
| Isovalerylcarnitine | -0,133 | 0,894 | 0,974 | -0,02 (-0,314;0,274) |
| Phosphatidylcholine(34:1) | 0,126 | 0,9 | 0,977 | 0,005 (-0,08;0,091) |
| O-Anandamide | -0,11 | 0,913 | 0,983 | -0,019 (-0,35;0,313) |
| Pimelylcarnitine | -0,104 | 0,917 | 0,983 | -0,016 (-0,325;0,293) |
| Linoleylcarnitine | -0,101 | 0,92 | 0,983 | -0,014 (-0,284;0,256) |
| Lysophosphatidylcholine(20:3) | 0,097 | 0,923 | 0,983 | 0,016 (-0,313;0,346) |
| Phosphatidylcholine(O-36:3) | 0,095 | 0,924 | 0,983 | 0,014 (-0,285;0,314) |
| TG(45:0) | -0,095 | 0,925 | 0,983 | -0,015 (-0,332;0,301) |
| 14,15-DiHETrE | -0,087 | 0,931 | 0,986 | -0,014 (-0,33;0,302) |
| 8-HETE | -0,08 | 0,936 | 0,988 | -0,01 (-0,244;0,225) |
| Octenoylcarnitine | 0,067 | 0,946 | 0,99 | 0,011 (-0,316;0,339) |
| Malonylcarnitine | -0,063 | 0,949 | 0,99 | -0,01 (-0,326;0,306) |
| L-Arginine | 0,062 | 0,95 | 0,99 | 0,01 (-0,309;0,33) |
| Lysophosphatidylethanolamine (16:0) | 0,06 | 0,952 | 0,99 | 0,01 (-0,313;0,333) |
| Sphingosine 1-phosphate 18(1) | 0,057 | 0,954 | 0,99 | 0,004 (-0,131;0,138) |
| (+/-) 11-HDoHE | -0,043 | 0,966 | 0,993 | -0,004 (-0,194;0,186) |
| TG(59:1) | -0,04 | 0,968 | 0,993 | -0,007 (-0,355;0,34) |
| Lysophosphatidic acid 16(1) | -0,039 | 0,969 | 0,993 | -0,005 (-0,279;0,268) |
| (+/-) 10-HDoHE | 0,037 | 0,971 | 0,993 | 0,003 (-0,179;0,186) |
| Anandamide | 0,03 | 0,976 | 0,995 | 0,005 (-0,324;0,334) |
| Lysophosphatidylinositol 18(0) | -0,026 | 0,979 | 0,995 | -0,004 (-0,297;0,289) |
| L-Isoleucine | -0,008 | 0,994 | 1 | -0,001 (-0,304;0,301) |
| (+/-) 14-HDoHE | -0,008 | 0,994 | 1 | -0,001 (-0,127;0,126) |
| Lysophosphatidylcholine(18:3) | 0,005 | 0,996 | 1 | 0,001 (-0,361;0,362) |
| 12(S)-HEPE | -0,002 | 0,998 | 1 | 0 (-0,134;0,134) |
| Sphingomyelin(d18:1/22:1) | 0 | 1 | 1 | 0 (-0,337;0,337) |

**Supplementary Table 5. Metabolites differently expressed in IPD patients with L-Dopa treatment (Dopa^positive^, n = 110) compared to controls (n = 64).** Shown are the results of the fixed effect meta-analysis: regression coefficient (CI: 95% confidence interval), nominal and adjusted p-values.

| **Name** | **z-value** | **Nominal *p*** | **Adjusted *p*** | **Beta (CI)** |
| --- | --- | --- | --- | --- |
| 3-Methoxytyrosine | 1,95 | 0 | 0 | 1,95 (1,873;2,026) |
| Methyldopa | 1,571 | 6.36e-40 | 9.67e-38 | 1,571 (1,339;1,804) |
| (+/-) 16-HDoHE | 0,809 | 2.24e-06 | 0,000227 | 0,809 (0,474;1,144) |
| Putrescine | 0,547 | 6.39e-06 | 0,000485 | 0,547 (0,309;0,784) |
| Ornithinie | 0,582 | 6.36e-05 | 0,00387 | 0,582 (0,297;0,868) |
| N6,N6,N6-Trimethyl-L-lysine | 0,493 | 0,00265 | 0,135 | 0,493 (0,171;0,814) |
| L-Threonine | 0,486 | 0,00366 | 0,159 | 0,486 (0,158;0,813) |
| Homocysteine | 0,403 | 0,00949 | 0,309 | 0,403 (0,099;0,708) |
| L-Tyrosine | 0,416 | 0,0102 | 0,309 | 0,416 (0,099;0,734) |
| 2-ketoglutaric acid | -0,424 | 0,00907 | 0,309 | -0,424 (-0,742;-0,105) |
| TG(54:7) | -0,446 | 0,0144 | 0,398 | -0,446 (-0,803;-0,089) |
| TG(54:1) | -0,39 | 0,0161 | 0,407 | -0,39 (-0,708;-0,073) |
| Methylmalonylcarnitine | -0,367 | 0,0176 | 0,41 | -0,367 (-0,67;-0,064) |
| Cystathionine | 0,324 | 0,0196 | 0,425 | 0,324 (0,052;0,597) |
| Gamma-Glutamylglutamine | 0,377 | 0,0232 | 0,47 | 0,377 (0,052;0,703) |
| Glycocholic acid | -0,321 | 0,0301 | 0,571 | -0,321 (-0,611;-0,031) |
| Glycochenodeoxycholic acid | -0,345 | 0,0319 | 0,571 | -0,345 (-0,66;-0,03) |
| Deoxycarnitine | -0,266 | 0,0346 | 0,584 | -0,266 (-0,513;-0,019) |
| Saccharopine | 0,354 | 0,0403 | 0,625 | 0,354 (0,016;0,692) |
| Hydroxybutyric acid | -0,361 | 0,0425 | 0,625 | -0,361 (-0,711;-0,012) |
| Glycoursodeoxycholic acid | -0,295 | 0,0473 | 0,625 | -0,295 (-0,587;-0,004) |
| Cer(d18:0/22:0) | -0,345 | 0,0444 | 0,625 | -0,345 (-0,68;-0,009) |
| Phosphatidylcholine(38:4) | -0,375 | 0,0456 | 0,625 | -0,375 (-0,742;-0,007) |
| Tiglylcarnitine | -0,336 | 0,0503 | 0,63 | -0,336 (-0,673;0) |
| TG(50:4) | -0,352 | 0,0518 | 0,63 | -0,352 (-0,708;0,003) |
| 1-Methylhistidine | -0,232 | 0,175 | 0,634 | -0,232 (-0,568;0,103) |
| Ethanolamine | 0,204 | 0,18 | 0,634 | 0,204 (-0,095;0,503) |
| Gamma-aminobutyric acid | -0,216 | 0,145 | 0,634 | -0,216 (-0,506;0,074) |
| Glycine | 0,322 | 0,061 | 0,634 | 0,322 (-0,015;0,659) |
| Glycylglycine | 0,222 | 0,172 | 0,634 | 0,222 (-0,096;0,54) |
| L-Homoserine | 0,249 | 0,172 | 0,634 | 0,249 (-0,108;0,606) |
| Methionine sulfone | -0,265 | 0,144 | 0,634 | -0,265 (-0,619;0,09) |
| Serotonine | -0,2 | 0,0703 | 0,634 | -0,2 (-0,417;0,017) |
| Betaine | -0,221 | 0,154 | 0,634 | -0,221 (-0,524;0,082) |
| Choline | -0,25 | 0,149 | 0,634 | -0,25 (-0,59;0,09) |
| 3-Hydroxypropionic Acid | -0,229 | 0,128 | 0,634 | -0,229 (-0,523;0,066) |
| FA 22(5) w6 | -0,268 | 0,119 | 0,634 | -0,268 (-0,605;0,069) |
| FA 22(6) | -0,256 | 0,121 | 0,634 | -0,256 (-0,58;0,068) |
| Lysophosphatidylethanolamine (16:1) | 0,249 | 0,123 | 0,634 | 0,249 (-0,067;0,566) |
| Lysophosphatidylinositol 16(1) | 0,25 | 0,156 | 0,634 | 0,25 (-0,095;0,595) |
| Sphingosine 1-phosphate 18(2) | 0,148 | 0,169 | 0,634 | 0,148 (-0,063;0,358) |
| Taurocholic acid | -0,21 | 0,188 | 0,634 | -0,21 (-0,522;0,102) |
| 1-AG/2-AG | -0,287 | 0,0936 | 0,634 | -0,287 (-0,623;0,049) |
| 11-HETE | 0,214 | 0,175 | 0,634 | 0,214 (-0,095;0,523) |
| 15(S)-HETrE | 0,199 | 0,147 | 0,634 | 0,199 (-0,07;0,467) |
| 9-HOTrE | 0,291 | 0,102 | 0,634 | 0,291 (-0,057;0,639) |
| Deoxycholic acid | 0,292 | 0,0838 | 0,634 | 0,292 (-0,039;0,623) |
| CE(20:4) | -0,325 | 0,0868 | 0,634 | -0,325 (-0,698;0,047) |
| Cer(d18:1/23:0) | -0,229 | 0,189 | 0,634 | -0,229 (-0,572;0,113) |
| Phosphatidylcholine(34:4) | -0,253 | 0,164 | 0,634 | -0,253 (-0,61;0,103) |
| Phosphatidylcholine(36:4) | -0,256 | 0,169 | 0,634 | -0,256 (-0,622;0,109) |
| Phosphatidylcholine(38:2) | -0,242 | 0,188 | 0,634 | -0,242 (-0,601;0,118) |
| Phosphatidylcholine(38:5) | -0,29 | 0,0929 | 0,634 | -0,29 (-0,628;0,048) |
| Phosphatidylcholine(38:6) | -0,218 | 0,176 | 0,634 | -0,218 (-0,534;0,098) |
| Phosphatidylcholine(38:7) | -0,299 | 0,111 | 0,634 | -0,299 (-0,666;0,069) |
| Phosphatidylcholine(40:5) | -0,303 | 0,0803 | 0,634 | -0,303 (-0,643;0,037) |
| Phosphatidylcholine(40:6) | -0,255 | 0,117 | 0,634 | -0,255 (-0,575;0,064) |
| Phosphatidylcholine(40:7) | -0,267 | 0,0997 | 0,634 | -0,267 (-0,585;0,051) |
| Phosphatidylcholine(O-36:4) | -0,329 | 0,0938 | 0,634 | -0,329 (-0,713;0,056) |
| Phosphatidylcholine(O-36:5) | -0,342 | 0,0744 | 0,634 | -0,342 (-0,717;0,034) |
| Phosphatidylcholine(O-38:5) | -0,369 | 0,0672 | 0,634 | -0,369 (-0,765;0,026) |
| Phosphatidylcholine(O-38:6) | -0,338 | 0,0627 | 0,634 | -0,338 (-0,694;0,018) |
| Phosphatidylethanolamine(O-36:5) | -0,3 | 0,115 | 0,634 | -0,3 (-0,674;0,074) |
| Phosphatidylethanolamine(O-38:5) | -0,328 | 0,0871 | 0,634 | -0,328 (-0,704;0,048) |
| Phosphatidylethanolamine(O-38:7) | -0,248 | 0,162 | 0,634 | -0,248 (-0,595;0,1) |
| Sphingomyelin(d18:1/15:0) | -0,279 | 0,121 | 0,634 | -0,279 (-0,632;0,074) |
| Sphingomyelin(d18:1/18:1) | 0,272 | 0,112 | 0,634 | 0,272 (-0,063;0,607) |
| Sphingomyelin(d18:1/18:2) | -0,321 | 0,124 | 0,634 | -0,321 (-0,729;0,088) |
| Sphingomyelin(d18:1/20:1) | 0,253 | 0,167 | 0,634 | 0,253 (-0,106;0,612) |
| Sphingomyelin(d18:1/23:0) | -0,26 | 0,151 | 0,634 | -0,26 (-0,616;0,095) |
| TG(46:2) | -0,222 | 0,174 | 0,634 | -0,222 (-0,541;0,098) |
| TG(48:1) | -0,27 | 0,122 | 0,634 | -0,27 (-0,612;0,072) |
| TG(48:2) | -0,268 | 0,124 | 0,634 | -0,268 (-0,61;0,073) |
| TG(48:3) | -0,331 | 0,0575 | 0,634 | -0,331 (-0,672;0,011) |
| TG(50:1) | -0,24 | 0,173 | 0,634 | -0,24 (-0,586;0,105) |
| TG(50:3) | -0,277 | 0,128 | 0,634 | -0,277 (-0,634;0,08) |
| TG(51:1) | -0,228 | 0,19 | 0,634 | -0,228 (-0,569;0,113) |
| TG(51:2) | -0,296 | 0,0963 | 0,634 | -0,296 (-0,645;0,053) |
| TG(51:3) | -0,272 | 0,136 | 0,634 | -0,272 (-0,629;0,085) |
| TG(51:4) | -0,334 | 0,0724 | 0,634 | -0,334 (-0,698;0,03) |
| TG(52:1) | -0,26 | 0,138 | 0,634 | -0,26 (-0,603;0,084) |
| TG(52:3) | -0,25 | 0,188 | 0,634 | -0,25 (-0,623;0,122) |
| TG(52:4) | -0,302 | 0,116 | 0,634 | -0,302 (-0,678;0,074) |
| TG(54:5) | -0,315 | 0,0952 | 0,634 | -0,315 (-0,684;0,055) |
| TG(56:1) | -0,301 | 0,0703 | 0,634 | -0,301 (-0,627;0,025) |
| TG(56:6) | -0,295 | 0,113 | 0,634 | -0,295 (-0,66;0,07) |
| TG(56:7) | -0,282 | 0,132 | 0,634 | -0,282 (-0,65;0,085) |
| TG(57:1) | -0,331 | 0,0763 | 0,634 | -0,331 (-0,698;0,035) |
| TG(58:2) | -0,335 | 0,093 | 0,634 | -0,335 (-0,727;0,056) |
| TG(60:1) | -0,256 | 0,177 | 0,634 | -0,256 (-0,628;0,116) |
| TG(60:3) | -0,278 | 0,139 | 0,634 | -0,278 (-0,647;0,09) |
| Glutathione | -0,184 | 0,195 | 0,64 | -0,184 (-0,463;0,094) |
| L-Methionine | -0,192 | 0,196 | 0,64 | -0,192 (-0,482;0,099) |
| L-2-aminoadipic acid | -0,21 | 0,201 | 0,65 | -0,21 (-0,532;0,112) |
| Phosphatidylethanolamine(38:2) | -0,215 | 0,206 | 0,658 | -0,215 (-0,549;0,119) |
| TG(50:2) | -0,22 | 0,208 | 0,658 | -0,22 (-0,562;0,122) |
| Isobutyrylcarnitine | -0,212 | 0,219 | 0,681 | -0,212 (-0,55;0,126) |
| Sphingomyelin(d18:1/18:0) | 0,212 | 0,219 | 0,681 | 0,212 (-0,126;0,55) |
| Isocitrate | -0,168 | 0,224 | 0,687 | -0,168 (-0,439;0,103) |
| Acetylcarnitine | -0,205 | 0,238 | 0,695 | -0,205 (-0,546;0,135) |
| 8(9)-EpETrE | 0,111 | 0,235 | 0,695 | 0,111 (-0,072;0,294) |
| DG(36:2) | -0,214 | 0,236 | 0,695 | -0,214 (-0,568;0,14) |
| TG(42:0) | -0,193 | 0,237 | 0,695 | -0,193 (-0,512;0,127) |
| TG(52:2) | -0,211 | 0,235 | 0,695 | -0,211 (-0,558;0,137) |
| Linoleoyl ethanolamide | -0,199 | 0,24 | 0,696 | -0,199 (-0,531;0,133) |
| O-Acetyl-L-serine | -0,183 | 0,257 | 0,697 | -0,183 (-0,499;0,133) |
| Palmitoylcarnitine | 0,199 | 0,253 | 0,697 | 0,199 (-0,142;0,539) |
| Lysophosphatidic acid 14(0) | -0,169 | 0,262 | 0,697 | -0,169 (-0,464;0,126) |
| Lysophosphatidylinositol 18(1) | 0,213 | 0,25 | 0,697 | 0,213 (-0,15;0,575) |
| 1-LG/2-LG | -0,192 | 0,264 | 0,697 | -0,192 (-0,528;0,144) |
| 11,12-DiHETrE | -0,195 | 0,249 | 0,697 | -0,195 (-0,526;0,136) |
| 9,10-DiHOME | 0,179 | 0,262 | 0,697 | 0,179 (-0,134;0,493) |
| Phosphatidylethanolamine(38:4) | -0,198 | 0,248 | 0,697 | -0,198 (-0,535;0,138) |
| TG(44:1) | -0,183 | 0,256 | 0,697 | -0,183 (-0,499;0,133) |
| TG(54:2) | -0,202 | 0,253 | 0,697 | -0,202 (-0,547;0,144) |
| TG(55:1) | -0,218 | 0,27 | 0,708 | -0,218 (-0,606;0,17) |
| TG(54:4) | -0,209 | 0,278 | 0,723 | -0,209 (-0,588;0,169) |
| Citrulline | -0,195 | 0,283 | 0,726 | -0,195 (-0,552;0,161) |
| Homocitrulline | -0,193 | 0,284 | 0,726 | -0,193 (-0,546;0,16) |
| Phosphatidylcholine(32:0) | -0,193 | 0,295 | 0,741 | -0,193 (-0,554;0,168) |
| TG(46:1) | -0,177 | 0,293 | 0,741 | -0,177 (-0,507;0,153) |
| Lysophosphatidylcholine(O-18:1) | -0,182 | 0,298 | 0,742 | -0,182 (-0,523;0,16) |
| Sphingomyelin(d18:1/23:1) | -0,184 | 0,305 | 0,745 | -0,184 (-0,535;0,167) |
| TG(42:1) | -0,173 | 0,306 | 0,745 | -0,173 (-0,504;0,158) |
| TG(60:2) | -0,196 | 0,306 | 0,745 | -0,196 (-0,571;0,179) |
| 12,13-DiHOME | 0,179 | 0,313 | 0,749 | 0,179 (-0,168;0,525) |
| Thromboxane-B2 | -0,158 | 0,311 | 0,749 | -0,158 (-0,464;0,148) |
| Lysophosphatidylinositol 20(4) | 0,163 | 0,319 | 0,758 | 0,163 (-0,158;0,485) |
| Decanoylcarnitine | 0,166 | 0,331 | 0,766 | 0,166 (-0,168;0,499) |
| Nonaylcarnitine | -0,152 | 0,343 | 0,766 | -0,152 (-0,467;0,163) |
| Dehydroepiandrosteron | -0,163 | 0,334 | 0,766 | -0,163 (-0,495;0,168) |
| Lysophosphatidylcholine(16:1) | 0,175 | 0,336 | 0,766 | 0,175 (-0,181;0,531) |
| TG(42:2) | -0,17 | 0,337 | 0,766 | -0,17 (-0,516;0,177) |
| TG(44:2) | -0,158 | 0,343 | 0,766 | -0,158 (-0,483;0,168) |
| TG(54:3) | -0,177 | 0,343 | 0,766 | -0,177 (-0,544;0,189) |
| TG(56:5) | -0,185 | 0,328 | 0,766 | -0,185 (-0,556;0,186) |
| Octanoylcarnitine | 0,163 | 0,347 | 0,77 | 0,163 (-0,177;0,504) |
| Lysophosphatidylethanolamine(18:1) | 0,156 | 0,35 | 0,771 | 0,156 (-0,171;0,482) |
| TG(58:9) | -0,162 | 0,353 | 0,771 | -0,162 (-0,503;0,179) |
| 12,13-EpOME | 0,182 | 0,36 | 0,782 | 0,182 (-0,208;0,572) |
| Sphingomyelin(d18:1/21:0) | -0,17 | 0,363 | 0,783 | -0,17 (-0,538;0,197) |
| Hexanoylcarnitine | 0,158 | 0,368 | 0,789 | 0,158 (-0,186;0,503) |
| Taurodeoxycholic acid | -0,146 | 0,375 | 0,797 | -0,146 (-0,47;0,177) |
| DGLEA | -0,156 | 0,381 | 0,805 | -0,156 (-0,506;0,193) |
| Lysophosphatidylcholine(O-16:0) | -0,162 | 0,39 | 0,812 | -0,162 (-0,532;0,207) |
| Phosphatidylcholine(34:3) | -0,154 | 0,39 | 0,812 | -0,154 (-0,504;0,197) |
| S-Methylcysteine | -0,153 | 0,397 | 0,817 | -0,153 (-0,508;0,201) |
| Citric acid | 0,129 | 0,407 | 0,817 | 0,129 (-0,175;0,432) |
| 17,18-DiHETE | 0,14 | 0,4 | 0,817 | 0,14 (-0,187;0,467) |
| Phosphatidylcholine(38:3) | -0,146 | 0,404 | 0,817 | -0,146 (-0,488;0,197) |
| Sphingomyelin(d18:1/25:1) | -0,139 | 0,404 | 0,817 | -0,139 (-0,465;0,187) |
| TG(56:0) | -0,157 | 0,408 | 0,817 | -0,157 (-0,531;0,216) |
| CE(18:2) | -0,15 | 0,411 | 0,817 | -0,15 (-0,507;0,207) |
| L-4-hydroxy-proline | 0,135 | 0,424 | 0,832 | 0,135 (-0,196;0,465) |
| L-Glutamine | 0,142 | 0,423 | 0,832 | 0,142 (-0,205;0,489) |
| TG(44:0) | -0,125 | 0,427 | 0,832 | -0,125 (-0,433;0,183) |
| Cer(d18:0/23:0) | -0,142 | 0,441 | 0,854 | -0,142 (-0,503;0,219) |
| Dodecenoylcarnitine | 0,117 | 0,491 | 0,858 | 0,117 (-0,216;0,45) |
| Hexadecenoylcarnitine | 0,121 | 0,489 | 0,858 | 0,121 (-0,221;0,463) |
| Lauroylcarnitine | 0,125 | 0,469 | 0,858 | 0,125 (-0,214;0,465) |
| Myristoilcarnitine | 0,126 | 0,461 | 0,858 | 0,126 (-0,209;0,461) |
| Trimethylamine N-oxide | 0,125 | 0,486 | 0,858 | 0,125 (-0,226;0,477) |
| 3-Hydroxybutyric acid | -0,133 | 0,462 | 0,858 | -0,133 (-0,488;0,222) |
| Pyroglutamic acid | -0,125 | 0,454 | 0,858 | -0,125 (-0,453;0,203) |
| Lysophosphatidic acid 16(0) | -0,108 | 0,473 | 0,858 | -0,108 (-0,404;0,187) |
| Cortisol | -0,122 | 0,479 | 0,858 | -0,122 (-0,46;0,216) |
| Cer(d18:1/22:0) | -0,133 | 0,448 | 0,858 | -0,133 (-0,476;0,211) |
| DG(36:3) | -0,137 | 0,452 | 0,858 | -0,137 (-0,494;0,22) |
| Lysophosphatidylcholine(18:1) | 0,129 | 0,488 | 0,858 | 0,129 (-0,236;0,494) |
| Lysophosphatidylcholine(O-16:1) | -0,135 | 0,485 | 0,858 | -0,135 (-0,515;0,245) |
| Lysophosphatidylethanolamine(18:0) | -0,127 | 0,479 | 0,858 | -0,127 (-0,477;0,224) |
| Phosphatidylcholine(36:3) | -0,132 | 0,466 | 0,858 | -0,132 (-0,488;0,224) |
| Phosphatidylcholine(36:5) | -0,123 | 0,464 | 0,858 | -0,123 (-0,454;0,207) |
| TG(58:8) | -0,13 | 0,469 | 0,858 | -0,13 (-0,482;0,222) |
| DL-3-aminoisobutyric acid | 0,118 | 0,496 | 0,861 | 0,118 (-0,222;0,458) |
| L-Alanine | -0,111 | 0,535 | 0,869 | -0,111 (-0,463;0,24) |
| L-Arginine | -0,113 | 0,522 | 0,869 | -0,113 (-0,457;0,232) |
| Symmetric dimethylarginine | 0,107 | 0,518 | 0,869 | 0,107 (-0,217;0,431) |
| Lysophosphatidylinositol 18(2) | 0,116 | 0,515 | 0,869 | 0,116 (-0,233;0,464) |
| Sphingosine 1-phosphate 18(0) | -0,047 | 0,54 | 0,869 | -0,047 (-0,196;0,102) |
| Taurolithocholic acid | -0,113 | 0,531 | 0,869 | -0,113 (-0,467;0,241) |
| 5-HETE | 0,103 | 0,535 | 0,869 | 0,103 (-0,223;0,43) |
| CE(22:6) | -0,104 | 0,507 | 0,869 | -0,104 (-0,411;0,203) |
| Phosphatidylcholine(32:1) | 0,107 | 0,539 | 0,869 | 0,107 (-0,234;0,448) |
| Phosphatidylcholine(36:2) | -0,115 | 0,537 | 0,869 | -0,115 (-0,48;0,25) |
| Phosphatidylethanolamine(38:6) | -0,101 | 0,516 | 0,869 | -0,101 (-0,407;0,204) |
| Sphingomyelin(d18:1/25:0) | -0,113 | 0,538 | 0,869 | -0,113 (-0,474;0,248) |
| TG(46:0) | -0,112 | 0,503 | 0,869 | -0,112 (-0,441;0,217) |
| TG(56:3) | -0,117 | 0,53 | 0,869 | -0,117 (-0,481;0,247) |
| Propionylcarnitine | -0,094 | 0,547 | 0,876 | -0,094 (-0,398;0,211) |
| L-Alpha-aminobutyric acid | -0,102 | 0,556 | 0,877 | -0,102 (-0,442;0,238) |
| L-Kynurenine | -0,099 | 0,579 | 0,877 | -0,099 (-0,449;0,251) |
| L-Leucine | -0,085 | 0,585 | 0,877 | -0,085 (-0,388;0,219) |
| L-Serine | -0,089 | 0,582 | 0,877 | -0,089 (-0,407;0,229) |
| Lysophosphatidic acid 18(1) | 0,092 | 0,558 | 0,877 | 0,092 (-0,216;0,4) |
| Taurochenodeoxycholic acid | 0,095 | 0,576 | 0,877 | 0,095 (-0,238;0,428) |
| 8,12-iPF2a IV | -0,082 | 0,568 | 0,877 | -0,082 (-0,364;0,2) |
| 8,9-DiHETrE | -0,092 | 0,564 | 0,877 | -0,092 (-0,403;0,22) |
| 9-HODE | 0,089 | 0,585 | 0,877 | 0,089 (-0,231;0,41) |
| Lysophosphatidylcholine(18:0) | -0,114 | 0,552 | 0,877 | -0,114 (-0,489;0,261) |
| Phosphatidylcholine(32:2) | -0,108 | 0,561 | 0,877 | -0,108 (-0,472;0,256) |
| Phosphatidylcholine(O-44:5) | 0,106 | 0,579 | 0,877 | 0,106 (-0,268;0,48) |
| TG(56:4) | -0,109 | 0,567 | 0,877 | -0,109 (-0,484;0,265) |
| Lysophosphatidylcholine(18:2) | 0,101 | 0,59 | 0,88 | 0,101 (-0,266;0,468) |
| (+/-) 11-HDoHE | -0,056 | 0,594 | 0,881 | -0,056 (-0,261;0,149) |
| 2-Methylbutyroylcarnitine | -0,081 | 0,606 | 0,887 | -0,081 (-0,39;0,228) |
| Butyrylcarnitine | -0,082 | 0,603 | 0,887 | -0,082 (-0,392;0,228) |
| Tetradecenoylcarnitine | 0,089 | 0,611 | 0,887 | 0,089 (-0,254;0,432) |
| Lysophosphatidylethanolamine (20:5) | 0,084 | 0,621 | 0,887 | 0,084 (-0,249;0,417) |
| 12,13-DiHODE | -0,089 | 0,618 | 0,887 | -0,089 (-0,437;0,26) |
| Lysophosphatidylcholine(20:3) | 0,089 | 0,619 | 0,887 | 0,089 (-0,261;0,438) |
| TG(53:1) | -0,089 | 0,614 | 0,887 | -0,089 (-0,434;0,256) |
| TG(56:2) | -0,089 | 0,611 | 0,887 | -0,089 (-0,432;0,254) |
| L-Histidine | -0,086 | 0,624 | 0,887 | -0,086 (-0,43;0,258) |
| Lysophosphatidylethanolamine (16:0) | 0,084 | 0,633 | 0,887 | 0,084 (-0,26;0,428) |
| LPS 18(1) | -0,051 | 0,628 | 0,887 | -0,051 (-0,259;0,156) |
| Cholic acid | -0,084 | 0,636 | 0,887 | -0,084 (-0,431;0,263) |
| Phosphatidylcholine(34:1) | 0,022 | 0,635 | 0,887 | 0,022 (-0,069;0,114) |
| Cysteine | -0,069 | 0,661 | 0,89 | -0,069 (-0,377;0,24) |
| Gamma-L-glutamyl-L-alanine | 0,075 | 0,665 | 0,89 | 0,075 (-0,263;0,412) |
| Taurine | -0,037 | 0,661 | 0,89 | -0,037 (-0,205;0,13) |
| L-Tryptophan | -0,074 | 0,646 | 0,89 | -0,074 (-0,392;0,243) |
| O-Anandamide | -0,081 | 0,659 | 0,89 | -0,081 (-0,443;0,28) |
| Phosphatidylcholine(36:1) | 0,078 | 0,651 | 0,89 | 0,078 (-0,259;0,414) |
| Phosphatidylcholine(40:8) | -0,067 | 0,653 | 0,89 | -0,067 (-0,358;0,224) |
| Phosphatidylethanolamine(36:4) | -0,073 | 0,662 | 0,89 | -0,073 (-0,401;0,255) |
| Sphingomyelin(d18:1/16:0) | -0,088 | 0,661 | 0,89 | -0,088 (-0,479;0,304) |
| FA 18(1) | 0,071 | 0,681 | 0,908 | 0,071 (-0,267;0,409) |
| O-Phosphoethanolamine | -0,027 | 0,691 | 0,91 | -0,027 (-0,161;0,107) |
| Lysophosphatidylcholine(14:0) | -0,075 | 0,691 | 0,91 | -0,075 (-0,445;0,295) |
| Phosphatidylcholine(O-34:2) | 0,071 | 0,691 | 0,91 | 0,071 (-0,28;0,422) |
| (+/-) 14-HDoHE | -0,027 | 0,699 | 0,912 | -0,027 (-0,163;0,109) |
| Lysophosphatidylcholine(20:4) | -0,069 | 0,697 | 0,912 | -0,069 (-0,419;0,281) |
| L-Valine | -0,062 | 0,712 | 0,918 | -0,062 (-0,39;0,267) |
| Carnitine | 0,061 | 0,712 | 0,918 | 0,061 (-0,262;0,384) |
| Sphingomyelin(d18:1/20:0) | 0,07 | 0,71 | 0,918 | 0,07 (-0,297;0,436) |
| L-Asparagine | -0,06 | 0,727 | 0,928 | -0,06 (-0,397;0,277) |
| Pimelylcarnitine | -0,059 | 0,724 | 0,928 | -0,059 (-0,387;0,269) |
| Prostaglandin F2α | 0,049 | 0,734 | 0,934 | 0,049 (-0,232;0,329) |
| Sarcosine | 0,05 | 0,745 | 0,94 | 0,05 (-0,249;0,349) |
| FA 18(2) | -0,052 | 0,742 | 0,94 | -0,052 (-0,365;0,26) |
| Hydroxylysine | 0,051 | 0,78 | 0,941 | 0,051 (-0,308;0,41) |
| Decenoylcarnitine | 0,054 | 0,761 | 0,941 | 0,054 (-0,291;0,399) |
| Stearoylcarnitine | 0,044 | 0,77 | 0,941 | 0,044 (-0,254;0,343) |
| Lysophosphatidic acid 16(1) | 0,042 | 0,775 | 0,941 | 0,042 (-0,246;0,329) |
| (+/-) 10-HDoHE | -0,028 | 0,78 | 0,941 | -0,028 (-0,224;0,168) |
| Anandamide | -0,056 | 0,759 | 0,941 | -0,056 (-0,413;0,301) |
| Prostaglandin E2 | -0,047 | 0,772 | 0,941 | -0,047 (-0,364;0,27) |
| Lysophosphatidylethanolamine(22:6) | -0,054 | 0,754 | 0,941 | -0,054 (-0,392;0,284) |
| Sphingomyelin(d18:1/14:0) | -0,056 | 0,765 | 0,941 | -0,056 (-0,42;0,309) |
| Sphingomyelin(d18:1/24:1) | -0,056 | 0,769 | 0,941 | -0,056 (-0,428;0,316) |
| TG(59:1) | -0,055 | 0,779 | 0,941 | -0,055 (-0,44;0,33) |
| Malic acid | -0,044 | 0,784 | 0,942 | -0,044 (-0,361;0,272) |
| Linoleylcarnitine | -0,039 | 0,797 | 0,947 | -0,039 (-0,337;0,259) |
| Malonylcarnitine | -0,038 | 0,826 | 0,947 | -0,038 (-0,376;0,3) |
| Lysophosphatidic acid 18(2) | 0,037 | 0,817 | 0,947 | 0,037 (-0,281;0,356) |
| 12(S)-HEPE | -0,016 | 0,822 | 0,947 | -0,016 (-0,16;0,127) |
| 12-HETE | 0,017 | 0,822 | 0,947 | 0,017 (-0,133;0,167) |
| 9,10,13-TriHOME | 0,043 | 0,796 | 0,947 | 0,043 (-0,285;0,371) |
| Palmitoyl ethanolamide | 0,041 | 0,824 | 0,947 | 0,041 (-0,318;0,399) |
| CE(18:1) | -0,045 | 0,807 | 0,947 | -0,045 (-0,407;0,317) |
| Lysophosphatidylcholine(16:0) | -0,046 | 0,805 | 0,947 | -0,046 (-0,412;0,32) |
| Phosphatidylethanolamine(34:2) | -0,04 | 0,819 | 0,947 | -0,04 (-0,386;0,306) |
| Sphingomyelin(d18:1/24:2) | 0,044 | 0,814 | 0,947 | 0,044 (-0,327;0,416) |
| TG(45:0) | -0,046 | 0,801 | 0,947 | -0,046 (-0,404;0,312) |
| L-Proline | 0,035 | 0,837 | 0,949 | 0,035 (-0,3;0,371) |
| Oleoylcarnitine | 0,036 | 0,836 | 0,949 | 0,036 (-0,308;0,38) |
| Valerylcarnitine | -0,034 | 0,838 | 0,949 | -0,034 (-0,357;0,289) |
| 9,12,13-TriHOME | 0,034 | 0,842 | 0,949 | 0,034 (-0,301;0,369) |
| Phosphatidylcholine(O-36:3) | -0,033 | 0,846 | 0,949 | -0,033 (-0,368;0,302) |
| TG(54:0) | -0,037 | 0,846 | 0,949 | -0,037 (-0,414;0,339) |
| Lactic acid | -0,033 | 0,851 | 0,951 | -0,033 (-0,375;0,31) |
| 8-HETE | -0,022 | 0,858 | 0,953 | -0,022 (-0,267;0,223) |
| Phosphatidylcholine(34:2) | -0,033 | 0,859 | 0,953 | -0,033 (-0,397;0,331) |
| Isovalerylcarnitine | -0,026 | 0,868 | 0,958 | -0,026 (-0,331;0,279) |
| 5,6-DiHETrE | -0,028 | 0,87 | 0,958 | -0,028 (-0,361;0,306) |
| L-Isoleucine | 0,023 | 0,886 | 0,962 | 0,023 (-0,295;0,341) |
| Lysophosphatidylinositol 18(0) | -0,023 | 0,885 | 0,962 | -0,023 (-0,338;0,291) |
| Sphingosine 1-phosphate 18(1) | 0,012 | 0,88 | 0,962 | 0,012 (-0,139;0,162) |
| Phosphatidylcholine(O-34:3) | -0,027 | 0,884 | 0,962 | -0,027 (-0,39;0,336) |
| (+/-) 8-HDoHE | 0,026 | 0,891 | 0,964 | 0,026 (-0,341;0,392) |
| Lysophosphatidylcholine(20:5) | 0,019 | 0,896 | 0,966 | 0,019 (-0,273;0,312) |
| Glycodeoxycholic acid | 0,02 | 0,904 | 0,972 | 0,02 (-0,311;0,352) |
| Lysophosphatidylcholine(22:6) | -0,018 | 0,914 | 0,974 | -0,018 (-0,348;0,312) |
| Lysophosphatidylethanolamine(20:4) | 0,019 | 0,92 | 0,974 | 0,019 (-0,344;0,381) |
| Sphingomyelin(d18:1/22:0) | -0,02 | 0,913 | 0,974 | -0,02 (-0,385;0,344) |
| Sphingomyelin(d18:1/22:1) | 0,019 | 0,918 | 0,974 | 0,019 (-0,352;0,39) |
| Sphingomyelin(d18:1/16:1) | -0,018 | 0,924 | 0,975 | -0,018 (-0,395;0,358) |
| L-Glutamic acid | -0,013 | 0,934 | 0,977 | -0,013 (-0,325;0,298) |
| 14,15-DiHETrE | -0,014 | 0,935 | 0,977 | -0,014 (-0,351;0,323) |
| Phosphatidylcholine(O-34:1) | 0,015 | 0,934 | 0,977 | 0,015 (-0,345;0,375) |
| L-Lysine | -0,005 | 0,979 | 0,985 | -0,005 (-0,346;0,337) |
| L-Phenylalanine | 0,01 | 0,954 | 0,985 | 0,01 (-0,313;0,332) |
| Octenoylcarnitine | 0,004 | 0,983 | 0,985 | 0,004 (-0,342;0,35) |
| Tetradecadienylcarnitine | 0,009 | 0,957 | 0,985 | 0,009 (-0,335;0,354) |
| Lysophosphatidylethanolamine (20:4) | -0,007 | 0,964 | 0,985 | -0,007 (-0,315;0,301) |
| 13-HODE | -0,005 | 0,974 | 0,985 | -0,005 (-0,289;0,28) |
| 14,15-DiHETE | 0,005 | 0,978 | 0,985 | 0,005 (-0,323;0,332) |
| 19,20-DiHDPA | -0,004 | 0,98 | 0,985 | -0,004 (-0,321;0,313) |
| Glycolithocholic acid | -0,003 | 0,985 | 0,985 | -0,003 (-0,344;0,337) |
| Cer(d18:1/24:1) | 0,003 | 0,985 | 0,985 | 0,003 (-0,34;0,347) |
| Lysophosphatidylcholine(18:3) | -0,007 | 0,972 | 0,985 | -0,007 (-0,388;0,374) |
| Phosphatidylcholine(O-38:4) | -0,007 | 0,968 | 0,985 | -0,007 (-0,366;0,351) |
| Sphingomyelin(d18:1/24:0) | 0,007 | 0,971 | 0,985 | 0,007 (-0,358;0,372) |

**Supplementary Table 6. Metabolites differently expressed in IPD patients without L-Dopa treatment (Dopa^negative^, n = 30) compared to controls (n = 64).** Shown are the results of the fixed effect meta-analysis: regression coefficient (CI: 95% confidence interval), nominal and adjusted p-values.

| **Name** | **z-value** | **Nominal *p*** | **Adjusted *p*** | **Beta (CI)** |
| --- | --- | --- | --- | --- |
| Lysophosphatidic acid 16(0) | -3,418 | 0,000631 | 0,0959 | -0,685 (-1,078;-0,292) |
| 1-AG/2-AG | -3,589 | 0,000332 | 0,0959 | -0,762 (-1,178;-0,346) |
| Glutathione | 2,751 | 0,00594 | 0,301 | 0,432 (0,124;0,74) |
| 1-LG/2-LG | -2,766 | 0,00567 | 0,301 | -0,573 (-0,978;-0,167) |
| Phosphatidylcholine(36:4) | -2,858 | 0,00426 | 0,301 | -0,634 (-1,069;-0,199) |
| Phosphatidylcholine(38:7) | -2,764 | 0,0057 | 0,301 | -0,571 (-0,975;-0,166) |
| Citric acid | 2,618 | 0,00885 | 0,384 | 0,57 (0,143;0,997) |
| Hexadecenoylcarnitine | 2,532 | 0,0114 | 0,432 | 0,533 (0,12;0,946) |
| Phosphatidylethanolamine(34:2) | -2,458 | 0,014 | 0,449 | -0,606 (-1,089;-0,123) |
| Phosphatidylethanolamine(36:4) | -2,438 | 0,0148 | 0,449 | -0,58 (-1,046;-0,114) |
| Oleoylcarnitine | 2,383 | 0,0172 | 0,474 | 0,505 (0,09;0,92) |
| Tiglylcarnitine | -2,321 | 0,0203 | 0,514 | -0,61 (-1,126;-0,095) |
| Putrescine | 2,227 | 0,026 | 0,56 | 0,386 (0,046;0,725) |
| Lysophosphatidic acid 14(0) | -2,107 | 0,0351 | 0,56 | -0,398 (-0,768;-0,028) |
| Glycoursodeoxycholic acid | -2,117 | 0,0342 | 0,56 | -0,449 (-0,865;-0,033) |
| Thromboxane-B2 | -2,132 | 0,033 | 0,56 | -0,374 (-0,718;-0,03) |
| Phosphatidylcholine(O-36:5) | -2,088 | 0,0368 | 0,56 | -0,505 (-0,978;-0,031) |
| TG(50:1) | -2,195 | 0,0282 | 0,56 | -0,49 (-0,928;-0,052) |
| TG(52:1) | -2,176 | 0,0296 | 0,56 | -0,49 (-0,932;-0,049) |
| TG(54:7) | -2,154 | 0,0313 | 0,56 | -0,515 (-0,984;-0,046) |
| FA 18(1) | 2,024 | 0,043 | 0,623 | 0,486 (0,015;0,956) |
| L-2-aminoadipic acid | -1,745 | 0,0811 | 0,653 | -0,315 (-0,669;0,039) |
| L-Arginine | 1,799 | 0,072 | 0,653 | 0,407 (-0,036;0,851) |
| L-Serine | 1,8 | 0,0719 | 0,653 | 0,47 (-0,042;0,982) |
| Tetradecenoylcarnitine | 1,963 | 0,0497 | 0,653 | 0,445 (0,001;0,89) |
| Lysophosphatidic acid 18(2) | -1,882 | 0,0598 | 0,653 | -0,411 (-0,84;0,017) |
| LPS 18(1) | -1,81 | 0,0704 | 0,653 | -0,257 (-0,536;0,021) |
| (+/-) 11-HDoHE | 1,717 | 0,086 | 0,653 | 0,234 (-0,033;0,501) |
| 19,20-DiHDPA | 1,721 | 0,0853 | 0,653 | 0,458 (-0,064;0,979) |
| (+/-) 8-HDoHE | 1,72 | 0,0855 | 0,653 | 0,443 (-0,062;0,949) |
| Prostaglandin E2 | -1,744 | 0,0812 | 0,653 | -0,305 (-0,648;0,038) |
| Phosphatidylcholine(38:4) | -1,723 | 0,0848 | 0,653 | -0,402 (-0,86;0,055) |
| Phosphatidylethanolamine(O-36:5) | -1,855 | 0,0636 | 0,653 | -0,543 (-1,117;0,031) |
| TG(50:2) | -1,805 | 0,0711 | 0,653 | -0,405 (-0,844;0,035) |
| TG(50:3) | -1,773 | 0,0762 | 0,653 | -0,42 (-0,885;0,044) |
| TG(50:4) | -1,797 | 0,0724 | 0,653 | -0,449 (-0,938;0,041) |
| TG(52:4) | -1,726 | 0,0844 | 0,653 | -0,414 (-0,885;0,056) |
| TG(54:1) | -1,928 | 0,0538 | 0,653 | -0,522 (-1,053;0,009) |
| TG(54:5) | -1,855 | 0,0635 | 0,653 | -0,46 (-0,946;0,026) |
| TG(56:6) | -1,968 | 0,049 | 0,653 | -0,478 (-0,955;-0,002) |
| 13-HODE | 1,693 | 0,0904 | 0,67 | 0,34 (-0,054;0,734) |
| Cer(d18:1/22:0) | -1,657 | 0,0975 | 0,674 | -0,334 (-0,73;0,061) |
| DG(36:2) | -1,659 | 0,097 | 0,674 | -0,14 (-0,305;0,025) |
| TG(48:1) | -1,67 | 0,0949 | 0,674 | -0,391 (-0,849;0,068) |
| Citrulline | 1,345 | 0,178 | 0,711 | 0,243 (-0,111;0,596) |
| Gamma-Glutamylglutamine | 1,221 | 0,222 | 0,711 | 0,287 (-0,174;0,748) |
| L-Glutamic acid | -1,449 | 0,147 | 0,711 | -0,322 (-0,757;0,114) |
| L-Kynurenine | -1,269 | 0,204 | 0,711 | -0,313 (-0,796;0,17) |
| Saccharopine | 1,342 | 0,18 | 0,711 | 0,335 (-0,154;0,823) |
| Sarcosine | 1,241 | 0,215 | 0,711 | 0,27 (-0,156;0,697) |
| Serotonine | 1,145 | 0,252 | 0,711 | 0,14 (-0,099;0,378) |
| L-Threonine | 1,394 | 0,163 | 0,711 | 0,363 (-0,147;0,873) |
| 2-Methylbutyroylcarnitine | -1,371 | 0,17 | 0,711 | -0,323 (-0,784;0,139) |
| Acetylcarnitine | 1,171 | 0,241 | 0,711 | 0,277 (-0,186;0,74) |
| Betaine | 1,196 | 0,232 | 0,711 | 0,311 (-0,199;0,821) |
| Dodecenoylcarnitine | 1,595 | 0,111 | 0,711 | 0,365 (-0,084;0,814) |
| Isobutyrylcarnitine | -1,275 | 0,202 | 0,711 | -0,302 (-0,766;0,162) |
| Myristoilcarnitine | 1,278 | 0,201 | 0,711 | 0,269 (-0,144;0,682) |
| Octanoylcarnitine | 1,133 | 0,257 | 0,711 | 0,279 (-0,204;0,763) |
| Palmitoylcarnitine | 1,33 | 0,183 | 0,711 | 0,269 (-0,127;0,665) |
| Propionylcarnitine | -1,37 | 0,171 | 0,711 | -0,321 (-0,78;0,138) |
| Tetradecadienylcarnitine | 1,17 | 0,242 | 0,711 | 0,269 (-0,182;0,719) |
| 2-ketoglutaric acid | -1,198 | 0,231 | 0,711 | -0,267 (-0,705;0,17) |
| FA 22(6) | 1,155 | 0,248 | 0,711 | 0,282 (-0,197;0,761) |
| Lysophosphatidic acid 16(1) | -1,544 | 0,123 | 0,711 | -0,302 (-0,686;0,081) |
| Lysophosphatidic acid 18(1) | -1,296 | 0,195 | 0,711 | -0,283 (-0,711;0,145) |
| Lysophosphatidylethanolamine (20:4) | -1,184 | 0,237 | 0,711 | -0,311 (-0,825;0,204) |
| (+/-) 10-HDoHE | 1,331 | 0,183 | 0,711 | 0,183 (-0,086;0,452) |
| 12(S)-HEPE | 1,161 | 0,246 | 0,711 | 0,117 (-0,081;0,315) |
| 14,15-DiHETE | 1,198 | 0,231 | 0,711 | 0,308 (-0,196;0,813) |
| (+/-) 14-HDoHE | 1,433 | 0,152 | 0,711 | 0,133 (-0,049;0,316) |
| 17,18-DiHETE | 1,367 | 0,172 | 0,711 | 0,35 (-0,152;0,852) |
| 5,6-DiHETrE | -1,174 | 0,24 | 0,711 | -0,292 (-0,78;0,196) |
| 9,10,13-TriHOME | 1,218 | 0,223 | 0,711 | 0,228 (-0,139;0,594) |
| 9,12,13-TriHOME | 1,257 | 0,209 | 0,711 | 0,295 (-0,165;0,756) |
| Cortisol | 1,232 | 0,218 | 0,711 | 0,306 (-0,181;0,792) |
| Dehydroepiandrosteron | 1,533 | 0,125 | 0,711 | 0,394 (-0,11;0,897) |
| Glycochenodeoxycholic acid | -1,593 | 0,111 | 0,711 | -0,329 (-0,733;0,076) |
| Linoleoyl ethanolamide | 1,143 | 0,253 | 0,711 | 0,271 (-0,194;0,736) |
| Palmitoyl ethanolamide | 1,23 | 0,219 | 0,711 | 0,309 (-0,183;0,801) |
| CE(20:4) | -1,564 | 0,118 | 0,711 | -0,384 (-0,866;0,097) |
| CE(22:6) | 1,231 | 0,218 | 0,711 | 0,322 (-0,19;0,834) |
| Cer(d18:0/22:0) | -1,207 | 0,228 | 0,711 | -0,319 (-0,837;0,199) |
| Lysophosphatidylcholine(16:0) | -1,613 | 0,107 | 0,711 | -0,393 (-0,87;0,084) |
| Lysophosphatidylcholine(22:6) | 1,469 | 0,142 | 0,711 | 0,42 (-0,14;0,981) |
| Lysophosphatidylethanolamine(18:1) | -1,146 | 0,252 | 0,711 | -0,082 (-0,222;0,058) |
| Phosphatidylcholine(36:3) | -1,318 | 0,188 | 0,711 | -0,286 (-0,712;0,139) |
| Phosphatidylcholine(38:3) | -1,267 | 0,205 | 0,711 | -0,286 (-0,729;0,157) |
| Phosphatidylcholine(40:5) | -1,26 | 0,208 | 0,711 | -0,315 (-0,805;0,175) |
| Phosphatidylcholine(O-36:3) | 1,217 | 0,224 | 0,711 | 0,208 (-0,127;0,543) |
| Phosphatidylethanolamine(O-38:5) | -1,466 | 0,143 | 0,711 | -0,452 (-1,055;0,152) |
| Sphingomyelin(d18:1/16:0) | -1,446 | 0,148 | 0,711 | -0,333 (-0,785;0,119) |
| Sphingomyelin(d18:1/16:1) | -1,231 | 0,218 | 0,711 | -0,273 (-0,708;0,162) |
| Sphingomyelin(d18:1/25:1) | 1,4 | 0,162 | 0,711 | 0,362 (-0,145;0,87) |
| TG(45:0) | 1,509 | 0,131 | 0,711 | 0,183 (-0,055;0,421) |
| TG(46:1) | -1,138 | 0,255 | 0,711 | -0,238 (-0,648;0,172) |
| TG(48:2) | -1,573 | 0,116 | 0,711 | -0,37 (-0,831;0,091) |
| TG(48:3) | -1,211 | 0,226 | 0,711 | -0,285 (-0,745;0,176) |
| TG(51:1) | -1,188 | 0,235 | 0,711 | -0,26 (-0,688;0,169) |
| TG(51:2) | -1,135 | 0,256 | 0,711 | -0,26 (-0,71;0,189) |
| TG(51:3) | -1,337 | 0,181 | 0,711 | -0,315 (-0,776;0,147) |
| TG(52:2) | -1,337 | 0,181 | 0,711 | -0,299 (-0,737;0,139) |
| TG(52:3) | -1,576 | 0,115 | 0,711 | -0,371 (-0,832;0,09) |
| TG(54:2) | -1,175 | 0,24 | 0,711 | -0,253 (-0,675;0,169) |
| TG(54:4) | -1,268 | 0,205 | 0,711 | -0,32 (-0,815;0,175) |
| TG(56:0) | 1,161 | 0,246 | 0,711 | 0,177 (-0,122;0,476) |
| TG(56:5) | -1,466 | 0,143 | 0,711 | -0,349 (-0,817;0,118) |
| TG(56:7) | -1,16 | 0,246 | 0,711 | -0,291 (-0,783;0,201) |
| TG(57:1) | 1,558 | 0,119 | 0,711 | 0,278 (-0,072;0,627) |
| TG(59:1) | 1,501 | 0,133 | 0,711 | 0,29 (-0,089;0,669) |
| Phosphatidylcholine(34:1) | -1,104 | 0,27 | 0,738 | -0,06 (-0,167;0,047) |
| DL-3-aminoisobutyric acid | -1,073 | 0,283 | 0,769 | -0,263 (-0,743;0,217) |
| Decanoylcarnitine | 1,053 | 0,292 | 0,78 | 0,251 (-0,217;0,719) |
| Methylmalonylcarnitine | -1,057 | 0,291 | 0,78 | -0,275 (-0,784;0,235) |
| Pimelylcarnitine | 1,044 | 0,297 | 0,784 | 0,229 (-0,201;0,659) |
| Phosphatidylcholine(34:4) | -1,026 | 0,305 | 0,792 | -0,246 (-0,717;0,224) |
| TG(46:0) | -1,031 | 0,302 | 0,792 | -0,182 (-0,528;0,164) |
| Ethanolamine | 1,011 | 0,312 | 0,792 | 0,229 (-0,215;0,674) |
| Pyroglutamic acid | -1,009 | 0,313 | 0,792 | -0,269 (-0,792;0,253) |
| Lysophosphatidylcholine(20:5) | 1,018 | 0,309 | 0,792 | 0,227 (-0,21;0,664) |
| 9-HOTrE | 0,995 | 0,32 | 0,797 | 0,224 (-0,217;0,664) |
| Phosphatidylcholine(O-44:5) | 0,999 | 0,318 | 0,797 | 0,263 (-0,253;0,78) |
| Lauroylcarnitine | 0,988 | 0,323 | 0,798 | 0,229 (-0,225;0,682) |
| 3-Hydroxybutyric acid | 0,953 | 0,34 | 0,803 | 0,249 (-0,263;0,762) |
| Lysophosphatidylinositol 16(1) | -0,96 | 0,337 | 0,803 | -0,221 (-0,671;0,23) |
| Prostaglandin F2α | 0,965 | 0,335 | 0,803 | 0,173 (-0,179;0,525) |
| Phosphatidylethanolamine(38:4) | -0,977 | 0,329 | 0,803 | -0,23 (-0,692;0,231) |
| TG(44:1) | -0,969 | 0,333 | 0,803 | -0,167 (-0,504;0,171) |
| TG(51:4) | -0,953 | 0,341 | 0,803 | -0,219 (-0,67;0,232) |
| L-Homoserine | 0,907 | 0,364 | 0,822 | 0,173 (-0,201;0,547) |
| Sphingosine 1-phosphate 18(0) | -0,895 | 0,371 | 0,822 | -0,093 (-0,298;0,111) |
| 12,13-DiHOME | 0,899 | 0,369 | 0,822 | 0,228 (-0,269;0,724) |
| 8(9)-EpETrE | 0,891 | 0,373 | 0,822 | 0,122 (-0,146;0,39) |
| Lysophosphatidylcholine(20:3) | -0,911 | 0,362 | 0,822 | -0,217 (-0,685;0,25) |
| Sphingomyelin(d18:1/18:2) | -0,895 | 0,371 | 0,822 | -0,288 (-0,918;0,342) |
| TG(42:0) | -0,892 | 0,372 | 0,822 | -0,084 (-0,267;0,1) |
| TG(42:1) | -0,893 | 0,372 | 0,822 | -0,162 (-0,517;0,193) |
| TG(46:2) | -0,915 | 0,36 | 0,822 | -0,171 (-0,536;0,195) |
| Malonylcarnitine | 0,88 | 0,379 | 0,829 | 0,199 (-0,244;0,642) |
| TG(56:1) | 0,869 | 0,385 | 0,835 | 0,116 (-0,146;0,378) |
| Nonaylcarnitine | -0,839 | 0,401 | 0,859 | -0,217 (-0,723;0,29) |
| Sphingomyelin(d18:1/20:1) | 0,843 | 0,399 | 0,859 | 0,191 (-0,253;0,635) |
| Lysophosphatidylcholine(20:4) | -0,832 | 0,405 | 0,862 | -0,212 (-0,712;0,288) |
| Glycine | 0,755 | 0,45 | 0,864 | 0,166 (-0,265;0,597) |
| Hydroxylysine | -0,691 | 0,49 | 0,864 | -0,19 (-0,729;0,349) |
| L-Lysine | 0,739 | 0,46 | 0,864 | 0,2 (-0,33;0,729) |
| O-Acetyl-L-serine | -0,746 | 0,456 | 0,864 | -0,174 (-0,63;0,283) |
| L-Proline | -0,761 | 0,447 | 0,864 | -0,187 (-0,667;0,294) |
| L-Tyrosine | -0,692 | 0,489 | 0,864 | -0,147 (-0,564;0,27) |
| Stearoylcarnitine | 0,795 | 0,426 | 0,864 | 0,18 (-0,264;0,624) |
| Trimethylamine N-oxide | 0,767 | 0,443 | 0,864 | 0,147 (-0,228;0,522) |
| Lactic acid | -0,688 | 0,491 | 0,864 | -0,145 (-0,558;0,268) |
| Lysophosphatidylethanolamine (16:0) | -0,683 | 0,494 | 0,864 | -0,184 (-0,714;0,345) |
| Lysophosphatidylethanolamine (16:1) | -0,693 | 0,488 | 0,864 | -0,161 (-0,615;0,294) |
| Lysophosphatidylinositol 18(1) | -0,739 | 0,46 | 0,864 | -0,171 (-0,626;0,283) |
| 12,13-DiHODE | 0,823 | 0,41 | 0,864 | 0,161 (-0,222;0,544) |
| 8,12-iPF2a IV | -0,685 | 0,493 | 0,864 | -0,148 (-0,571;0,275) |
| Anandamide | 0,728 | 0,466 | 0,864 | 0,184 (-0,312;0,681) |
| Cholic acid | -0,719 | 0,472 | 0,864 | -0,142 (-0,53;0,245) |
| O-Anandamide | 0,805 | 0,421 | 0,864 | 0,198 (-0,285;0,681) |
| CE(18:2) | 0,779 | 0,436 | 0,864 | 0,196 (-0,296;0,688) |
| Lysophosphatidylcholine(O-16:1) | -0,688 | 0,492 | 0,864 | -0,165 (-0,633;0,304) |
| Phosphatidylcholine(34:2) | -0,732 | 0,464 | 0,864 | -0,176 (-0,647;0,295) |
| Phosphatidylcholine(40:6) | 0,753 | 0,452 | 0,864 | 0,183 (-0,293;0,658) |
| Phosphatidylcholine(40:7) | 0,706 | 0,48 | 0,864 | 0,165 (-0,293;0,624) |
| Phosphatidylcholine(O-36:4) | -0,697 | 0,486 | 0,864 | -0,188 (-0,718;0,341) |
| Phosphatidylethanolamine(38:6) | -0,732 | 0,464 | 0,864 | -0,191 (-0,703;0,321) |
| Sphingomyelin(d18:1/23:0) | -0,683 | 0,495 | 0,864 | -0,175 (-0,678;0,328) |
| TG(42:2) | -0,794 | 0,427 | 0,864 | -0,166 (-0,575;0,244) |
| TG(44:0) | -0,812 | 0,417 | 0,864 | -0,1 (-0,341;0,141) |
| TG(54:3) | -0,792 | 0,429 | 0,864 | -0,183 (-0,635;0,269) |
| TG(56:3) | -0,797 | 0,425 | 0,864 | -0,169 (-0,586;0,247) |
| TG(56:4) | -0,692 | 0,489 | 0,864 | -0,166 (-0,634;0,303) |
| TG(60:2) | 0,753 | 0,451 | 0,864 | 0,162 (-0,259;0,582) |
| Linoleylcarnitine | 0,643 | 0,52 | 0,888 | 0,127 (-0,26;0,514) |
| Malic acid | 0,656 | 0,512 | 0,888 | 0,128 (-0,255;0,511) |
| Taurodeoxycholic acid | -0,647 | 0,518 | 0,888 | -0,166 (-0,669;0,337) |
| TG(58:9) | -0,65 | 0,516 | 0,888 | -0,152 (-0,61;0,306) |
| Gamma-aminobutyric acid | -0,633 | 0,526 | 0,889 | -0,125 (-0,511;0,261) |
| Hexanoylcarnitine | 0,635 | 0,525 | 0,889 | 0,155 (-0,323;0,634) |
| Phosphatidylcholine(O-34:1) | 0,626 | 0,531 | 0,889 | 0,131 (-0,279;0,54) |
| Phosphatidylcholine(O-38:6) | 0,624 | 0,532 | 0,889 | 0,177 (-0,38;0,735) |
| Cysteine | 0,523 | 0,601 | 0,894 | 0,115 (-0,315;0,545) |
| O-Phosphoethanolamine | 0,524 | 0,6 | 0,894 | 0,048 (-0,132;0,228) |
| Ornithine | 0,6 | 0,549 | 0,894 | 0,139 (-0,316;0,595) |
| L-Phenylalanine | -0,581 | 0,561 | 0,894 | -0,151 (-0,661;0,358) |
| S-Methylcysteine | -0,52 | 0,603 | 0,894 | -0,12 (-0,57;0,331) |
| Taurine | 0,549 | 0,583 | 0,894 | 0,069 (-0,178;0,316) |
| Carnitine | -0,572 | 0,567 | 0,894 | -0,149 (-0,658;0,361) |
| Isocitrate | -0,576 | 0,565 | 0,894 | -0,107 (-0,47;0,256) |
| 11,12-DiHETrE | 0,531 | 0,596 | 0,894 | 0,132 (-0,356;0,621) |
| 12-HETE | 0,53 | 0,596 | 0,894 | 0,053 (-0,144;0,25) |
| 15(S)-HETrE | 0,578 | 0,563 | 0,894 | 0,114 (-0,273;0,501) |
| 8-HETE | 0,574 | 0,566 | 0,894 | 0,106 (-0,255;0,467) |
| 9,10-DiHOME | 0,528 | 0,598 | 0,894 | 0,124 (-0,338;0,587) |
| 9-HODE | 0,524 | 0,6 | 0,894 | 0,107 (-0,292;0,506) |
| Glycodeoxycholic acid | -0,535 | 0,593 | 0,894 | -0,136 (-0,634;0,362) |
| DG(36:3) | -0,543 | 0,587 | 0,894 | -0,059 (-0,271;0,154) |
| Lysophosphatidylcholine(O-18:1) | -0,547 | 0,584 | 0,894 | -0,128 (-0,588;0,331) |
| Lysophosphatidylethanolamine(20:4) | -0,576 | 0,564 | 0,894 | -0,131 (-0,575;0,314) |
| Phosphatidylcholine(38:6) | 0,552 | 0,581 | 0,894 | 0,141 (-0,36;0,642) |
| Sphingomyelin(d18:1/15:0) | -0,603 | 0,546 | 0,894 | -0,144 (-0,611;0,323) |
| TG(44:2) | -0,591 | 0,555 | 0,894 | -0,104 (-0,451;0,242) |
| TG(53:1) | -0,571 | 0,568 | 0,894 | -0,137 (-0,609;0,334) |
| TG(60:1) | 0,56 | 0,576 | 0,894 | 0,12 (-0,3;0,539) |
| Lysophosphatidylethanolamine(22:6) | 0,511 | 0,609 | 0,899 | 0,123 (-0,349;0,595) |
| L-Methionine | 0,506 | 0,613 | 0,9 | 0,117 (-0,335;0,568) |
| 14,15-DiHETrE | 0,492 | 0,622 | 0,91 | 0,119 (-0,356;0,594) |
| FA 22(5) w6 | 0,486 | 0,627 | 0,911 | 0,109 (-0,331;0,549) |
| Lysophosphatidylinositol 18(0) | -0,483 | 0,629 | 0,911 | -0,113 (-0,574;0,347) |
| 12,13-EpOME | -0,474 | 0,635 | 0,911 | -0,129 (-0,664;0,405) |
| Lysophosphatidylcholine(14:0) | -0,477 | 0,633 | 0,911 | -0,124 (-0,635;0,386) |
| Hydroxybutyric acid | -0,422 | 0,673 | 0,935 | -0,088 (-0,499;0,322) |
| Taurocholic acid | -0,441 | 0,659 | 0,935 | -0,115 (-0,627;0,396) |
| 11-HETE | -0,417 | 0,676 | 0,935 | -0,09 (-0,514;0,334) |
| 8,9-DiHETrE | -0,428 | 0,669 | 0,935 | -0,098 (-0,548;0,351) |
| Glycocholic acid | -0,432 | 0,666 | 0,935 | -0,11 (-0,607;0,388) |
| Glycolithocholic acid | -0,426 | 0,67 | 0,935 | -0,103 (-0,579;0,372) |
| Sphingomyelin(d18:1/14:0) | -0,436 | 0,663 | 0,935 | -0,106 (-0,584;0,371) |
| TG(58:8) | 0,42 | 0,674 | 0,935 | 0,102 (-0,375;0,58) |
| Lysophosphatidylethanolamine(18:0) | -0,409 | 0,682 | 0,938 | -0,102 (-0,591;0,387) |
| Sphingomyelin(d18:1/24:2) | 0,403 | 0,687 | 0,94 | 0,091 (-0,352;0,534) |
| L-Isoleucine | -0,393 | 0,695 | 0,947 | -0,086 (-0,517;0,344) |
| Cystathionine | 0,336 | 0,737 | 0,96 | 0,063 (-0,307;0,434) |
| Decenoylcarnitine | 0,342 | 0,732 | 0,96 | 0,084 (-0,399;0,568) |
| Lysophosphatidylethanolamine (20:5) | 0,334 | 0,739 | 0,96 | 0,083 (-0,402;0,567) |
| Lysophosphatidylcholine(18:0) | -0,339 | 0,735 | 0,96 | -0,082 (-0,554;0,391) |
| Lysophosphatidylcholine(18:1) | 0,347 | 0,728 | 0,96 | 0,086 (-0,399;0,571) |
| Lysophosphatidylcholine(O-16:0) | -0,37 | 0,711 | 0,96 | -0,101 (-0,634;0,433) |
| Phosphatidylcholine(36:1) | -0,361 | 0,718 | 0,96 | -0,075 (-0,484;0,334) |
| Phosphatidylcholine(36:2) | -0,336 | 0,737 | 0,96 | -0,069 (-0,468;0,331) |
| Phosphatidylcholine(O-38:5) | -0,357 | 0,721 | 0,96 | -0,096 (-0,622;0,43) |
| TG(58:2) | 0,354 | 0,723 | 0,96 | 0,06 (-0,272;0,392) |
| TG(60:3) | 0,333 | 0,739 | 0,96 | 0,088 (-0,431;0,608) |
| Homocysteine | 0,294 | 0,769 | 0,973 | 0,066 (-0,375;0,508) |
| L-Valine | 0,291 | 0,771 | 0,973 | 0,065 (-0,374;0,504) |
| Valerylcarnitine | -0,295 | 0,768 | 0,973 | -0,072 (-0,551;0,407) |
| 3-Hydroxypropionic Acid | -0,281 | 0,779 | 0,973 | -0,062 (-0,493;0,369) |
| Lysophosphatidylinositol 18(2) | -0,287 | 0,774 | 0,973 | -0,068 (-0,536;0,399) |
| 5-HETE | -0,273 | 0,785 | 0,973 | -0,063 (-0,514;0,389) |
| Cer(d18:0/23:0) | -0,274 | 0,784 | 0,973 | -0,068 (-0,556;0,42) |
| Phosphatidylcholine(34:3) | -0,27 | 0,787 | 0,973 | -0,072 (-0,595;0,451) |
| Phosphatidylcholine(40:8) | -0,285 | 0,775 | 0,973 | -0,062 (-0,49;0,365) |
| Sphingomyelin(d18:1/20:0) | 0,294 | 0,769 | 0,973 | 0,071 (-0,4;0,542) |
| Sphingomyelin(d18:1/21:0) | -0,29 | 0,772 | 0,973 | -0,074 (-0,574;0,426) |
| Sphingomyelin(d18:1/22:0) | -0,274 | 0,784 | 0,973 | -0,068 (-0,552;0,416) |
| Phosphatidylcholine(O-38:4) | -0,264 | 0,792 | 0,975 | -0,062 (-0,525;0,401) |
| Symmetric dimethylarginine | -0,251 | 0,802 | 0,975 | -0,064 (-0,566;0,438) |
| L-Tryptophan | -0,254 | 0,8 | 0,975 | -0,057 (-0,495;0,381) |
| TG(54:0) | 0,252 | 0,801 | 0,975 | 0,069 (-0,465;0,602) |
| Sphingomyelin(d18:1/24:0) | -0,245 | 0,807 | 0,977 | -0,061 (-0,548;0,426) |
| Sphingomyelin(d18:1/23:1) | -0,24 | 0,811 | 0,978 | -0,056 (-0,518;0,405) |
| L-Alpha-aminobutyric acid | -0,235 | 0,814 | 0,978 | -0,059 (-0,551;0,433) |
| L-4-hydroxy-proline | -0,228 | 0,819 | 0,981 | -0,055 (-0,526;0,416) |
| Methyldopa | -0,212 | 0,832 | 0,992 | -0,018 (-0,187;0,15) |
| 1-Methylhistidine | 0,027 | 0,978 | 0,992 | 0,006 (-0,457;0,469) |
| 3-Methoxytyrosine | 0,113 | 0,91 | 0,992 | 0,004 (-0,071;0,079) |
| L-Alanine | -0,178 | 0,859 | 0,992 | -0,043 (-0,514;0,428) |
| L-Asparagine | 0,068 | 0,946 | 0,992 | 0,015 (-0,422;0,452) |
| Gamma-L-glutamyl-L-alanine | -0,023 | 0,982 | 0,992 | -0,005 (-0,419;0,409) |
| L-Glutamine | -0,022 | 0,982 | 0,992 | -0,005 (-0,454;0,444) |
| Glycylglycine | -0,099 | 0,921 | 0,992 | -0,024 (-0,491;0,444) |
| L-Histidine | -0,032 | 0,975 | 0,992 | -0,008 (-0,478;0,462) |
| Homocitrulline | -0,04 | 0,968 | 0,992 | -0,01 (-0,504;0,484) |
| L-Leucine | -0,085 | 0,933 | 0,992 | -0,019 (-0,448;0,411) |
| Methionine sulfone | -0,145 | 0,885 | 0,992 | -0,033 (-0,485;0,419) |
| N6,N6,N6-Trimethyl-L-lysine | -0,035 | 0,972 | 0,992 | -0,007 (-0,396;0,382) |
| Butyrylcarnitine | 0,104 | 0,917 | 0,992 | 0,028 (-0,506;0,563) |
| Choline | -0,155 | 0,877 | 0,992 | -0,038 (-0,523;0,446) |
| Deoxycarnitine | -0,104 | 0,917 | 0,992 | -0,016 (-0,321;0,288) |
| Isovalerylcarnitine | -0,075 | 0,94 | 0,992 | -0,018 (-0,487;0,451) |
| Octenoylcarnitine | 0,049 | 0,961 | 0,992 | 0,011 (-0,446;0,469) |
| FA 18(2) | -0,077 | 0,939 | 0,992 | -0,018 (-0,48;0,443) |
| Lysophosphatidylinositol 20(4) | 0,075 | 0,94 | 0,992 | 0,018 (-0,453;0,488) |
| Sphingosine 1-phosphate 18(1) | -0,147 | 0,883 | 0,992 | -0,014 (-0,203;0,175) |
| Taurochenodeoxycholic acid | -0,044 | 0,965 | 0,992 | -0,011 (-0,525;0,502) |
| Taurolithocholic acid | -0,146 | 0,884 | 0,992 | -0,036 (-0,515;0,444) |
| Deoxycholic acid | 0,054 | 0,957 | 0,992 | 0,012 (-0,432;0,456) |
| DGLEA | 0,161 | 0,872 | 0,992 | 0,034 (-0,379;0,447) |
| CE(18:1) | 0,032 | 0,975 | 0,992 | 0,007 (-0,433;0,448) |
| Cer(d18:1/23:0) | 0,121 | 0,904 | 0,992 | 0,033 (-0,503;0,569) |
| Cer(d18:1/24:1) | 0,051 | 0,959 | 0,992 | 0,016 (-0,59;0,621) |
| Lysophosphatidylcholine(16:1) | 0,163 | 0,87 | 0,992 | 0,04 (-0,444;0,525) |
| Lysophosphatidylcholine(18:2) | 0,171 | 0,865 | 0,992 | 0,041 (-0,431;0,513) |
| Lysophosphatidylcholine(18:3) | -0,165 | 0,869 | 0,992 | -0,056 (-0,721;0,609) |
| Phosphatidylcholine(32:0) | -0,194 | 0,846 | 0,992 | -0,043 (-0,476;0,39) |
| Phosphatidylcholine(32:1) | 0,133 | 0,894 | 0,992 | 0,031 (-0,428;0,49) |
| Phosphatidylcholine(32:2) | -0,157 | 0,875 | 0,992 | -0,038 (-0,512;0,436) |
| Phosphatidylcholine(36:5) | 0,146 | 0,884 | 0,992 | 0,038 (-0,467;0,542) |
| Phosphatidylcholine(38:2) | -0,137 | 0,891 | 0,992 | -0,026 (-0,396;0,344) |
| Phosphatidylcholine(38:5) | -0,192 | 0,848 | 0,992 | -0,047 (-0,528;0,433) |
| Phosphatidylcholine(O-34:2) | -0,045 | 0,964 | 0,992 | -0,013 (-0,578;0,552) |
| Phosphatidylcholine(O-34:3) | -0,089 | 0,929 | 0,992 | -0,021 (-0,493;0,45) |
| Phosphatidylethanolamine(O-38:7) | -0,154 | 0,878 | 0,992 | -0,044 (-0,612;0,523) |
| Sphingomyelin(d18:1/18:0) | 0,073 | 0,942 | 0,992 | 0,016 (-0,416;0,449) |
| Sphingomyelin(d18:1/18:1) | 0,181 | 0,856 | 0,992 | 0,035 (-0,345;0,415) |
| Sphingomyelin(d18:1/22:1) | -0,164 | 0,87 | 0,992 | -0,037 (-0,479;0,405) |
| Sphingomyelin(d18:1/24:1) | 0,129 | 0,897 | 0,992 | 0,03 (-0,425;0,485) |
| Sphingomyelin(d18:1/25:0) | 0,131 | 0,896 | 0,992 | 0,031 (-0,427;0,488) |
| TG(55:1) | 0,106 | 0,916 | 0,992 | 0,029 (-0,503;0,56) |
| TG(56:2) | -0,06 | 0,952 | 0,992 | -0,013 (-0,427;0,402) |
| Phosphatidylethanolamine(38:2) | -0,017 | 0,986 | 0,993 | -0,004 (-0,487;0,478) |
| (+/-) 16-HDoHE | 0,008 | 0,993 | 0,997 | 0,001 (-0,296;0,298) |
| Sphingosine 1-phosphate 18(2) | 0,002 | 0,999 | 0,999 | 0 (-0,281;0,281) |

**Supplementary Table 7. Metabolites differently expressed in mitoPD patients with L-Dopa treatment (n = 16) compared to healthy controls (n = 64).** Shown are the results of the fixed effect meta-analysis: regression coefficient (CI: 95% confidence interval), nominal and adjusted p-values.

| **Name** | **z-value** | **Nominal p** | **Adjusted p** | **Beta (CI)** |
| --- | --- | --- | --- | --- |
| 3-Methoxytyrosine | 1,879 | 4.62e-144 | 1.41e-141 | 1,95 (1,873;2,026) |
| Methyldopa | 1,348 | 2.28e-26 | 3.47e-24 | 1,571 (1,339;1,804) |
| (+/-) 16-HDoHE | 0,546 | 0,00177 | 0,108 | 0,809 (0,474;1,144) |
| 5-HETE | 0,913 | 0,00123 | 0,108 | 0,547 (0,309;0,784) |
| 8-HETE | 0,691 | 0,00156 | 0,108 | 0,582 (0,297;0,868) |
| 11-HETE | 0,79 | 0,0057 | 0,26 | 0,493 (0,171;0,814) |
| 15(S)-HETrE | 0,684 | 0,006 | 0,26 | 0,486 (0,158;0,813) |
| L-Tyrosine | 0,743 | 0,007 | 0,266 | 0,403 (0,099;0,708) |
| L-4-hydroxy-proline | 0,741 | 0,0103 | 0,312 | 0,416 (0,099;0,734) |
| Putrescine | 0,619 | 0,00959 | 0,312 | -0,424 (-0,742;-0,105) |
| Sphingosine 1-phosphate 18(1) | 0,327 | 0,0182 | 0,426 | -0,446 (-0,803;-0,089) |
| Prostaglandin F2α | 0,634 | 0,0179 | 0,426 | -0,39 (-0,708;-0,073) |
| TG(60:2) | 0,593 | 0,0164 | 0,426 | -0,367 (-0,67;-0,064) |
| Ornithine | 0,721 | 0,0285 | 0,54 | 0,324 (0,052;0,597) |
| 3-Hydroxypropionic Acid | 0,659 | 0,0302 | 0,54 | 0,377 (0,052;0,703) |
| 8,12-iPF2a IV | 0,543 | 0,0253 | 0,54 | -0,321 (-0,611;-0,031) |
| 8(9)-EpETrE | 0,331 | 0,03 | 0,54 | -0,345 (-0,66;-0,03) |
| Lysophosphatidic acid 16(1) | 0,557 | 0,0351 | 0,575 | -0,266 (-0,513;-0,019) |
| Lysophosphatidylinositol 20(4) | 0,573 | 0,036 | 0,575 | 0,354 (0,016;0,692) |
| Gamma-aminobutyric acid | 0,501 | 0,0621 | 0,589 | -0,361 (-0,711;-0,012) |
| L-Glutamic acid | 0,554 | 0,0433 | 0,589 | -0,295 (-0,587;-0,004) |
| Glycylglycine | 0,562 | 0,0533 | 0,589 | -0,345 (-0,68;-0,009) |
| Sarcosine | 0,469 | 0,0771 | 0,589 | -0,375 (-0,742;-0,007) |
| Trimethylamine N-oxide | 0,369 | 0,0835 | 0,589 | -0,336 (-0,673;0) |
| Sphingosine 1-phosphate 18(0) | 0,244 | 0,0827 | 0,589 | -0,352 (-0,708;0,003) |
| Sphingosine 1-phosphate 18(2) | 0,38 | 0,0522 | 0,589 | -0,232 (-0,568;0,103) |
| 12-HETE | 0,258 | 0,0547 | 0,589 | 0,204 (-0,095;0,503) |
| 14,15-DiHETrE | 0,565 | 0,0497 | 0,589 | -0,216 (-0,506;0,074) |
| Anandamide | 0,699 | 0,0485 | 0,589 | 0,322 (-0,015;0,659) |
| DGLEA | 0,541 | 0,0654 | 0,589 | 0,222 (-0,096;0,54) |
| DG(36:2) | 0,213 | 0,0764 | 0,589 | 0,249 (-0,108;0,606) |
| Lysophosphatidylcholine(16:0) | -0,543 | 0,0563 | 0,589 | -0,265 (-0,619;0,09) |
| Lysophosphatidylcholine(18:0) | -0,528 | 0,0872 | 0,589 | -0,2 (-0,417;0,017) |
| Lysophosphatidylcholine(18:2) | -0,616 | 0,0504 | 0,589 | -0,221 (-0,524;0,082) |
| Lysophosphatidylethanolamine(18:0) | -0,584 | 0,0668 | 0,589 | -0,25 (-0,59;0,09) |
| Lysophosphatidylethanolamine(18:1) | -0,195 | 0,0577 | 0,589 | -0,229 (-0,523;0,066) |
| Phosphatidylcholine(36:1) | -0,432 | 0,0716 | 0,589 | -0,268 (-0,605;0,069) |
| Phosphatidylcholine(38:5) | -0,434 | 0,0617 | 0,589 | -0,256 (-0,58;0,068) |
| Phosphatidylcholine(40:8) | -0,458 | 0,0831 | 0,589 | 0,249 (-0,067;0,566) |
| Phosphatidylethanolamine(38:2) | -0,462 | 0,0693 | 0,589 | 0,25 (-0,095;0,595) |
| Sphingomyelin(d18:1/16:0) | -0,516 | 0,0837 | 0,589 | 0,148 (-0,063;0,358) |
| Sphingomyelin(d18:1/16:1) | -0,483 | 0,078 | 0,589 | -0,21 (-0,522;0,102) |
| TG(56:2) | 0,544 | 0,0832 | 0,589 | -0,287 (-0,623;0,049) |
| TG(58:2) | 0,458 | 0,0662 | 0,589 | 0,214 (-0,095;0,523) |
| TG(59:1) | 0,372 | 0,0856 | 0,589 | 0,199 (-0,07;0,467) |
| Lysophosphatidylcholine(O-16:1) | -0,517 | 0,0898 | 0,593 | 0,291 (-0,057;0,639) |
| Lysophosphatidylcholine(20:5) | -0,424 | 0,093 | 0,602 | 0,292 (-0,039;0,623) |
| DG(36:3) | 0,224 | 0,0973 | 0,616 | -0,325 (-0,698;0,047) |
| TG(56:4) | 0,528 | 0,103 | 0,638 | -0,229 (-0,572;0,113) |
| Sphingomyelin(d18:1/22:1) | -0,454 | 0,108 | 0,656 | -0,253 (-0,61;0,103) |
| Hydroxylysine | 0,52 | 0,12 | 0,658 | -0,256 (-0,622;0,109) |
| Serotonine | -0,3 | 0,122 | 0,658 | -0,242 (-0,601;0,118) |
| Lysophosphatidic acid 14(0) | 0,383 | 0,122 | 0,658 | -0,29 (-0,628;0,048) |
| Lysophosphatidylethanolamine (20:4) | 0,473 | 0,125 | 0,658 | -0,218 (-0,534;0,098) |
| O-Anandamide | 0,519 | 0,128 | 0,658 | -0,299 (-0,666;0,069) |
| Phosphatidylcholine(36:5) | -0,432 | 0,127 | 0,658 | -0,303 (-0,643;0,037) |
| Phosphatidylcholine(38:6) | -0,44 | 0,114 | 0,658 | -0,255 (-0,575;0,064) |
| Phosphatidylcholine(O-34:3) | -0,508 | 0,123 | 0,658 | -0,267 (-0,585;0,051) |
| TG(54:0) | -0,472 | 0,126 | 0,658 | -0,329 (-0,713;0,056) |
| L-2-aminoadipic acid | 0,332 | 0,172 | 0,682 | -0,342 (-0,717;0,034) |
| Cystathionine | 0,317 | 0,16 | 0,682 | -0,369 (-0,765;0,026) |
| O-Acetyl-L-serine | 0,394 | 0,175 | 0,682 | -0,338 (-0,694;0,018) |
| L-Phenylalanine | 0,482 | 0,156 | 0,682 | -0,3 (-0,674;0,074) |
| Saccharopine | 0,422 | 0,152 | 0,682 | -0,328 (-0,704;0,048) |
| Citric acid | 0,38 | 0,181 | 0,682 | -0,248 (-0,595;0,1) |
| FA 18(1) | 0,447 | 0,152 | 0,682 | -0,279 (-0,632;0,074) |
| Lysophosphatidylinositol 18(1) | -0,374 | 0,171 | 0,682 | 0,272 (-0,063;0,607) |
| Palmitoyl ethanolamide | 0,445 | 0,163 | 0,682 | -0,321 (-0,729;0,088) |
| CE(18:2) | -0,438 | 0,161 | 0,682 | 0,253 (-0,106;0,612) |
| CE(20:4) | -0,398 | 0,143 | 0,682 | -0,26 (-0,616;0,095) |
| CE(22:6) | -0,434 | 0,154 | 0,682 | -0,222 (-0,541;0,098) |
| Lysophosphatidylcholine(18:1) | -0,405 | 0,182 | 0,682 | -0,27 (-0,612;0,072) |
| Phosphatidylcholine(32:0) | -0,379 | 0,175 | 0,682 | -0,268 (-0,61;0,073) |
| Phosphatidylcholine(40:6) | -0,363 | 0,181 | 0,682 | -0,331 (-0,672;0,011) |
| Phosphatidylcholine(40:7) | -0,39 | 0,136 | 0,682 | -0,24 (-0,586;0,105) |
| Phosphatidylethanolamine(38:4) | -0,407 | 0,155 | 0,682 | -0,277 (-0,634;0,08) |
| Phosphatidylethanolamine(38:6) | -0,396 | 0,162 | 0,682 | -0,228 (-0,569;0,113) |
| Phosphatidylethanolamine(O-38:7) | -0,446 | 0,151 | 0,682 | -0,296 (-0,645;0,053) |
| Sphingomyelin(d18:1/15:0) | -0,419 | 0,178 | 0,682 | -0,272 (-0,629;0,085) |
| TG(56:1) | 0,283 | 0,17 | 0,682 | -0,334 (-0,698;0,03) |
| TG(56:3) | 0,435 | 0,159 | 0,682 | -0,26 (-0,603;0,084) |
| 9,10,13-TriHOME | 0,314 | 0,187 | 0,693 | -0,25 (-0,623;0,122) |
| 9-HODE | 0,384 | 0,189 | 0,693 | -0,302 (-0,678;0,074) |
| Ethanolamine | 0,353 | 0,2 | 0,722 | -0,315 (-0,684;0,055) |
| FA 22(5) w6 | 0,371 | 0,215 | 0,76 | -0,301 (-0,627;0,025) |
| TG(54:7) | -0,369 | 0,215 | 0,76 | -0,295 (-0,66;0,07) |
| N6,N6,N6-Trimethyl-L-lysine | 0,303 | 0,258 | 0,824 | -0,282 (-0,65;0,085) |
| Choline | 0,404 | 0,25 | 0,824 | -0,331 (-0,698;0,035) |
| Tiglylcarnitine | -0,331 | 0,255 | 0,824 | -0,335 (-0,727;0,056) |
| Malic acid | 0,294 | 0,24 | 0,824 | -0,256 (-0,628;0,116) |
| Lysophosphatidylethanolamine(22:6) | -0,31 | 0,258 | 0,824 | -0,278 (-0,647;0,09) |
| Sphingomyelin(d18:1/23:0) | -0,359 | 0,255 | 0,824 | -0,184 (-0,463;0,094) |
| Sphingomyelin(d18:1/23:1) | -0,342 | 0,256 | 0,824 | -0,192 (-0,482;0,099) |
| Sphingomyelin(d18:1/24:2) | -0,314 | 0,253 | 0,824 | -0,21 (-0,532;0,112) |
| TG(44:2) | 0,31 | 0,242 | 0,824 | -0,215 (-0,549;0,119) |
| Glycodeoxycholic acid | -0,302 | 0,265 | 0,838 | -0,22 (-0,562;0,122) |
| O-Phosphoethanolamine | 0,133 | 0,294 | 0,843 | -0,212 (-0,55;0,126) |
| Isobutyrylcarnitine | -0,311 | 0,308 | 0,843 | 0,212 (-0,126;0,55) |
| Oleoylcarnitine | 0,29 | 0,29 | 0,843 | -0,168 (-0,439;0,103) |
| Isocitrate | 0,253 | 0,276 | 0,843 | -0,205 (-0,546;0,135) |
| Lysophosphatidic acid 16(0) | 0,292 | 0,292 | 0,843 | 0,111 (-0,072;0,294) |
| 11,12-DiHETrE | 0,319 | 0,3 | 0,843 | -0,214 (-0,568;0,14) |
| 13-HODE | 0,28 | 0,294 | 0,843 | -0,193 (-0,512;0,127) |
| Glycoursodeoxycholic acid | 0,366 | 0,275 | 0,843 | -0,211 (-0,558;0,137) |
| CE(18:1) | -0,285 | 0,277 | 0,843 | -0,199 (-0,531;0,133) |
| Lysophosphatidylcholine(14:0) | -0,304 | 0,307 | 0,843 | -0,183 (-0,499;0,133) |
| Lysophosphatidylcholine(22:6) | -0,324 | 0,306 | 0,843 | 0,199 (-0,142;0,539) |
| Phosphatidylcholine(34:2) | -0,294 | 0,287 | 0,843 | -0,169 (-0,464;0,126) |
| Sphingomyelin(d18:1/14:0) | -0,305 | 0,301 | 0,843 | 0,213 (-0,15;0,575) |
| TG(53:1) | 0,376 | 0,286 | 0,843 | -0,192 (-0,528;0,144) |
| TG(55:1) | 0,315 | 0,293 | 0,843 | -0,195 (-0,526;0,136) |
| Lysophosphatidylcholine(O-16:0) | -0,347 | 0,311 | 0,844 | 0,179 (-0,134;0,493) |
| Hexadecenoylcarnitine | 0,265 | 0,315 | 0,846 | -0,198 (-0,535;0,138) |
| (+/-) 8-HDoHE | 0,289 | 0,324 | 0,864 | -0,183 (-0,499;0,133) |
| L-Glutamine | -0,306 | 0,331 | 0,875 | -0,202 (-0,547;0,144) |
| FA 22(6) | -0,285 | 0,335 | 0,879 | -0,218 (-0,606;0,17) |
| Sphingomyelin(d18:1/22:0) | -0,311 | 0,339 | 0,881 | -0,209 (-0,588;0,169) |
| TG(52:1) | -0,261 | 0,343 | 0,884 | -0,195 (-0,552;0,161) |
| L-Alpha-aminobutyric acid | -0,286 | 0,381 | 0,895 | -0,193 (-0,546;0,16) |
| Gamma-Glutamylglutamine | 0,243 | 0,407 | 0,895 | -0,193 (-0,554;0,168) |
| L-Serine | 0,245 | 0,459 | 0,895 | -0,177 (-0,507;0,153) |
| Taurine | 0,113 | 0,473 | 0,895 | -0,182 (-0,523;0,16) |
| Betaine | -0,225 | 0,463 | 0,895 | -0,184 (-0,535;0,167) |
| Dodecenoylcarnitine | 0,235 | 0,412 | 0,895 | -0,173 (-0,504;0,158) |
| Hydroxybutyric acid | 0,182 | 0,471 | 0,895 | -0,196 (-0,571;0,179) |
| 3-Hydroxybutyric acid | 0,242 | 0,432 | 0,895 | 0,179 (-0,168;0,525) |
| Lactic acid | 0,228 | 0,424 | 0,895 | -0,158 (-0,464;0,148) |
| Lysophosphatidic acid 18(1) | 0,227 | 0,387 | 0,895 | 0,163 (-0,158;0,485) |
| Lysophosphatidylethanolamine (16:0) | -0,265 | 0,471 | 0,895 | 0,166 (-0,168;0,499) |
| Lysophosphatidylethanolamine (16:1) | 0,242 | 0,394 | 0,895 | -0,152 (-0,467;0,163) |
| LPS 18(1) | 0,138 | 0,472 | 0,895 | -0,163 (-0,495;0,168) |
| Taurochenodeoxycholic acid | -0,293 | 0,364 | 0,895 | 0,175 (-0,181;0,531) |
| Taurolithocholic acid | -0,227 | 0,477 | 0,895 | -0,17 (-0,516;0,177) |
| (+/-) 10-HDoHE | 0,135 | 0,459 | 0,895 | -0,158 (-0,483;0,168) |
| (+/-) 11-HDoHE | 0,145 | 0,419 | 0,895 | -0,177 (-0,544;0,189) |
| 12,13-DiHODE | -0,301 | 0,42 | 0,895 | -0,185 (-0,556;0,186) |
| 12(S)-HEPE | 0,097 | 0,473 | 0,895 | 0,163 (-0,177;0,504) |
| 17,18-DiHETE | -0,228 | 0,413 | 0,895 | 0,156 (-0,171;0,482) |
| 9,12,13-TriHOME | 0,234 | 0,445 | 0,895 | -0,162 (-0,503;0,179) |
| CHOLIC ACID | 0,22 | 0,405 | 0,895 | 0,182 (-0,208;0,572) |
| Lysophosphatidylcholine(18:3) | -0,268 | 0,437 | 0,895 | -0,17 (-0,538;0,197) |
| Lysophosphatidylcholine(20:4) | -0,259 | 0,4 | 0,895 | 0,158 (-0,186;0,503) |
| Phosphatidylcholine(32:1) | 0,249 | 0,431 | 0,895 | -0,146 (-0,47;0,177) |
| Phosphatidylcholine(36:2) | -0,16 | 0,443 | 0,895 | -0,156 (-0,506;0,193) |
| Phosphatidylcholine(36:4) | -0,246 | 0,353 | 0,895 | -0,162 (-0,532;0,207) |
| Phosphatidylcholine(38:2) | -0,185 | 0,383 | 0,895 | -0,154 (-0,504;0,197) |
| Phosphatidylcholine(38:7) | -0,211 | 0,372 | 0,895 | -0,153 (-0,508;0,201) |
| Phosphatidylcholine(O-36:3) | -0,164 | 0,452 | 0,895 | 0,129 (-0,175;0,432) |
| Phosphatidylethanolamine(36:4) | -0,211 | 0,457 | 0,895 | 0,14 (-0,187;0,467) |
| Phosphatidylethanolamine(O-36:5) | -0,239 | 0,477 | 0,895 | -0,146 (-0,488;0,197) |
| Sphingomyelin(d18:1/18:1) | -0,19 | 0,448 | 0,895 | -0,139 (-0,465;0,187) |
| Sphingomyelin(d18:1/18:2) | -0,156 | 0,452 | 0,895 | -0,157 (-0,531;0,216) |
| Sphingomyelin(d18:1/21:0) | -0,266 | 0,406 | 0,895 | -0,15 (-0,507;0,207) |
| Sphingomyelin(d18:1/24:1) | -0,221 | 0,452 | 0,895 | 0,135 (-0,196;0,465) |
| Sphingomyelin(d18:1/25:0) | -0,224 | 0,394 | 0,895 | 0,142 (-0,205;0,489) |
| TG(42:0) | 0,154 | 0,366 | 0,895 | -0,125 (-0,433;0,183) |
| TG(45:0) | 0,141 | 0,357 | 0,895 | -0,142 (-0,503;0,219) |
| TG(46:2) | 0,236 | 0,411 | 0,895 | 0,117 (-0,216;0,45) |
| TG(48:3) | 0,246 | 0,466 | 0,895 | 0,121 (-0,221;0,463) |
| TG(50:1) | -0,236 | 0,372 | 0,895 | 0,125 (-0,214;0,465) |
| TG(54:3) | 0,244 | 0,423 | 0,895 | 0,126 (-0,209;0,461) |
| TG(56:0) | 0,14 | 0,44 | 0,895 | 0,125 (-0,226;0,477) |
| L-Isoleucine | 0,18 | 0,501 | 0,918 | -0,133 (-0,488;0,222) |
| L-Threonine | 0,209 | 0,507 | 0,918 | -0,125 (-0,453;0,203) |
| L-Tryptophan | -0,197 | 0,499 | 0,918 | -0,108 (-0,404;0,187) |
| Taurocholic acid | -0,228 | 0,497 | 0,918 | -0,122 (-0,46;0,216) |
| 14,15-DiHETE | -0,187 | 0,506 | 0,918 | -0,133 (-0,476;0,211) |
| TG(56:5) | 0,185 | 0,5 | 0,918 | -0,137 (-0,494;0,22) |
| Dehydroepiandrosteron | -0,208 | 0,52 | 0,932 | 0,129 (-0,236;0,494) |
| Sphingomyelin(d18:1/24:0) | -0,202 | 0,521 | 0,932 | -0,135 (-0,515;0,245) |
| Glycine | -0,169 | 0,583 | 0,936 | -0,127 (-0,477;0,224) |
| Homocitrulline | 0,176 | 0,573 | 0,936 | -0,132 (-0,488;0,224) |
| L-Lysine | 0,19 | 0,562 | 0,936 | -0,123 (-0,454;0,207) |
| S-Methylcysteine | -0,192 | 0,542 | 0,936 | -0,13 (-0,482;0,222) |
| L-Valine | -0,147 | 0,58 | 0,936 | 0,118 (-0,222;0,458) |
| Decenoylcarnitine | 0,149 | 0,597 | 0,936 | -0,111 (-0,463;0,24) |
| Tetradecenoylcarnitine | 0,162 | 0,537 | 0,936 | -0,113 (-0,457;0,232) |
| Lysophosphatidylethanolamine (20:5) | -0,165 | 0,594 | 0,936 | 0,107 (-0,217;0,431) |
| 12,13-DiHOME | 0,176 | 0,601 | 0,936 | 0,116 (-0,233;0,464) |
| 8,9-DiHETrE | 0,144 | 0,581 | 0,936 | -0,047 (-0,196;0,102) |
| Cortisol | 0,219 | 0,546 | 0,936 | -0,113 (-0,467;0,241) |
| Glycolithocholic acid | -0,174 | 0,561 | 0,936 | 0,103 (-0,223;0,43) |
| Cer(d18:1/24:1) | 0,185 | 0,574 | 0,936 | -0,104 (-0,411;0,203) |
| Lysophosphatidylcholine(O-18:1) | -0,171 | 0,549 | 0,936 | 0,107 (-0,234;0,448) |
| Phosphatidylcholine(32:2) | -0,156 | 0,597 | 0,936 | -0,115 (-0,48;0,25) |
| Phosphatidylcholine(38:4) | -0,141 | 0,551 | 0,936 | -0,101 (-0,407;0,204) |
| Phosphatidylcholine(O-34:2) | -0,185 | 0,592 | 0,936 | -0,113 (-0,474;0,248) |
| Phosphatidylcholine(O-36:4) | -0,175 | 0,599 | 0,936 | -0,112 (-0,441;0,217) |
| Phosphatidylcholine(O-36:5) | -0,183 | 0,547 | 0,936 | -0,117 (-0,481;0,247) |
| Phosphatidylethanolamine(34:2) | -0,148 | 0,559 | 0,936 | -0,094 (-0,398;0,211) |
| Sphingomyelin(d18:1/20:0) | -0,157 | 0,596 | 0,936 | -0,102 (-0,442;0,238) |
| TG(42:2) | 0,173 | 0,551 | 0,936 | -0,099 (-0,449;0,251) |
| TG(46:1) | 0,18 | 0,553 | 0,936 | -0,085 (-0,388;0,219) |
| TG(51:2) | -0,152 | 0,599 | 0,936 | -0,089 (-0,407;0,229) |
| TG(56:7) | -0,181 | 0,529 | 0,936 | 0,092 (-0,216;0,4) |
| Nonaylcarnitine | -0,161 | 0,608 | 0,939 | 0,095 (-0,238;0,428) |
| 9-HOTrE | 0,156 | 0,608 | 0,939 | -0,082 (-0,364;0,2) |
| TG(54:4) | 0,163 | 0,612 | 0,939 | -0,092 (-0,403;0,22) |
| Butyrylcarnitine | 0,143 | 0,629 | 0,953 | 0,089 (-0,231;0,41) |
| TG(42:1) | 0,113 | 0,629 | 0,953 | -0,114 (-0,489;0,261) |
| TG(46:0) | -0,126 | 0,63 | 0,953 | -0,108 (-0,472;0,256) |
| Phosphatidylcholine(34:3) | -0,135 | 0,638 | 0,955 | 0,106 (-0,268;0,48) |
| TG(58:8) | -0,117 | 0,636 | 0,955 | -0,109 (-0,484;0,265) |
| Pimelylcarnitine | 0,131 | 0,641 | 0,956 | 0,101 (-0,266;0,468) |
| Octanoylcarnitine | 0,123 | 0,65 | 0,959 | -0,056 (-0,261;0,149) |
| Glycocholic acid | -0,141 | 0,656 | 0,959 | -0,081 (-0,39;0,228) |
| TG(50:2) | -0,123 | 0,648 | 0,959 | -0,082 (-0,392;0,228) |
| TG(56:6) | -0,116 | 0,656 | 0,959 | 0,089 (-0,254;0,432) |
| 9,10-DiHOME | 0,128 | 0,666 | 0,969 | 0,084 (-0,249;0,417) |
| DL-3-aminoisobutyric acid | 0,136 | 0,671 | 0,971 | -0,089 (-0,437;0,26) |
| Glutathione | 0,1 | 0,691 | 0,971 | 0,089 (-0,261;0,438) |
| L-Histidine | 0,105 | 0,733 | 0,971 | -0,089 (-0,434;0,256) |
| L-Leucine | 0,096 | 0,722 | 0,971 | -0,089 (-0,432;0,254) |
| Methionine sulfone | -0,124 | 0,692 | 0,971 | -0,086 (-0,43;0,258) |
| 2-Methylbutyroylcarnitine | -0,104 | 0,73 | 0,971 | 0,084 (-0,26;0,428) |
| Linoleylcarnitine | -0,095 | 0,739 | 0,971 | -0,051 (-0,259;0,156) |
| Malonylcarnitine | 0,114 | 0,686 | 0,971 | -0,084 (-0,431;0,263) |
| Propionylcarnitine | 0,108 | 0,722 | 0,971 | 0,022 (-0,069;0,114) |
| Tetradecadienylcarnitine | 0,09 | 0,738 | 0,971 | -0,069 (-0,377;0,24) |
| Valerylcarnitine | 0,11 | 0,716 | 0,971 | 0,075 (-0,263;0,412) |
| Pyroglutamic acid | 0,097 | 0,734 | 0,971 | -0,037 (-0,205;0,13) |
| FA 18(2) | -0,109 | 0,74 | 0,971 | -0,074 (-0,392;0,243) |
| Lysophosphatidic acid 18(2) | 0,092 | 0,72 | 0,971 | -0,081 (-0,443;0,28) |
| Taurodeoxycholic acid | -0,114 | 0,741 | 0,971 | 0,078 (-0,259;0,414) |
| Glycochenodeoxycholic acid | -0,112 | 0,705 | 0,971 | -0,067 (-0,358;0,224) |
| Linoleoyl ethanolamide | 0,12 | 0,711 | 0,971 | -0,073 (-0,401;0,255) |
| Thromboxane-B2 | -0,086 | 0,733 | 0,971 | -0,088 (-0,479;0,304) |
| Phosphatidylcholine(O-34:1) | -0,095 | 0,738 | 0,971 | 0,071 (-0,267;0,409) |
| Phosphatidylcholine(O-38:6) | -0,117 | 0,717 | 0,971 | -0,027 (-0,161;0,107) |
| Phosphatidylcholine(O-44:5) | -0,13 | 0,699 | 0,971 | -0,075 (-0,445;0,295) |
| Phosphatidylethanolamine(O-38:5) | -0,134 | 0,703 | 0,971 | 0,071 (-0,28;0,422) |
| TG(48:1) | -0,107 | 0,722 | 0,971 | -0,027 (-0,163;0,109) |
| Phosphatidylcholine(34:1) | -0,025 | 0,746 | 0,974 | -0,069 (-0,419;0,281) |
| Citrulline | -0,075 | 0,753 | 0,975 | -0,062 (-0,39;0,267) |
| TG(54:2) | 0,092 | 0,754 | 0,975 | 0,061 (-0,262;0,384) |
| Homocysteine | -0,088 | 0,759 | 0,975 | 0,07 (-0,297;0,436) |
| Stearoylcarnitine | 0,082 | 0,761 | 0,975 | -0,06 (-0,397;0,277) |
| 1-Methylhistidine | -0,044 | 0,883 | 0,982 | -0,059 (-0,387;0,269) |
| L-Alanine | -0,042 | 0,889 | 0,982 | 0,049 (-0,232;0,329) |
| L-Arginine | 0,057 | 0,841 | 0,982 | 0,05 (-0,249;0,349) |
| Cysteine | -0,069 | 0,831 | 0,982 | -0,052 (-0,365;0,26) |
| Gamma-L-glutamyl-L-alanine | 0,054 | 0,84 | 0,982 | 0,051 (-0,308;0,41) |
| L-Kynurenine | -0,074 | 0,81 | 0,982 | 0,054 (-0,291;0,399) |
| L-Methionine | 0,049 | 0,857 | 0,982 | 0,044 (-0,254;0,343) |
| L-Proline | 0,066 | 0,853 | 0,982 | 0,042 (-0,246;0,329) |
| Symmetric dimethylarginine | -0,04 | 0,904 | 0,982 | -0,028 (-0,224;0,168) |
| Carnitine | 0,091 | 0,807 | 0,982 | -0,056 (-0,413;0,301) |
| Decanoylcarnitine | 0,047 | 0,859 | 0,982 | -0,047 (-0,364;0,27) |
| Deoxycarnitine | -0,033 | 0,866 | 0,982 | -0,054 (-0,392;0,284) |
| Hexanoylcarnitine | 0,057 | 0,836 | 0,982 | -0,056 (-0,42;0,309) |
| Lauroylcarnitine | -0,036 | 0,89 | 0,982 | -0,056 (-0,428;0,316) |
| Methylmalonylcarnitine | -0,044 | 0,883 | 0,982 | -0,055 (-0,44;0,33) |
| Palmitoylcarnitine | -0,073 | 0,771 | 0,982 | -0,044 (-0,361;0,272) |
| Lysophosphatidylinositol 16(1) | 0,052 | 0,867 | 0,982 | -0,039 (-0,337;0,259) |
| Lysophosphatidylinositol 18(2) | 0,073 | 0,791 | 0,982 | -0,038 (-0,376;0,3) |
| 1-LG/2-LG | -0,043 | 0,877 | 0,982 | 0,037 (-0,281;0,356) |
| (+/-) 14-HDoHE | 0,036 | 0,779 | 0,982 | -0,016 (-0,16;0,127) |
| Prostaglandin E2 | 0,045 | 0,857 | 0,982 | 0,017 (-0,133;0,167) |
| Cer(d18:0/22:0) | -0,037 | 0,899 | 0,982 | 0,043 (-0,285;0,371) |
| Cer(d18:1/22:0) | 0,048 | 0,85 | 0,982 | 0,041 (-0,318;0,399) |
| Cer(d18:1/23:0) | 0,095 | 0,774 | 0,982 | -0,045 (-0,407;0,317) |
| Lysophosphatidylcholine(16:1) | 0,074 | 0,793 | 0,982 | -0,046 (-0,412;0,32) |
| Lysophosphatidylethanolamine(20:4) | -0,064 | 0,818 | 0,982 | -0,04 (-0,386;0,306) |
| Phosphatidylcholine(34:4) | -0,049 | 0,874 | 0,982 | 0,044 (-0,327;0,416) |
| Phosphatidylcholine(36:3) | -0,08 | 0,784 | 0,982 | -0,046 (-0,404;0,312) |
| Phosphatidylcholine(38:3) | 0,067 | 0,811 | 0,982 | 0,035 (-0,3;0,371) |
| Phosphatidylcholine(O-38:4) | 0,04 | 0,895 | 0,982 | 0,036 (-0,308;0,38) |
| Phosphatidylcholine(O-38:5) | -0,073 | 0,825 | 0,982 | -0,034 (-0,357;0,289) |
| Sphingomyelin(d18:1/18:0) | 0,06 | 0,812 | 0,982 | 0,034 (-0,301;0,369) |
| TG(44:0) | -0,029 | 0,896 | 0,982 | -0,033 (-0,368;0,302) |
| TG(44:1) | 0,06 | 0,825 | 0,982 | -0,037 (-0,414;0,339) |
| TG(48:2) | 0,034 | 0,911 | 0,982 | -0,033 (-0,375;0,31) |
| TG(51:1) | 0,051 | 0,871 | 0,982 | -0,022 (-0,267;0,223) |
| TG(51:4) | 0,089 | 0,776 | 0,982 | -0,033 (-0,397;0,331) |
| TG(52:2) | -0,029 | 0,91 | 0,982 | -0,026 (-0,331;0,279) |
| TG(52:3) | 0,073 | 0,796 | 0,982 | -0,028 (-0,361;0,306) |
| TG(54:1) | -0,031 | 0,909 | 0,982 | 0,023 (-0,295;0,341) |
| TG(54:5) | 0,038 | 0,904 | 0,982 | -0,023 (-0,338;0,291) |
| TG(57:1) | -0,056 | 0,806 | 0,982 | 0,012 (-0,139;0,162) |
| TG(58:9) | -0,042 | 0,865 | 0,982 | -0,027 (-0,39;0,336) |
| TG(60:1) | -0,078 | 0,796 | 0,982 | 0,026 (-0,341;0,392) |
| TG(60:3) | 0,065 | 0,837 | 0,982 | 0,019 (-0,273;0,312) |
| Acetylcarnitine | 0,023 | 0,939 | 0,989 | 0,02 (-0,311;0,352) |
| Isovalerylcarnitine | -0,026 | 0,929 | 0,989 | -0,018 (-0,348;0,312) |
| Myristoilcarnitine | -0,018 | 0,939 | 0,989 | 0,019 (-0,344;0,381) |
| 19,20-DiHDPA | 0,022 | 0,942 | 0,989 | -0,02 (-0,385;0,344) |
| Lysophosphatidylcholine(20:3) | 0,02 | 0,939 | 0,989 | 0,019 (-0,352;0,39) |
| Phosphatidylcholine(40:5) | 0,021 | 0,938 | 0,989 | -0,018 (-0,395;0,358) |
| Sphingomyelin(d18:1/25:1) | -0,022 | 0,943 | 0,989 | -0,013 (-0,325;0,298) |
| TG(50:4) | 0,031 | 0,923 | 0,989 | -0,014 (-0,351;0,323) |
| L-Asparagine | 0,018 | 0,954 | 0,989 | 0,015 (-0,345;0,375) |
| Lysophosphatidylinositol 18(0) | -0,017 | 0,953 | 0,989 | -0,005 (-0,346;0,337) |
| 12,13-EpOME | 0,022 | 0,947 | 0,989 | 0,01 (-0,313;0,332) |
| Sphingomyelin(d18:1/20:1) | 0,013 | 0,957 | 0,989 | 0,004 (-0,342;0,35) |
| TG(52:4) | -0,014 | 0,962 | 0,991 | 0,009 (-0,335;0,354) |
| L-Homoserine | -0,004 | 0,987 | 0,994 | -0,007 (-0,315;0,301) |
| Octenoylcarnitine | 0,011 | 0,97 | 0,994 | -0,005 (-0,289;0,28) |
| 2-ketoglutaric acid | 0,008 | 0,977 | 0,994 | 0,005 (-0,323;0,332) |
| Deoxycholic acid | 0,005 | 0,984 | 0,994 | -0,004 (-0,321;0,313) |
| Cer(d18:0/23:0) | 0,006 | 0,986 | 0,994 | -0,003 (-0,344;0,337) |
| TG(50:3) | 0,01 | 0,973 | 0,994 | 0,003 (-0,34;0,347) |
| TG(51:3) | -0,007 | 0,982 | 0,994 | -0,007 (-0,388;0,374) |
| 1-AG/2-AG | 0,002 | 0,994 | 0,997 | -0,007 (-0,366;0,351) |
| 5,6-DiHETrE | 0 | 0,999 | 0,999 | 0,007 (-0,358;0,372) |
